# *TARGET-AI*: a foundational approach for the targeted deployment of artificial intelligence electrocardiography in the electronic health record

**DOI:** 10.1101/2025.08.25.25334266

**Authors:** Evangelos K. Oikonomou, Bruno Batinica, Lovedeep S. Dhingra, Arya Aminorroaya, Andreas Coppi, Rohan Khera

## Abstract

**Background:** Artificial intelligence (AI) applied to routine electrocardiograms (ECGs) offers promise for screening of structural heart disease (SHD), yet broad clinical integration remains limited by high false positive rates and the lack of tailored deployment strategies.

**Methods:** We developed TARGET-AI, a multimodal AI-enabled pipeline that integrates longitudinal electronic health record (EHR) data with ECG images to identify optimal intersections of healthcare encounters and patient phenotypes for targeted AI-ECG screening of SHD. The approach is built on (1) a pretrained EHR foundation model (CLMBR-T) applied to 118 million coded events from 159,322 individuals to generate temporal patient embeddings and identify high-risk screening candidates, followed by (2) a novel contrastive vision-language model trained on 754,533 ECG image-echocardiogram report pairs to detect SHD subtypes with tunable performance characteristics. We evaluated this sequential, gated strategy in 5,198 individuals referred for their first transthoracic echocardiogram (TTE) within 90 days of an ECG (temporal validation), as well as in geographically distinct cohorts, including 33,518 UK Biobank participants undergoing protocolized ECG and cardiac magnetic resonance imaging, and a geographically distinct inpatient EHR cohort of 3,628 patients with ECG-TTE pairs (MIMIC-IV). Significance was determined by comparing metric differences between targeted and untargeted strategies, with bootstrap-derived 95% confidence intervals excluding zero considered significant.

**Results:** Our pre-trained AI-ECG image foundation model discriminated 26 SHD subtypes, including left ventricular systolic dysfunction (AUROC of 0.90), severe aortic stenosis (AUROC of 0.85) and elevated right ventricular systolic pressure (AUROC of 0.82). Compared with untargeted AI-ECG screening, targeted screening in the temporal validation set (n=5,198) was associated with a significant increase in F1 scores (median of 0.25 [range: 0.09 to 0.75]) and decrease in false positives (median of –303 [range: –715 to –77]) across 26 SHD labels. Similar increases in F1 scores and reductions in false positives were seen in the UK Biobank (n=33,518; median change in false positives of –819 [range: –3,521 to –459] across 7 SHD labels) and MIMIC-IV (n=3,628; median false positive change of –255 [range: –716 to –86] across 5 SHD labels).

**Conclusions:** TARGET-AI may guide the targeted deployment of AI-ECG for SHD screening by integrating longitudinal EHR phenotypes with multimodal ECG-echocardiogram representations in an interoperable framework, enabling adaptive, data-driven screening strategies across health systems.

## INTRODUCTION

Artificial intelligence (AI) technologies hold promise for the scalable detection of structural heart disease (SHD) from routine cardiac diagnostic tests, such as traditional 12-lead electrocardiograms (ECG).^1,2^ Nevertheless, evidence remains limited on how best to integrate these tools into clinical care pathways.^3,4^ Two key restrictions, in particular, limit the uptake of these technologies within the electronic health records (EHR) of large health systems. First, the prevalence of SHD in unselected cohorts is low, and may lead to high false discovery rates,^5^ unnecessary testing, patient anxiety and increased downstream costs.^6–8^ Second, any system intended to guide the targeted deployment of AI-ECG technologies for cardiac screening must exhibit foundational capabilities that allow flexible adaptation to different forms of SHD and diverse patient populations.^9^ Despite the emergence of foundation models that learn global representations of health and disease across modalities,^10^ including ECG,^11^ transthoracic echocardiography (TTE),^12^ and EHR records,^13,14^ there is currently no systematic strategy that integrates these data sources to dynamically estimate the pre-test probability of SHD and guide the targeted deployment of AI-ECG.

To bridge this gap, we propose *TARGET-AI*, a foundational, multi-modal approach that learns interoperable, longitudinal representations from the EHR, alongside ECG image embeddings that have been contrastively pre-trained against linked echocardiograms, to guide the deployment of AI-ECG screening in health systems. We first deployed this system in a cohort of more than 150,000 unique individuals, contributing nearly 18 million EHR events and more than 700,000 ECG-TTE pairs. We then tested the model in independent, temporally distinct subsets from the same cohort, as well as in two geographically distinct populations: (1) hospitalized patients admitted through the emergency department or intensive care unit of a tertiary hospital (MIMIC-IV) and (2) participants of the UK Biobank, a community-based cohort with standardized ECG and cardiac MRI imaging. Consistent with the overarching goal of reducing false discovery, we evaluated the system’s ability to improve the balance of precision and recall across multiple “hidden” SHD labels. Conceptually, this enabled us to evaluate two alternative clinical deployment strategies (**Fig. 1**): (i) an *untargeted* approach, where AI-ECG models are applied opportunistically to all individuals with available ECGs; and (ii) a *targeted* strategy, in which deployment is guided by data-driven estimates of disease likelihood derived from personalized EHR trajectories.

**Fig. 1:**
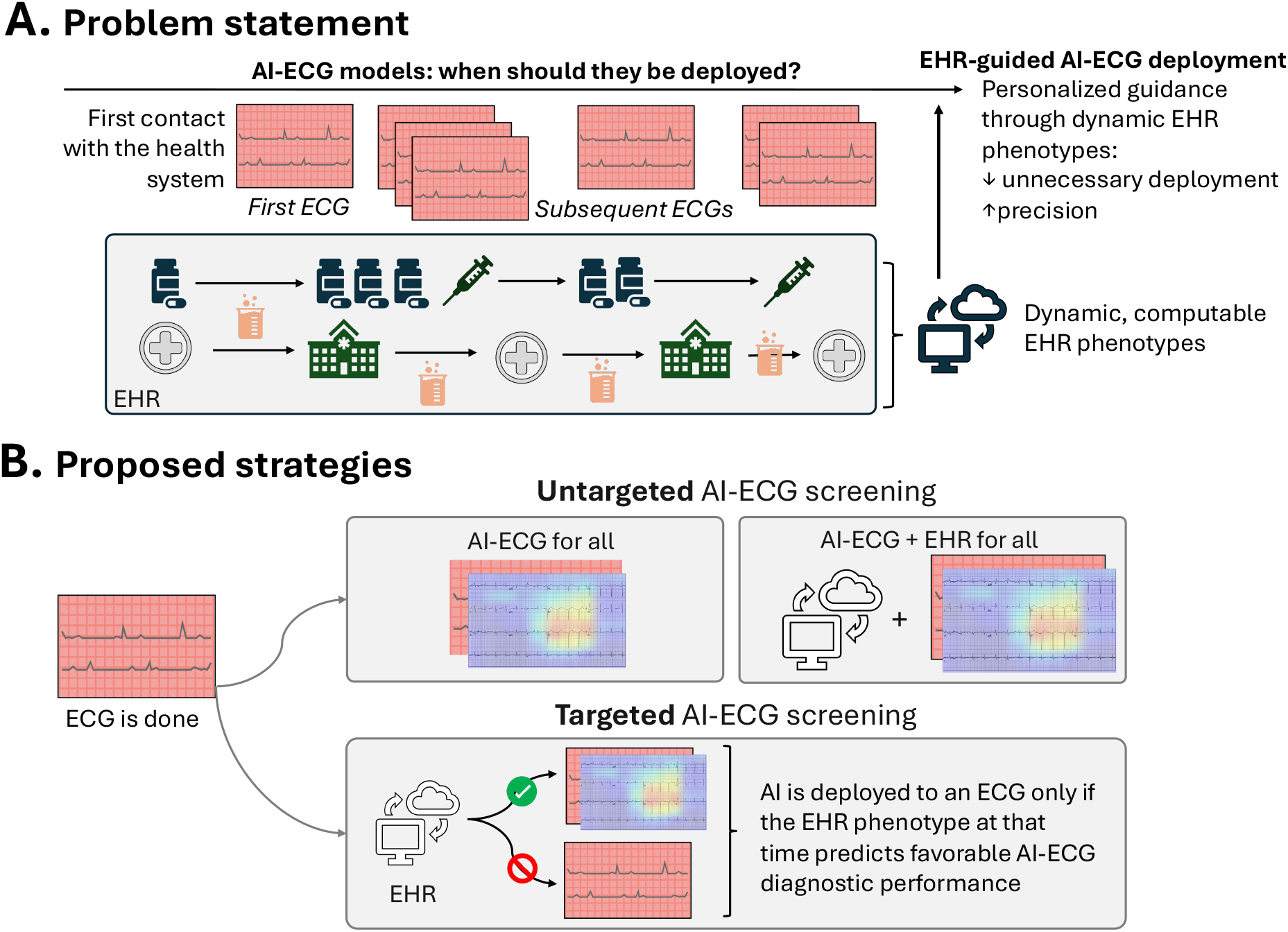
Targeted deployment of AI screening technologies in the EHR. **(A) Problem statement:** Across their longitudinal care in an integrated health system, individuals may receive multiple diagnostic tests, including standard 12-lead electrocardiograms (ECG). ECG phenotypes evolve in tandem with the broader clinical profile of an individual, as recorded and computed in the electronic health record (EHR). Artificial intelligence (AI)-guided technologies may screen for structural heart disease (SHD) directly from ECGs, but there is currently no data-driven strategy to enable their targeted deployment, to minimize false discovery and unnecessary downstream testing. **(B) Potential strategies:** The integration of AI-ECG technologies may follow different strategies. For example, AI-ECG can be deployed opportunistically across all eligible ECGs with or without incorporating EHR-derived clinical characteristics into the modeling (*“untargeted strategy”*). Alternatively, the computable phenotypes of individuals in the EHR may be leveraged as gatekeepers to prevent unnecessary deployment of AI-ECG, potentially optimizing the precision of the technology (“*targeted strategy*”). *AI: artificial intelligence; ECG: electrocardiography; EHR: electronic health record*.

## METHODS

### Data Source and Population

This retrospective study was approved by the Yale Institutional Review Board (IRB) with a waiver of informed consent. UK Biobank analyses were conducted under project #71033.

An extended version of the methods can be found in the **Supplement**. Briefly, we queried the Yale-New Haven Health System (YNHHS) EHR (2013-2023) to identify adults (≥18 years) who underwent a 12-lead ECG and TTE within 90 days (±). In total, we assembled 754,533 ECG-TTE pairs from 159,322 individuals (*2013-2021*) for model training, with a cap set at 10 ECGs per person. A held-out test set included 8,979 individuals with a single ECG-TTE pair per participant (*2022-2023*). To model a temporally valid screening strategy, we restricted this to 5,198 patients undergoing their first known TTE in our system up to 90 days *after* the index ECG.

In the UK Biobank, we identified 33,518 participants with protocolized 12-lead ECG and CMR imaging (assessment visit #2). Demographic and clinical data were extracted from structured fields and protocol-derived labs. Imaging labels were based on automated CMR measurements, using established thresholds for key parameters, such as LVEF ≤40%, or cohort-specific 99.5^th^ percentiles for volumetric and linear measurements.^15^

Finally, to further evaluate the generalizability of this approach to guide locally-adapted systems for targeted AI-ECG screening, we queried the MIMIC-IV EHR dataset for available discharge summaries and identified a total of 5,231 individuals (≥18 years) with embedded TTE reports as well as ECGs performed within 90 days.^16,17^

### Data Preprocessing and Label Definition

In all cohorts ECG images were generated at random across four common layouts (standard, alternate, standard shuffled and alternate shuffled) to enhance generalizability, using a pipeline and augmentations described and validated in our previous work.^18–21^ In YNHHS, unstructured (textual) and structured TTE reports were exported from the local reporting software (Lumedx, Oakland, CA).^22^ To cover the broad spectrum of SHD phenotypes, we defined 26 discrete SHD labels, spanning parameters of left and right ventricular function, size, thickness, atrial anatomy, valvular function and related measurements (**Table S1**). In the UK Biobank, CMR-derived phenotypes were computed using automated fields from Bai et al.^15^ In the MIMIC-EHR, we used our previously published HeartDX-LLM algorithm to transform text reports extracted through the available discharge summaries into structured fields.^23^ Unless specified otherwise, SHD definitions incorporated standard cutoffs and population-based thresholds (see ***Supplement***).^24^

### Extracting EHR Embeddings

To encode longitudinal EHR representations we used CLMBR-T (**Fig. S1**), a 141M parameter autoregressive foundation model pre-trained on 2.57M EHR records from Stanford Medicine.^13,14^ CLMBR-T models a sequence of coded medical events, spanning diagnoses, medications, laboratory measurements, procedures, and encounter history, based on the Observational Medical Outcomes Partnership (OMOP) Common Data Model (CDM) vocabulary and MEDS (Medical Event Data Standard) schema. For each ECG, a MEDS-compatible event sequence was generated using all events recorded in the EHR from its inception until the timepoint the ECG was performed. We extracted the final 768-dimensional hidden layer as a representation of a patient’s longitudinal profile. This architecture treats each patient’s record as a temporal narrative of medical events, allowing the transformer to learn from contextual patterns and account for missingness, effectively capturing both observed and unobserved elements in the sequence without explicit imputation. In total, we mapped 26,828 [96.0%] of the original 27,946 non-laboratory-related codes using Unified Medical Language System (UMLS) resources (e.g., SNOMED CT, RxNorm) (**Tables S2-S3**). In addition to age, sex, race and ethnicity, these spanned a comprehensive list of 1,982 diagnosis concepts, 213 medications, 25,771 medical procedures, and 37 commonly performed laboratory measurements across 3 possible encounter settings (inpatient, ED/observation, outpatient).

### Contrastive ECG-TTE Vision-Text Model Training

To learn echocardiography-defined SHD representations from ECG images, we trained a CLIP (Contrastive Language-Image Pretraining)-style model to align ECG images with TTE reports using a dual encoder architecture (**Fig. 2a**). In this framework, the “ground truth” supervisory signal is the linkage between an ECG and its paired echocardiography report: matching ECG-TTE pairs are treated as positives, whereas all other non-matching combinations within a batch are treated as negatives. First, we defined patient-level splits (95% train, 5% validation), with a maximum of 10 ECGs per patient and randomized layout augmentation. Next, we initialized our ECG image (vision) encoder using BEiT-base-patch 16-384 ViT (vision transformer; 12 layers, 768 hidden dimensions),^25^ whereas for the text encoder we used a CLIP text transformer (512 hidden dimensions, 256 tokens). To account for the limited vocabulary found in TTE reports, we trained a custom BPE (byte-pair encoding) tokenizer with a final vocabulary size of 16,030 and a minimum token frequency of 3. We trained the contrastive model in up to 8 NVIDIA H100 graphics processing units (GPUs) for a total of 100 epochs using the AdamW optimizer, a 10^-5^ learning rate, linear warm-up, a cosine decay schedule, mixed-precision training to reduce memory usage, and a batch size of 64. The loss function was the symmetric cross-entropy objective, as introduced in CLIP,^26^ which operates directly on pairwise cosine similarities between image and text embeddings; these similarity values are directional measures of alignment and are not probability-scaled. Performance during training was tracked by the median percentile rank of true report retrieval via cosine similarity for ECG-TTE pairs in the validation set. We did not explicitly account for clustering at the patient level during training; however, across all downstream testing sets included only 1 ECG per unique participant.

**Fig. 2:**
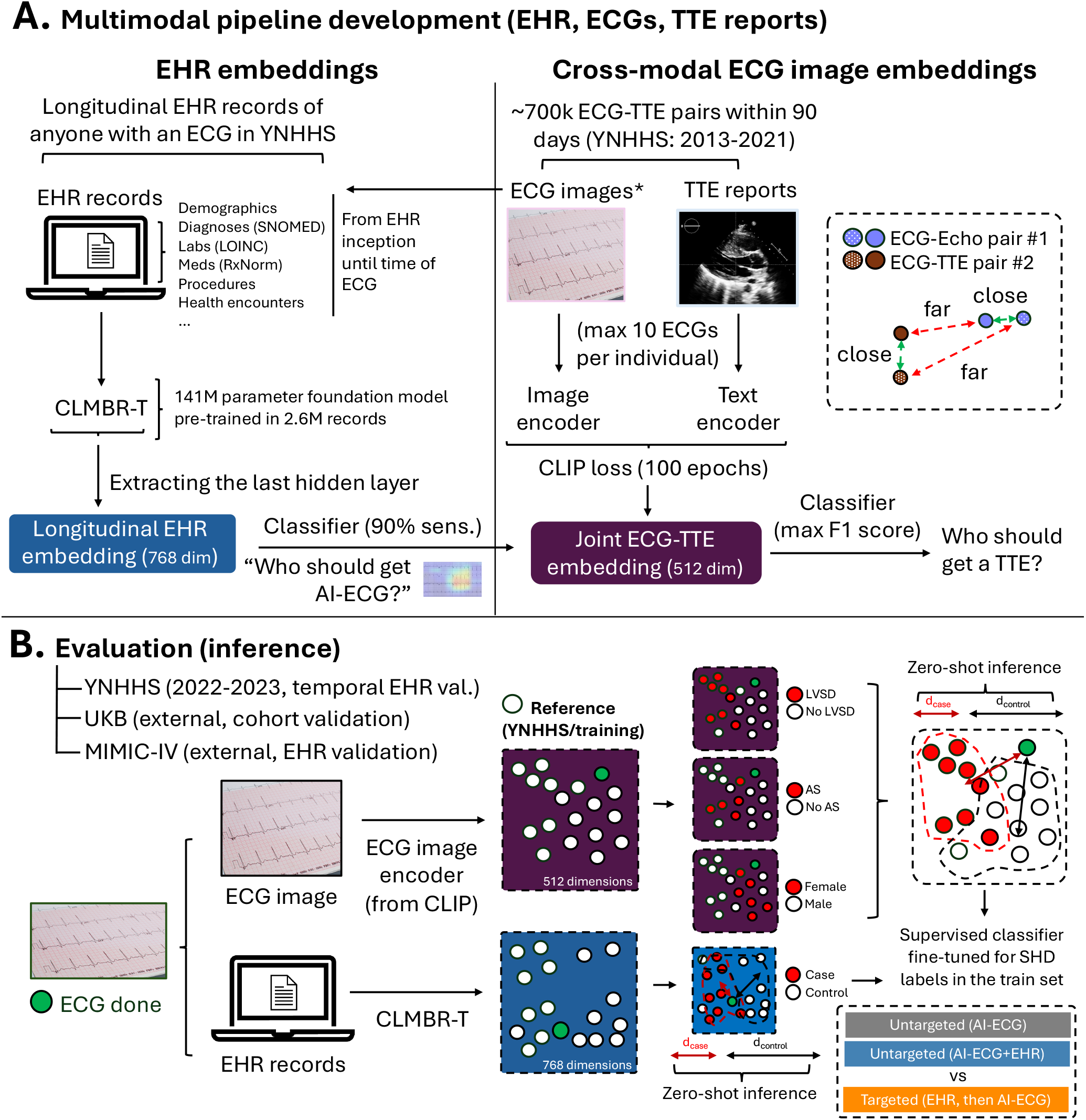
Study design. **(A) Training:** To evaluate the feasibility and performance of leveraging the EHR for the targeted screening of SHD by AI-ECG, we designed a strategy that jointly leverages two foundation models. First, a pre-trained foundation model built for longitudinal EHR phenotypes (CLMBR-T) is used to define an embedding of an individual’s longitudinal EHR trajectory up until the time an ECG is performed. Second, we apply contrastive pre-training to learn a joint embedding between ECG images (using a vision transformer) and text embeddings of corresponding TTE reports performed within 90 days of each other. This allows us to extract a 768-dimensional embedding reflecting an individual’s EHR history up until that point, as well as a 512-dimensional embedding reflecting structural correlates from the ECG (image). These can be combined to define a sequential, gated paradigm for targeted AI-ECG deployment. **(B) Evaluation:** The models are evaluated in temporally and geographically distinct datasets of individuals not seen during training. We examine both the zero-shot performance of the embeddings in identifying discrete anomalies by leveraging the relative cosine similarity of a new case versus the reference embeddings of individuals with and without the condition of interest in the training set. Finally, we evaluate the performance of a targeted vs untargeted AI-ECG screening strategy using the fine-tuned sequential “*targeted*” (EHR pre-screening, followed by AI-ECG) or “*untargeted*” strategies (AI-ECG for all). *AI: artificial intelligence; CLIP: Contrastive Language-Image Pre-training; CLMBR: clinical language model-based representation – transformer; ECG: electrocardiography; EHR: electronic health record; MIMIC-IV: Medical Information Mart for Intensive Care; SHD: structural heart disease; TTE: transthoracic echocardiography; YNHHS: Yale-New Haven Health System*.

### Zero-Shot and Few-Shot Classification of SHD from ECG images

For each SHD phenotype, embeddings from positive (case) and negative (control) training cases at YNHHS were averaged to create centroids, defined as the mean vector across positive (or negative) observations in the training dataset (**Fig. 2b**). For unseen data in the temporally distinct test set (*2022-2023*), we computed embeddings by passing the index EHR records and ECG images through the pretrained CLMBR-T and ViT models, respectively. For each example in the YNHHS test set, we calculated cosine similarities to the two reference centroids derived from the training set: one representing known positive cases and the other known controls. The final classification score was defined as the arithmetic difference in similarity to the positive versus the negative centroid. We used this score to estimate the area under the receiver operating characteristic curve (AUROC) separately for each label. For CLMBR-T embeddings, we further performed an ablation experiment masking key demographics (sex, race, ethnicity) and extracting reference embeddings without including this key information. Finally, to evaluate the foundational properties of the ECG image ViT model, we used progressively larger sets of cases and controls from the publicly released subset of the EchoNext training set, including pairs of ECG and TTEs from New-York Presbyterian Hospital/Columbia University Medical Center, and benchmarked its performance for the available SHD labels in the corresponding testing set.^27–29^ Critically, this was done without retraining the model, but rather defining cohort-specific reference centroids based on the respective EchoNext training set.

### Targeted vs Untargeted Screening Strategies

We operationalized two deployment paradigms for AI-ECG evaluation: an *untargeted* strategy that broadly applies AI-ECG to all individuals with available data, and a *targeted* strategy that prioritizes individuals based on their EHR-derived disease likelihood and ECG characteristics. In the *untargeted* approach, a simple logistic regression classifier was trained solely on ECG ViT embeddings and applied to every patient. In the *targeted* (sequential) approach, individuals were first scored using a high-recall logistic classifier added on top of the EHR (CLMBR-T) embeddings; only those exceeding a ≥90% sensitivity threshold (per the training set) proceeded to the ECG ViT classifier, while all others were automatically classified as screen-negative. As part of the untargeted paradigm, we additionally examined an integrated model concatenating ECG ViT and CLMBR-T embeddings for direct classification. Model thresholds for AI-ECG (across strategies) were tuned on the training set via F1 optimization. In the targeted strategy, anyone below the EHR CLMBR-T threshold was trated as a negative screen, and no AI-ECG inference was run. During internal validation at YNHHS and external validation in the UK Biobank, the regression models were directly deployed without recalibration. Due to differences in the study population and SHD label definition in MIMIC-IV, a hospital-based cohort of patients seen in the emergency room or admitted to the intensive care unit of the Beth Israel Deaconess Medical Center in Boston, MA,^16,17^ we evaluated the adaptability of this framework using data collected before 2011 to guide targeted AI-ECG deployment during the later years. Across all analyses the CLMBR-T and ECG ViT weights were frozen, ensuring consistency in the produced embeddings.

### Statistical Analysis

We summarize continuous variables using medians [25^th^-75^th^ percentiles, or minimum-maximum range, as specified], and categorical variables as counts (percentages). The zero-shot discrimination performance of the different strategies for each one of the 26 SHD labels was first evaluated using the respective AUROC.^30^ Logistic regression models were trained on the standardized feature embeddings (with z-score normalization) using an L2-regularized objective (ridge regression) for stable convergence in high-dimensional feature embeddings. Ridge regularization was applied to penalize large coefficient magnitudes, reducing overfitting and improving model generalizability across cohorts. All binary thresholds for classification were derived solely in the training set: F1 optimization was used to maximize precision-recall balance for the ECG ViT classifiers, while the gating threshold for EHR CLMBR-T-based classifiers in the targeted strategy was chosen to meet a recall (sensitivity) of ≥90%, as described above. Cutoffs were determined separately for the targeted and untargeted strategies: in the targeted design, thresholds were first optimized for the EHR model to define the subset undergoing downstream ECG evaluation, followed by independent threshold selection for the ECG model within that subset. To quantify uncertainty, we used 500 paired bootstrap resamples to derive 95% confidence intervals (CIs) for absolute performance metrics and their pairwise differences. These CIs are conditional on the trained prediction model and do not incorporate uncertainty from model selection or development. For pairwise comparisons and absolute differences in metrics (i.e., F1-score, false positive counts) statistical significance was defined as a 95% CI excluding zero, without correction for multiple comparisons. All statistical analyses were performed in Python 3.9.

## RESULTS

### Data Source and Study Population

The *development cohort* consisted of 159,322 adult individuals across YNHHS (2013-2021). In total, we extracted 754,533 unique ECG-TTE pairs, in addition to a list of 118,734,437 eligible EHR-coded events from the same individuals. The training set included a balanced representation of men and women (80,275 [50.4%] female), with nearly 1 in 4 (38,062 [23.9%]) self-reporting non-White race. A separate dataset of 8,979 individuals with linked ECG-TTE studies from 2022-2023, who were *not* included in the development set, was used as a *temporally distinct test cohort*. Among these, 5,198 individuals had their first known TTE within 90 days *following* an ECG. The *geographically distinct test cohorts* from the UK Biobank included 33,518 individuals who underwent protocolized ECG and CMR imaging, whereas from the MIMIC-IV EHR dataset we identified 5,231 adult individuals with paired ECG and TTE reports within 90 days of each other (see **Table 1**).

**Table 1:** Study Population Characteristics.

| Participant Characteristics |  | YNHHS |  |  |  |
| --- | --- | --- | --- | --- | --- |
|  |  | Development | Temporal testing | UK Biobank | MIMIC-IV |
| **Year of Visit** |  | 2013 to 2021 | 2022 or 2023 | 2014-2020 | 2008-2020 |
| **Number of ECGs** |  | 754,533 | 8,979 | 33,518 | 5,231 |
| **Number of unique participants** |  | 159,322 | 8,979 | 33,518 | 5,231 |
| **Age at ECG (years)** |  | 68 [57-78] | 68 [58-78] | 65 [58-70] | 68 [56-79] |
| **Sex** |  |  |  |  |  |
|  | Female | 80,275 (50.4) | 4,521 (50.4) | 17,463 (52.1) | 2,348 (55.1) |
|  | Male | 79,034 (49.6) | 4,458 (49.6) | 16,055 (47.9) | 2,883 (44.9) |
|  | Not specified/Missing | 13 (0.0) | 0 (0.0) | 0 (0.0) | 0 (0.0) |
| **Race** |  |  |  |  |  |
|  | American Indian or Alaska Native | 479 (0.3) | 28 (0.3) | 0 (0) | 14 (0.3) |
|  | Asian | 2,764 (1.7) | 183 (2.0) | 445 (1.3) | 148 (2.8) |
|  | Black or African American | 20,531 (12.9) | 1,161 (12.9) | 219 (0.7) | 613 (11.7) |
|  | Native Hawaiian or Other Pacific Islander | 313 (0.2) | 18 (0.2) | 0 (0) | 8 (0.2) |
|  | White | 121,260 (76.1) | 6,723 (74.9) | 32,430 (96.8) | 3,708 (70.9) |
|  | Other/Unknown | 13,975 (8.8) | 866 (9.6) | 424 (1.3) | 740 (14.1) |
| **Ethnicity** |  |  |  |  |  |
|  | Hispanic | 14,034 (8.8) | 902 (10.0) | n/a | 220 (4.2) |
|  | Non-Hispanic | 140,977 (88.7) | 7,801 (86.9) | n/a | 4,675 (89.4) |
|  | Unknown | 4,311 (2.5) | 276 (3.1) | n/a | 336 (6.4) |
| **Extractable EHR phenotypes\*** |  |  |  |  |  |
|  | Hypertension | 117,398 (73.7) | 6,485 (72.2) | 6,331 (14.8) | 3,445 (65.9) |
|  | Diabetes mellitus | 42,735 (26.8) | 2,272 (25.3) | 1,289 (3.0) | 1,035 (19.8) |
|  | Hypercholesterolemia | 34,930 (21.9) | 1,917 (21.4) | 2,779 (6.5) | 936 (17.9) |
|  | Chronic cardiac disease | 73,225 (46.0) | 3,243 (36.1) | 2,457 (5.8) | 3,655 (69.9) |
|  | Heart failure | 37,710 (23.7) | 1,689 (18.8) | 228 (0.5) | 2,458 (47.0) |
|  | Stroke | 33,275 (20.9) | 1,646 (18.3) | 585 (1.4) | 2,458 (18.8) |
|  | Chronic kidney disease | 15,886 (10.0) | 551 (6.1) | 127 (0.3) | 138 (2.6) |
|  | Chronic respiratory disease | 55,904 (35.1) | 2,801 (31.2) | 1,069 (2.5) | 1,918 (36.7) |
|  | Cancer | 20,159 (12.7) | 1,126 (12.5) | 2,991 (7.0) | 886 (16.9) |

Abbreviations: ECG: electrocardiography; EHR: electronic health record; MIMIC: Medical Information Mart for Intensive Care; YNHHS: Yale-New Haven Health System. Data are presented as counts (percentages) or median [25<sup>th</sup>-75<sup>th</sup> percentile].
\* These are EHR phenotypes extracted from administrative codes, and do not include self-reported diagnoses or conditions.

### Learning EHR and ECG Image Representations of SHD

Our ECG image (vision)-TTE (language) contrastive model was trained for a total of 100 epochs, with validation loss reaching a plateau (**Fig. S2**). In the validation set, the trained encoder ranked the correct TTE report at the 85.6^th^ percentile [IQR: 64.6–96.1] for its corresponding ECG image. During temporally distinct testing (n=8,979), zero-shot classification of new cases based on the reference centroids of SHD cases and controls from the training set, successfully discriminated 26 SHD labels with a median AUROC of 0.78 [25^th^-75^th^ percentile: 0.76-0.80] and range of 0.71 to 0.90 (**Fig. 3a**), including left ventricular systolic dysfunction (LVSD) with an AUROC of 0.90, severe aortic stenosis (AS) with an AUROC of 0.85, elevated right ventricular systolic pressure (RVSP ≥50 mm Hg) with an AUROC of 0.82, and mitral stenosis with AUROC of 0.75. For reference, similar comparison of the associated EHR CLMBR-T embeddings at the time the ECG was recorded yielded a median AUROC of 0.62 [25^th^-75^th^ perc.: 0.60-0.66]. Importantly, these metrics were comparable in an ablation study that removed sex, race and ethnicity from the EHR embeddings (median AUROC of 0.61 [25^th^-75^th^ perc.: 0.69-0.65]) (**Fig. 3b**). Finally, we confirmed that the ECG ViT model and derived embeddings preserved their foundational properties when deployed in the released subset of the EchoNext dataset (v1.1.0),^27–29^ maintaining excellent performance even when selecting as few as 10 reference examples for a range of 12 SHD labels (from a released training set of 72,475 ECGs from 26,218 unique patients). In the testing cohort of EchoNext (n = 5,442 ECGs from 5,442 unique participants; median age 64 [52–74] years; 2,731 [50.2%] female), the model achieved AUROCs of 0.88 for LVEF≤45%, 0.82 for moderate-or-greater AS, and 0.82 for a composite of 11 major SHD subtypes (**Fig. S3**).

**Fig. 3:**
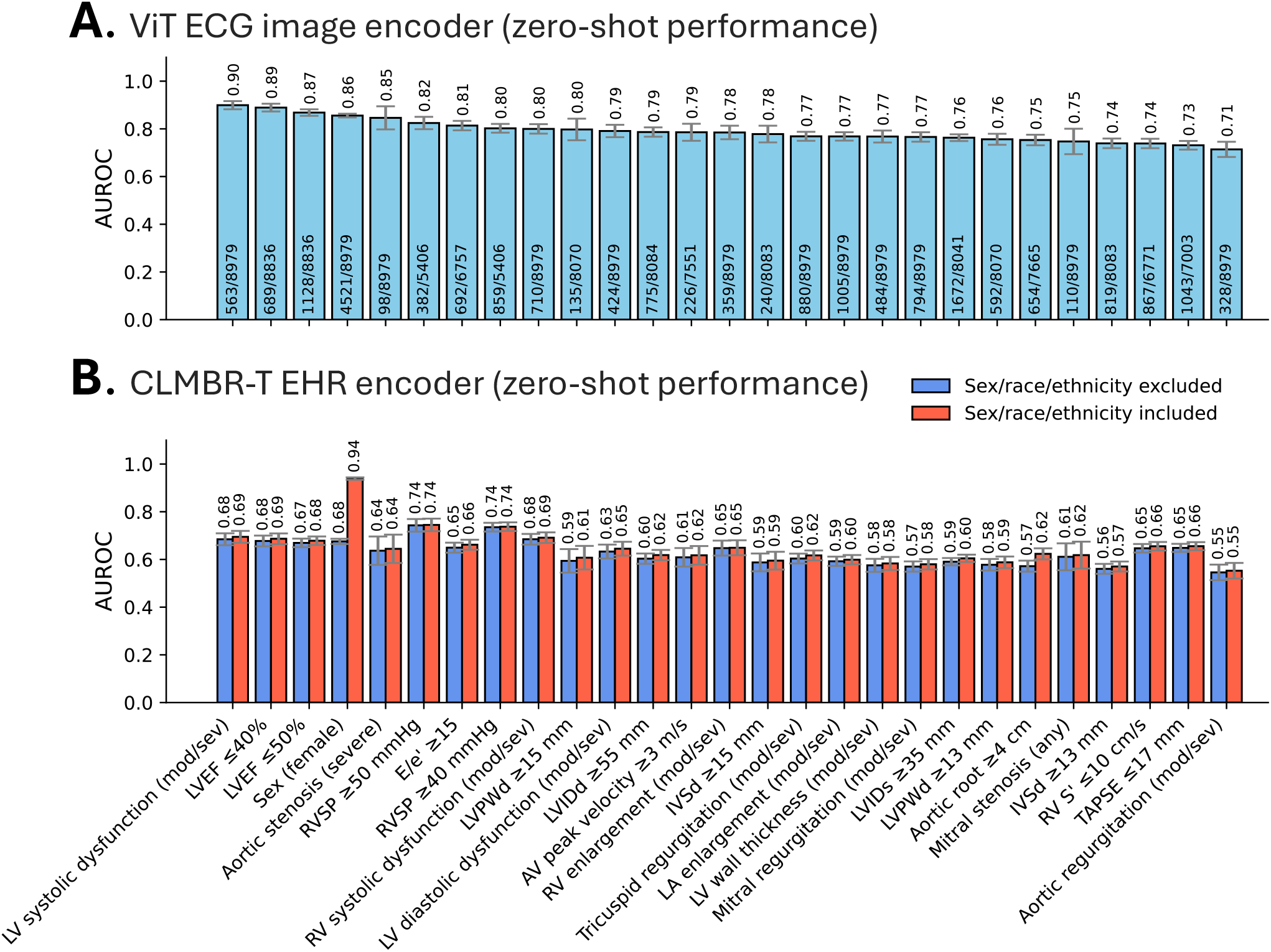
Zero-shot evaluation of AI-ECG ViT and CLMBR-T across the spectrum of echocardiographic phenotypes. During zero shot evaluation, we use the ECG Image ViT and EHR CLMBR-T to project each new case to the respective embeddings. For each SHD label (ascertained based on an echocardiogram performed within 90 days), we calculate the absolute difference in the cosine similarity to the centroid of all cases in the training set versus the centroid of all controls. This relative similarity is then used to rank the test set observations for a given label and compute discrimination metrics (AUROC), which are then visualized using bar plots for the ViT ECG Image **(A)** and CLMBR-T encoders **(B)**, respectively. We present two versions of CLMBR-T metrics, both with (red) and without (blue) protected demographics (sex, race, ethnicity). Error bars denote 95% confidence intervals. *AI: artificial intelligence; AUROC: area under the receiver operating characteristic curve; AV: aortic valve; E/e’: ratio of early mitral inflow velocity to early diastolic mitral annular velocity; ECG: electrocardiography; IVSd: interventricular septum thickness at end-diastole; LA: left atrium; LV: left ventricle; LVEF: left ventricular ejection fraction; LVIDd: left ventricular internal diameter at end-diastole; LVIDs: left ventricular internal diameter at end-systole; LVPWd: left ventricular posterior wall thickness at end-diastole; RA: right atrium; RV: right ventricle; RVSP: right ventricular systolic pressure; SHD: structural heart disease; TAPSE: tricuspid annular plane systolic excursion; ViT: vision transformer*.

### Joint Modeling of EHR and ECG Image Embeddings for Targeted AI-ECG Deployment

A representation of EHR and ECG phenotypes across the YNHHS testing population is shown in **Fig. S4**. We found that, compared with the *untargeted* approach, a *targeted* sequential strategy yielded significantly higher F1 scores (values range from 0 to 1) across all 26 SHD labels, with a median absolute F1 increase of 0.25 (range: 0.09 to 0.75) (**Fig. 4**), driven by a 0.48 (range: 0.19 to 0.93) median absolute increase in precision (PPV), and a marginal absolute change in recall (sensitivity) of 0.07 (range: –0.12 to 0.58). These findings translated into a median change (reduction) of –303 (range: –715 to –77) in false positive counts, with only marginal changes in false negatives (median change of –18 [range: –99 to 68]), and were robust to a sensitivity analysis where the untargeted approach used concatenated EHR and ECG embeddings (see comprehensive results in **Table S4**).

**Fig. 4:**
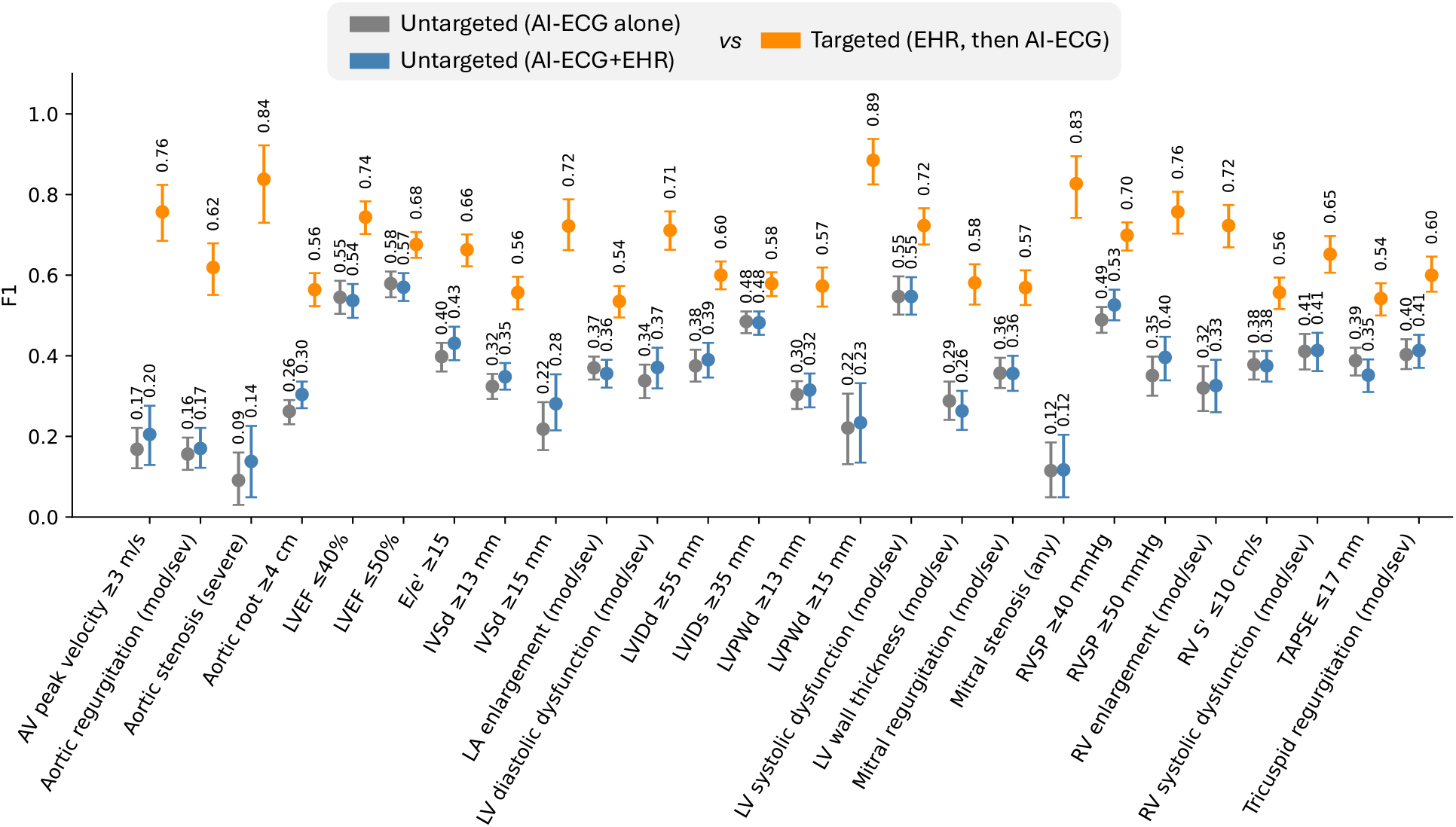
Comparative performance of untargeted and targeted AI-ECG strategies for detection of SHD. We compared three approaches for detecting SHD phenotypes defined by transthoracic echocardiography: an untargeted AI-ECG-alone strategy (gray), an untargeted AI-ECG + EHR strategy (blue), and a targeted EHR-guided approach (orange), in which finetuned EHR embedding models (CLMBR-T) identify individuals for subsequent AI-ECG evaluation. For each label and strategy, we present the point F1-score estimate (dot) accompanied by error bars summarizing the 95% confidence intervals calculated with bootstrapping (500 replications). *AI: artificial intelligence; AUROC: area under the receiver operating characteristic curve; AV: aortic valve; ECG: electrocardiogram; E/e′: ratio of early mitral inflow velocity to early diastolic mitral annular velocity; IVSd: interventricular septal thickness at end-diastole; LA: left atrium; LV: left ventricle; LVEF: left ventricular ejection fraction; LVIDd: left ventricular internal diameter at end-diastole; LVIDs: left ventricular internal diameter at end-systole; LVPWd: left ventricular posterior wall thickness at end-diastole; MR: mitral regurgitation; MS: mitral stenosis; RV: right ventricle; RVSP: right ventricular systolic pressure; SHD: structural heart disease; TAPSE: tricuspid annular plane systolic excursion; TR: tricuspid regurgitation; ViT: vision transformer*.

### External Validation

Upon external deployment in the UK Biobank (**Fig. 5a**), we first confirmed the out-of-the-box ability of the ECG ViT to classify structural abnormalities by CMR, including LVSD (AUROC of 0.85), increased left ventricular thickness (AUROC of 0.85), and left atrial enlargement (AUROC 0.88) based on the original YNHHS reference embeddings (**Fig. S5**). Across 7 SHD labels, we confirmed a median absolute increase in F1 score of 0.20 (range: 0.05-0.33) with targeted versus untargeted AI-ECG deployment, corresponding to a median change (reduction) in false positives of –1,104 (range: –9,991 to –60) (**Fig. S6**). Similarly, in MIMIC-IV, a targeted framework informed by historical data in the same hospital system (median anchor dates of 2011 or earlier, n=1,603) and deployed in temporally distinct encounters from different patients (2012 or later, n=3,628) resulted in a median absolute improvement in F1 scores of 0.13 (range: 0.08 to 0.16) with a median absolute change (decrease) of –255 (range: –716 to –86) in false positives across 5 SHD labels (**Fig. S7**). On both occasions, results were consistent when comparing the targeted approach against an untargeted approach relying on concatenated EHR and ECG image embeddings (**Fig. S6-7**), with significant reductions in F1 and false positive counts seen across all labels and comparisons.

**Fig. 5:**
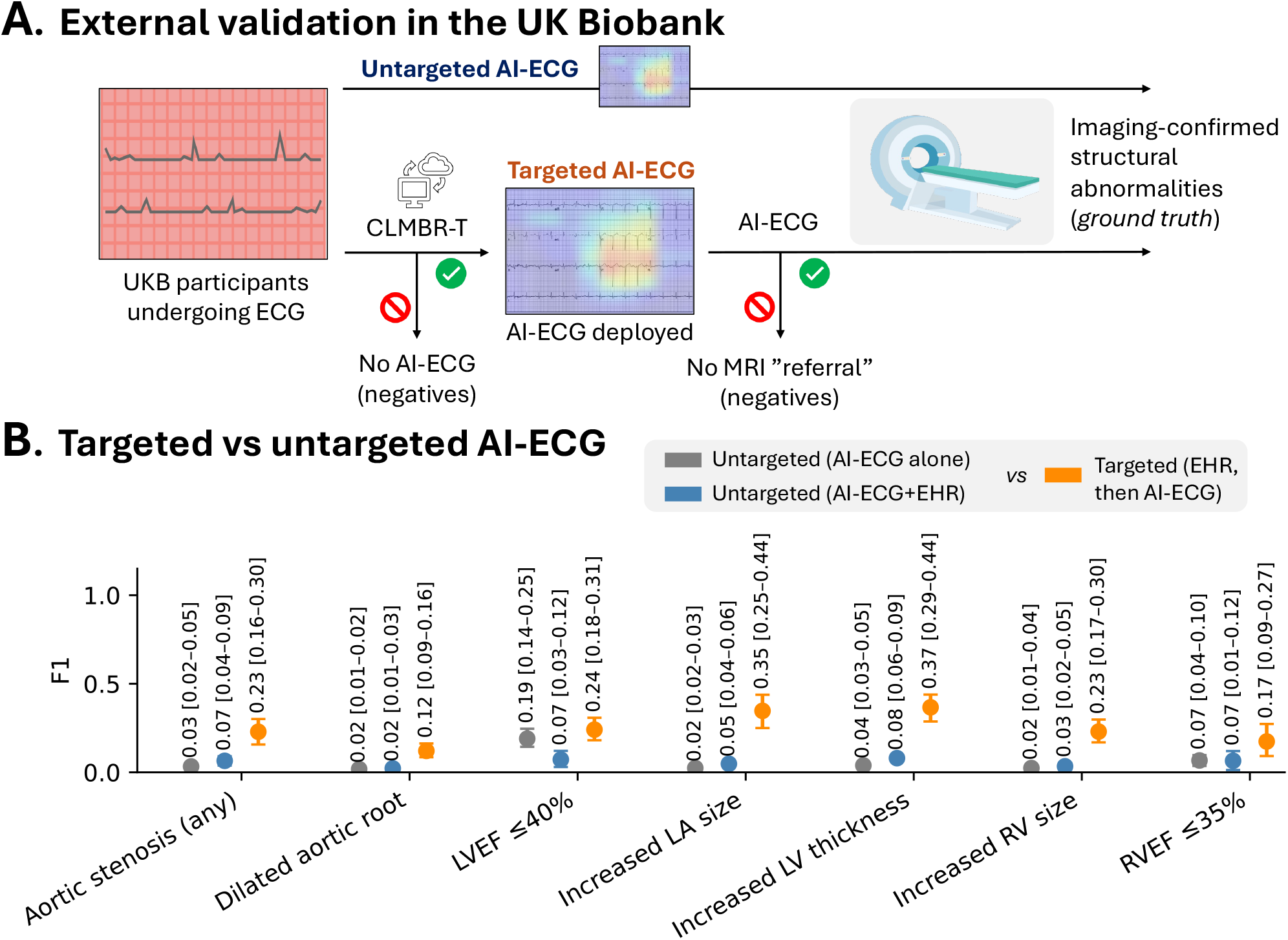
External validation in the UK Biobank. **(A)** External deployment of the targeted AI-ECG approach in the UK Biobank among 33,518 UK Biobank participants undergoing concurrent ECG and cardiac MRI imaging. We applied the pre-trained encoders and label-specific supervised headers in parallel for both the untargeted and the targeted approach, with SHD defined based by CMR. **(B)** Summary of F1-score (including 95% confidence intervals derived from bootstrapping with 500 replications) for the targeted vs untargeted AI-ECG screening strategies across eligible SHD labels. *AI: artificial intelligence; ECG: electrocardiography; MRI: magnetic resonance imaging; SHD: structural heart disease*.

## DISCUSSION

This work introduces two key innovations for the scalable and clinically meaningful deployment of AI-ECG. First, we develop and release an ECG image-based foundation model, by leveraging contrastive pretraining at scale across linked echocardiographic reports to learn generalizable representations spanning the full spectrum of SHD phenotypes. Second, we integrate this vision model with a longitudinal EHR foundation model, jointly modeling spot ECG-derived embeddings and dynamic patient trajectories to inform a targeted AI-ECG screening paradigm. This multimodal approach enables greater precision in the identification of individuals most likely to benefit from AI-ECG evaluation, potentially reducing unnecessary deployment and improving diagnostic yield. Across temporally and geographically distinct cohorts, this framework improves the balance of precision and recall across diverse SHD phenotypes, while substantially reducing false positives and testing burden. Together, these findings define a foundational, interoperable strategy for deploying AI-ECG in real-world care pathways, maximizing clinical utility with potential unintended downstream costs.

As the number of AI-ECG models continues to rise exponentially,^1^ little guidance exists on how to implement these tools effectively within real-world clinical care pathways.^31^ The controlled environment of retrospective case-control validation studies where most models are developed and tested does not replicate the multimodal reasoning that underpins clinical decision-making, which draws on both recent diagnostics and longitudinal patient history.^2^ In practice, test interpretation is inherently contextual, influenced by an individual’s trajectory, comorbidities, and prior risk assessments, factors that are often missing from AI deployment pipelines. To address this gap, our proposed solution combines two complementary foundation models: an ECG image-based vision transformer, designed for direct and flexible use with raw ECG images (e.g., screenshots or smartphone captures),^18–21^ and a pre-trained CLMBR-T transformer that encodes longitudinal EHR trajectories using standardized, interoperable data schemas.^13,14^ This enables a dynamic, data-driven gating system that selects candidates for AI-ECG screening based on their evolving clinical context in the EHR. Importantly, we show that neither modality alone is sufficient: EHR embeddings lack the phenotypic specificity captured by ECG signals, while ECG-only models may lack awareness on key features that best define an individual’s pre-test probability and thus the optimal interpretation of a positive versus negative screen. Instead, we find that their joint integration allows for high-fidelity, high-precision screening. Given an estimated cost of ∼$125 per AI-ECG-enabled interpretation per the 2025 Centers for Medicare & Medicaid Services, this potential reduction in low-yield model deployment has implications for scalability and cost-effectiveness across health systems.

From a clinical standpoint, the utility of TARGET-AI extends beyond improved model discrimination; it provides a tunable, resource-aware framework for deploying AI-ECG screening safely and efficiently. Across validation cohorts, the targeted sequential strategy improved positive predictive value and overall F1 performance without consistently compromising recall. Because both the EHR gating and ECG thresholds can be dynamically adjusted, the framework can be tailored to specific diseases and resource environments, such as by emphasizing sensitivity in high-consequence phenotypes or precision in low-prevalence conditions. This adaptability may potentially enable context-specific optimization of screening strategies, supporting responsible and scalable implementation of AI-enabled cardiac diagnostics within diverse health systems. Importantly, the goal of TARGET-AI is not to limit ECG acquisition or to discourage the use of AI itself, but to optimize how AI is applied. By coupling AI-ECG with EHR-derived phenotypes, TARGET-AI may enable more precise, resource-aware deployment, ensuring that AI is used where it provides the greatest incremental value rather than simply greater volume of testing.

Our approach also offers methodological advances. First, it introduces the first standalone AI-ECG foundation model for SHD screening using ECG images, the most accessible, interoperable, and clinically ubiquitous format, enabling scalable diagnostic deployment across care settings, including remote and resource-limited environments. Second, it establishes and validates the novel concept that longitudinal EHR embeddings can function as interoperable gatekeepers for AI deployment, identifying individuals most likely to benefit from AI diagnostics. Given its foundational nature, the system also supports dynamic retuning to accommodate new disease labels and evolving practice patterns,^32^ as well as locally optimal adaptation across distinct health systems. Importantly, the EHR-derived risk enrichment operates largely independently of protected demographic variables such as sex, race, or ethnicity.^33,34^ Third, the framework is built with a high degree of interoperability across both ECG and EHR domains, covering a broad spectrum of SHD phenotypes and supporting integration into heterogeneous health systems. Fourth, by open-sourcing the model weights and codebase, we empower both individual investigators and institutions to fine-tune and adapt the system to their unique populations, accelerating innovation and deployment. Finally, although EHR embeddings reduce interpretability, they provide unified patient representations reusable across disease labels, capturing longitudinal trajectories and informative missingness.

## LIMITATIONS

Certain limitations merit consideration. First, the EHR-derived embeddings capture longitudinal trajectories of structured phenotypes but do not incorporate unstructured information such as encounter notes. Second, while the UK Biobank provides valuable external validation, its protocolized, population-based design differs fundamentally from clinical care, since ECGs and CMR scans were protocolized. Similarly, the analysis in the YNHHS and MIMIC-IV EHR demonstrates how the approach can be locally adapted to heterogeneous environments, illustrating both its flexibility and the importance of local calibration for precision deployment. Third, formal confirmation of operational sufficiency will require prospective evaluation in real-world clinical workflows to account for local implementation constraints, disease prevalence and resource variability. Fourth, we acknowledge that the ECG ViT model may not generalize to less typical ECG formats beyond the ones previously described and seen during training.^18^ Fifth, we did not perform direct comparisons against expert reads, as such annotations were inconsistently available and conventional readings focus on visible rather than hidden labels. Sixth, differences between strategies may reflect both threshold optimization and underlying population enrichment, as the targeted framework is applied within an EHR-selected cohort that differs systematically from the full population. Finally, prospective studies should explore the deployment of this targeted strategy across the EHR to examine its validity and robustness to potential data drifts or shifts.

## CONCLUSION

We introduce TARGET-AI, a foundational framework for the targeted deployment of AI-enabled screening technologies within the EHR. By selectively enriching the population through multimodal integration, this approach provides a flexible and interoperable paradigm for optimizing AI-ECG and other AI-assisted tools, which may be locally adapted across health systems and computable disease labels with appropriate validation.

## DATA AVAILABILITY

We have released code for the training and inference steps of our manuscript, including a sample (demo) pipeline for inference and embedding extraction from new ECG images, as well reference embeddings for the centroids of cases and controls across 26 representative echocardiographic labels from the YNHHS training set, and the 12 (11 individual, 1 composite) SHD labels from the public release of the EchoNext dataset: https://github.com/CarDS-Yale/target-ai-shared. Model weights for the ECG image ViT are accessible from: https://huggingface.co/CarDSLab/ecg-clip-beit-base-384. The original CLMBR-T model may be accessed from: https://huggingface.co/StanfordShahLab/clmbr-t-base.

