## Supplementary material for "*TARGET-AI*: a foundational approach for the targeted deployment of artificial intelligence electrocardiography in the electronic health record": Online Supplement

### *Supplementary Appendix*

Evangelos K. Oikonomou, MD, DPhil[a,b*], Bruno Batinica, MBBS[a,b], Lovedeep S. Dhingra, MBBS[a,b], Arya Aminorroaya, MD, MPH[a,b], Andreas Coppi, PhD,[b,c] Rohan Khera, MD, MS[a,b,c,d,e*]

a. Section of Cardiovascular Medicine, Department of Internal Medicine, Yale School of Medicine, New Haven, CT, USA.
b. Cardiovascular Data Science (CarDS) Lab, Yale School of Medicine, New Haven, CT, USA.
c. Center for Outcomes Research and Evaluation, Yale-New Haven Hospital, New Haven, CT, USA.
d. Section of Biomedical Informatics and Data Science, Yale School of Medicine, New Haven, CT, USA.
e. Section of Health Informatics, Department of Biostatistics, Yale School of Public Health, New Haven, CT, USA.

**Table of contents:**
***Pages 2-6**:* Supplemental Methods
***Pages 7-13**:* Supplemental Figures S1-S7
***Pages 14-24**:* Supplemental Tables S1-S4
***Pages 25-26***: Supplemental References

***Correspondence:**
*Evangelos K. Oikonomou, MD, DPhil*
195 Church St, 6th Floor, New Haven, CT 06510
; @ekoikonomou

*Rohan Khera, MD, MS*
195 Church St, 6th Floor, New Haven, CT 06510
; @rohan_khera

### SUPPLEMENTAL METHODS (Extended Version)

#### Data Source and Population

The study was approved by the Yale Institutional Review Board (IRB) which waived the need for informed consent as a retrospective analysis of previously collected data. The UK Biobank analysis was conducted under the approved project #71033.

***Yale-New Haven Health System (YNHHS)***: YNHHS is a large, integrated system of five hospitals and several outpatient ambulatory clinical locations across Connecticut and Rhode Island. We queried the institutional electronic health record (EHR) since its inception (from 2013 until November 28, 2023) to identify all individuals who underwent a 12-lead electrocardiogram (ECG) and standard transthoracic echocardiography (TTE) within 90 days of each other (-90 to +90 days). Eligible individuals were 18 years or older at the time of their study and had not previously opted out from research studies. For all eligible individuals, we extracted structured records from the EHR spanning diagnosis codes (both inpatient and outpatient), medications, procedures, laboratory measurements and results, codes and health system encounters. ECGs were extracted as signals and randomly plotted as images across various lead layouts, as per our previously described protocol and work,[1–4] whereas the summary echocardiographic report was extracted in text format. A total of 754,533 ECG-TTE pairs from 159,322 unique individuals from 2013 until the end of 2021 were used for model development, while capping the maximal number of ECGs per individual at 10 to prevent over-representation of phenotypes more likely to interact with the health system. A total of 8,979 unique participants from 2022 and 2023 who were not seen during model development were kept as a temporally distinct test set, with one random pair of ECG-TTE retrieved for each patient (8,979 ECG-TTE pairs in total). When modeling the targeted vs untargeted deployment of AI-ECG testing, we further restricted the analysis to those individuals who had their first ever TTE in our system within a maximum of 90 days *after* their ECG (n=5,198), thus ensuring that EHR codes did not reflect previously known echocardiographic phenotypes.

***UK Biobank***: In the UK Biobank, we identified a total of 33,518 individuals who underwent 12-lead ECG and cardiac magnetic resonance (CMR) imaging as part of a standard protocolized assessment and had available CMR measurements. Briefly, the UK Biobank is a prospective observational study of 502,468 participants aged 40-69 years recruited between 2006 and 2010 which continues to collect extensive phenotypic and genotypic details using multimodal data capture.[5] This geographically distinct and protocolized context enabled a robust framework to evaluate the external validity of our approach. To extract longitudinal health records, we relied on structured fields describing demographics (f34/f32: year/month of birth set to the first day of the calendar month; f31: gender; f21000: ethnicity), medications (f29039), hospital diagnosis codes (f41270 and f41280), whereas laboratory measurements (e.g., f30000 for leukocyte count) were derived from protocolized assessments embedded in the study protocol across sequential assessment visits (0, 1, 2; excluding any visits done after the index study where ECG and CMR were performed).

***MIMIC-IV***: To evaluate the generalizability of the TARGET-AI framework to an independent health system and its EHR, we used deidentified records from Medical Information Mart for Intensive Care (MIMIC)-IV, a large dataset of patients admitted to the emergency department or an intensive care unit at the Beth Israel Deaconess Medical Center in Boston, MA. Here, we queried the available discharge summaries to retrieve embedded diagnostic reports of transthoracic echocardiograms, which were then extracted and pre-processed through a validated model (HeartDx-LLM)[6] to allow transformation into structured columns. We then identified ECGs (with available signals) which were performed within 90 days of the index admission (defined based on the date of the discharge summary) to create pairs of temporally linked ECGs and TTEs. Replicating the framework implemented across YNHHS, we leveraged EHR-derived fields across diagnosis codes, laboratory measurements, medications, procedures, and key demographics to define longitudinal representations, ranging from the inception of the local EHR up until the time the ECG was performed. Given the deidentified nature of the dataset, we estimated index dates using the provided anchor dates and using their median values (e.g. 2013 for anchor period: 2012-2014), before applying the same time difference across all relevant dates and events.

**Data Preprocessing & Label Definition**

***Yale-New Haven Health System (YNHHS)***: All 12-lead ECG studies were extracted and pre-processed to create ECG images across four different layouts (standard, alternate, standard shuffled and alternate shuffled), as described and validated in our previous work.[1–4] This strategy addresses the need for interoperability and boosts the accessibility of such algorithms to the end-users who generally lack access to vendor-specific ECG signals. TTE reports were extracted from the local EHR and echocardiography reporting software and linked database (Lumedx, Oakland, CA). In addition to textual reports that represent distilled summaries of the main findings for key structures and parameters, we also extracted structured labels, as per our previous work.[7] These served as ground truth labels for the presence of SHD during the inference and evaluation stage. A summary and linked definitions of the 27 representative labels included in the YNHHS analysis is presented in **Table S1**.

***UK Biobank***: In the UK Biobank analysis, cardiac magnetic resonance (CMR) imaging measurements were derived from automated measurements, representing automated measurements as per the method described by Bai et al (f24103 for left ventricular ejection fraction with a threshold of ≤40%; f24100: left ventricular end-diastolic volume index; maximum of f24124 through f24139 as the maximal left ventricular thickness; f24109 for right ventricular ejection fraction with a threshold of ≤35%; f24106 for right ventricular end-diastolic volume index; f24110 for left atrial volume index; and f24118 for ascending aorta diameter).[8] Given differences in the measurement techniques across modalities, for thickness, linear and volumetric measurements on CMR we defined significant abnormalities using population-specific thresholds reflecting ≥99.5$^{th}$ percentile. This resulted in the reliable definition of 7 echocardiographic abnormalities: aortic stenosis (defined based on any ICD-10 code, e.g. I35-; with severity unspecified), as well as dilated aortic root, left ventricular systolic dysfunction (LVEF ≤40%), left atrial enlargement, left ventricular hypertrophy, right ventricular enlargement or systolic dysfunction (RVEF ≤35%).

***MIMIC-IV***: Using the structured reports generated by the HeartDx-LLM[6] model we defined 5 key echocardiographic abnormalities, namely left ventricular ejection fraction ≤40%, ≤50%, any grade of diastolic dysfunction, left ventricular hypertrophy (moderate-or-severe), and any moderate-or-severe left-sided valvular abnormality (composite of aortic stenosis, aortic regurgitation, mitral stenosis or mitral regurgitation). Since these estimates were derived from summary reports containing the impression and conclusions of the echocardiograms, we assumed that any condition not explicitly mentioned was absent (e.g., if no moderate or severe aortic stenosis was mentioned, we assumed that there was no moderate-or-severe AS present).

**CLMBR-T model embeddings**

CLMBR-T (clinical language modeling based representations-transformer) is a 141 million parameter autoregressive foundation model pretrained on 2.57 million deidentified EHRs from Stanford Medicine.[9,10] The input for the model is a sequence of coded medical events mapped along the OMOP-CDM vocabulary and standard concepts, with each individual presented based on the Medical Event Data Standard (MEDS) schema. This transformer-based architecture trained to predict the next code/token in a patient's EHR trajectory based on previous codes enables flexible and interoperable definition of longitudinal phenotypes that is robust to sparse embeddings and missing data. In **Table S2**, we present a detailed summary of the mapping process across domains and fields in the YNHHS, UK Biobank and MIMIC-IV cohorts. In summary, we successfully mapped 26,828 [96.0%] of 27,946 non-laboratory-related codes across YNHHS. To facilitate the mapping process, we relied on external dictionary files from the Unified Medical Language System (UMLS) to map ICD-10 (international classification of diseases - 10th edition) codes to SNOMED CT (Systematized Nomenclature of Medicine - Clinical Terms) concepts and medication names to RxNorm concept unique identifiers (RXCUI). For summarization purposes, we pooled related SNOMED CT codes into grouped concepts (i.e., hypertension, diabetes mellitus) by referring to open repositories of code groupings (see **Table S3**). For the remaining codes (e.g., Logical Observation Identifiers Names and Codes [LOINC]-related), we curated a list of 46 commonly performed tests, spanning complete blood count, comprehensive metabolic panel, thyroid function, brain natriuretic peptides, troponin, glycated hemoglobin a1c and standard lipid panel measurements. At the time of each ECG, we built a MEDS (Medical Event Data Standard)-compatible schema for each patient and excluded any events recorded after the timepoint of the ECG. This was processed through CLMBR-T, and the last 768-dimensional hidden layer was extracted as a reference embedding reflecting an individual's longitudinal trajectory at the time an ECG was performed.

**Contrastive ECG-TTE vision-text model training**

We used a CLIP-like contrastive learning paradigm to align ECG images with corresponding textual ECG reports, leveraging a dual-encoder architecture: the vision encoder (ViT) used a BEiT-base-patch 16-384 backbone configured for 384x384-pixel inputs split into 16×16 patches,[11] along with 12 hidden layers, 12 self-attention heads, and the default hidden dimension of 768 projected to a 512-dimensional embedding. The text encoder followed the standard CLIP transformer design, adapted to a maximum token length of 256, featuring 12 hidden layers, 8 attention heads, and a hidden dimension of 512. Both encoders projected embeddings into a shared 512-dimensional latent space. Given the repetition and limited vocabulary of TTE reports, we trained a custom Byte-Pair Encoding (BPE) tokenizer on the training TTE reports, with the maximal vocabulary size set at 20,000. A frequency filter required a token to be present ≥3 times to be considered for further inclusion, limiting the number of tokens to 16,030. We split ECG reports at the patient level, allocating 95% of unique participants for training and 5% for validation, while ensuring that no patient overlapped across sets and restricting each patient to a maximum of 10 ECGs. We further applied standard data augmentation by including random rotation (±5° degree rotations), whereas each image was randomly plotted using one of the four different layouts, in line with our prior work.[1–4]

We trained the contrastive model for a total of 100 epochs using the AdamW optimizer, with a $10^{-5}$ learning rate, linear warm-up and a cosine decay schedule. We employed mixed-precision training to reduce GPU memory usage, using a batch size of 64 and gradient accumulation steps set to 1. The loss function was the symmetric cross-entropy objective, as introduced in CLIP,[12] computed by averaging cross-entropy between (i) images and their matching text indices and (ii) texts and their corresponding image indices. In the validation set, we tracked performance based on a retrieval-based paradigm. Here, for each ECG image, we evaluated its cosine similarity against all text embeddings in the validation set, determining the percentile rank of the true matching report. The median rank percentile (with IQR) served as the primary performance metric, reflecting how frequently the correct pair outranked random mismatches (**Fig. S2**). This plot was used for qualitative evaluation purposes and as a check that the model learned reproducible associations. However, we did not set an early stopping function, rather training our model for the total duration of 100 epochs, as done in other similar approaches.[13] Training was done on 8 NVIDIA H100 graphics processing units (GPUs).

**Zero-shot classification**

During the zero-shot evaluation process in the testing set of the YNHHS cohort, for both the AI-ECG ViT and EHR CLMBR-T models and for each SHD label, we split the training data based on whether each observation was positive or negative for that label (i.e., left ventricular systolic dysfunction, aortic stenosis). The image embeddings from each positive group were averaged to produce two centroids, capturing the prototypical embedding of the positive and negative classes. For unseen data in the temporally distinct test set from 2022-2023, embeddings were directly computed by feeding the index EHR recordings and ECG images through the ViT and CLMBR-T models. We then computed the cosine similarity between each testing set embedding and the two reference centroids from the training set, thus producing a single classification score, defined as the difference in similarity to the positive versus negative centroids. We used this score to estimate the area under the receiver operating characteristic curve (AUROC) separately for each label. For CLMBR-T embeddings, we further performed an ablation experiment masking key demographics (sex, race, ethnicity) and extracting reference embeddings without including this key information, before estimating zero-shot classification performance across labels.

**Comparative evaluation of targeted vs untargeted AI-ECG screening**

We evaluated two main strategies for AI-ECG screening: an *untargeted* approach, in which a logistic regression classifier was trained solely on ECG ViT embeddings, as specified in the Statistical Analysis section in the main manuscript, and applied to every patient, and a *targeted* (sequential) strategy, where individuals were first scored by a high-recall EHR model (CLMBR-T). In the targeted scenario, only those above a ≥90% sensitivity threshold proceeded to the ECG ViT classifier, while everyone else was automatically assigned a negative label. As part of the untargeted scenario, we also explored an integrated model that concatenated ECG ViT and CLMBR-T embeddings for direct classification. Thresholds for the ECG-based models were tuned on the training set via F1 optimization, whereas the EHR gating threshold was selected to achieve ≥90% recall. During testing, we applied each strategy and models directly to temporally distinct YNHHS (2022-2023) and geographically distinct UK Biobank cohorts. Standard classification metrics (F1 score, precision, recall, false positive rate, and other confusion-matrix derivatives) were computed to quantify performance across strategies and assess the trade-offs between missed cases, positive tests, and resource utilization.

**Supplemental Figures**

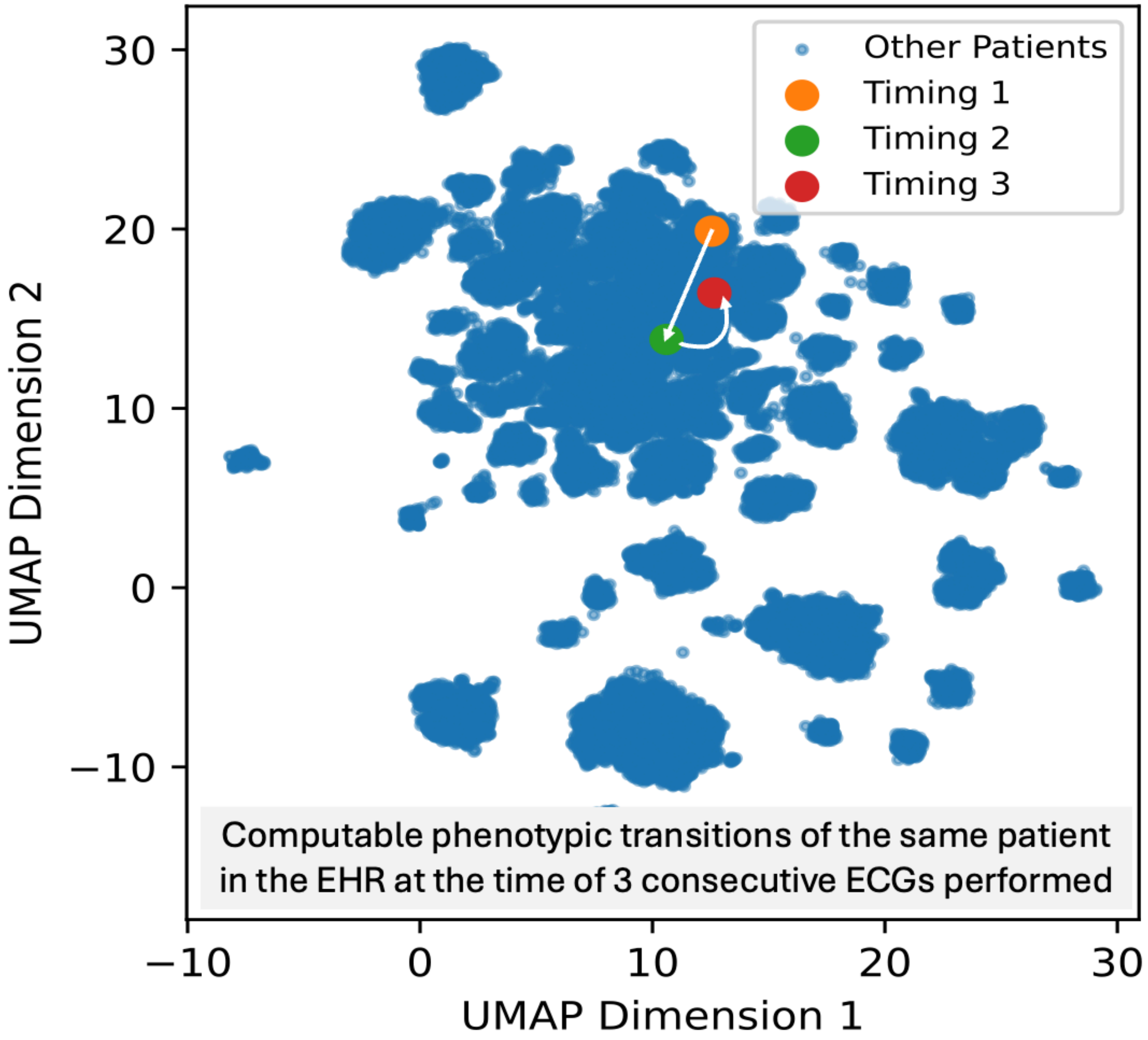


**Fig. S1 | Evolution of an individual's EHR (CLMBR-T) phenotype across sequential ECGs.** The UMAP representation reflects the longitudinal EHR phenotype based on the CLMBR-T-defined embedding at the timepoints of 3 consecutive ECGs for the same patient (performed over 12 months). The three colored dots represent different timepoints where ECG was performed in the same patient, with differences in the embedding location reflecting evolving EHR phenotypes. *CLMBR-T: clinical language model-based representation - transformer; ECG: electrocardiography; EHR: electronic health record; UMAP: Uniform Manifold Approximation and Projection.*

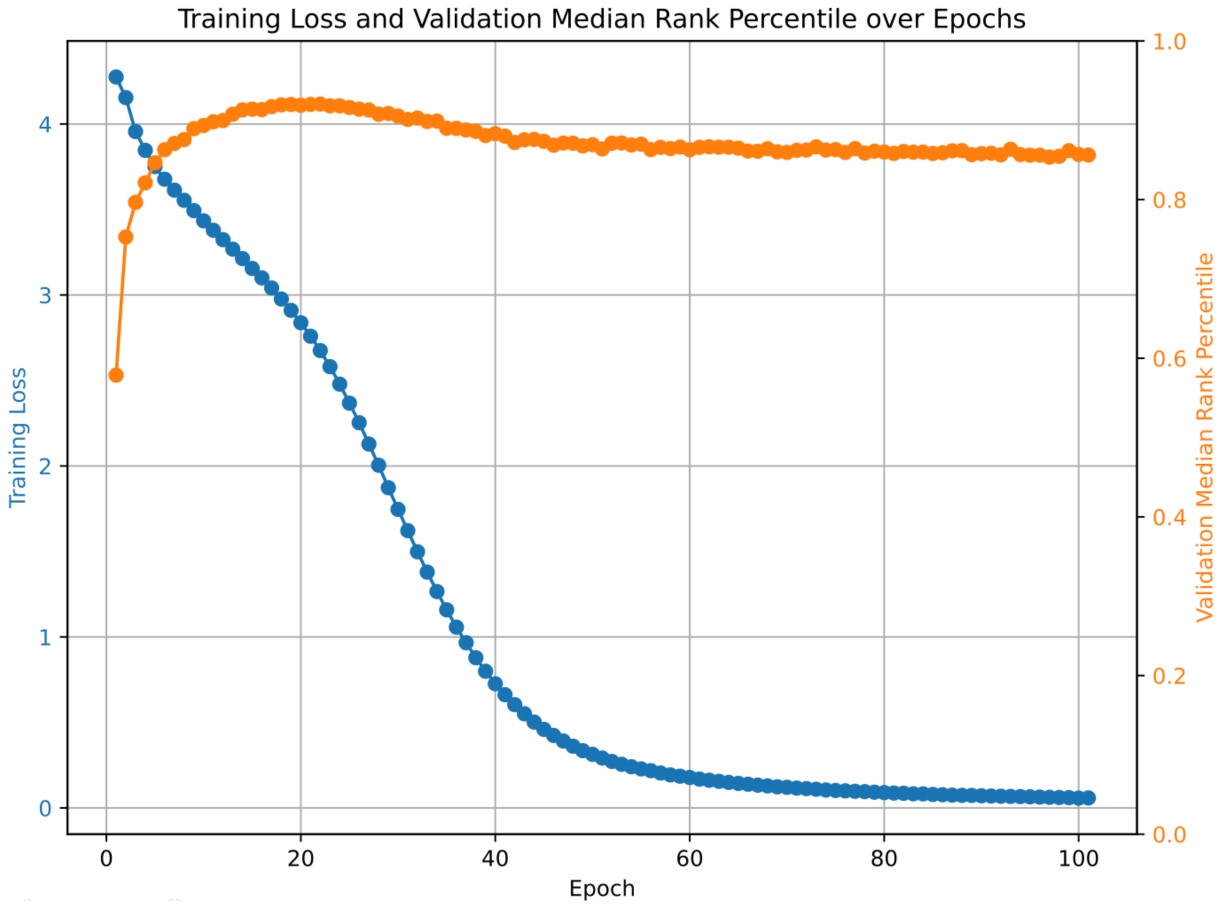


**Fig. S2 | Tracking training and validation performance for the ECG-TTE image-language model.** Tracking training loss (blue line) and validation median rank percentile for retrieving the corresponding echocardiography report based on a given input ECG image (orange line). More specifically, for each ECG image, we evaluated its cosine similarity against all text embeddings in the validation set, determining the percentile rank of the true matching report. The median rank percentile (with IQR) served as the primary performance metric, reflecting how frequently the correct pair outranked random mismatches. *ECG: electrocardiography; TTE: transthoracic echocardiogram; ViT: vision transformer.*

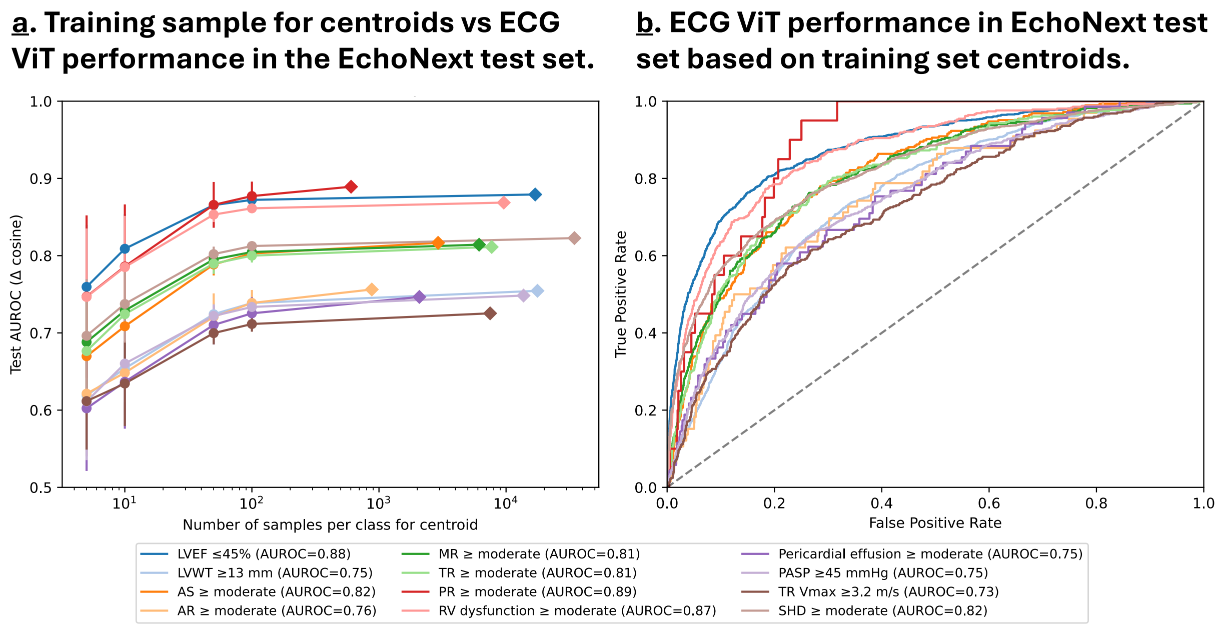


**Fig. S3 | Performance of ECG Image ViT in the EchoNext test set (v1..** (**A**) Relationship between the number of ECG image samples (5, 10, 50, 100, all examples for a given label in the provided training set) used to derive centroids for each disease phenotype and corresponding model discrimination (area under the receiver-operating-characteristic curve [AUROC]) in the independent EchoNext test set.[14] Each curve represents a distinct echocardiographic phenotype; points denote the mean AUROC across 500 bootstrap iterations, with vertical bars showing the 95% confidence intervals based on bootstrapping. Performance saturates beyond ≈ 50 samples per class, indicating that centroid estimates stabilize with modest training data. (**B**) AUROC curves for inference of representative phenotypes in the EchoNext test set, using case and control centroids computed from the full training cohort. No model retraining or fine-tuning was performed; performance reflects fixed ECG encoder representations transferred directly to the EchoNext cohort. *AR: aortic regurgitation; AS: aortic stenosis; AUROC: area under the receiver-operating-characteristic curve; ECG: electrocardiogram; LVEF: left ventricular ejection fraction; LVWT: left ventricular wall thickness; MR: mitral regurgitation; PASP: pulmonary artery systolic pressure; PR: pulmonary regurgitation; RV: right ventricle; SHD: structural heart disease; and TR: tricuspid regurgitation.*

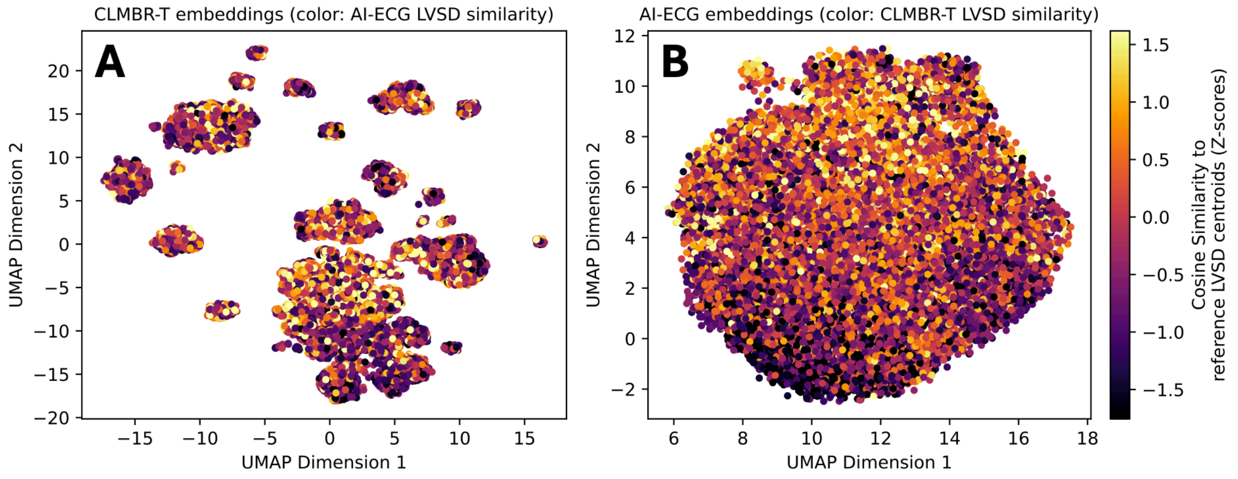


**Fig. S4 | Variation in computable phenotypes across AI-generated EHR and ECG image embeddings in the YNHHS test set.** The panels represent the low-dimensional representation of the high-dimensional space for the CLMBR-T (**A**) and AI-ECG ViT (**B**) models, respectively. The color scale denotes the similarity of each projected observation to the centroid of the cases of left ventricular systolic dysfunction in the reference training embedding. For instance, in **panel A**, the coordinates of each observation represent the low-dimensional EHR phenotype (by CLMBR-T), whereas the color map denotes the similarity or dissimilarity to the LVSD phenotype. Inversely, in **panel B**, the UMAP dimensions reflect the last hidden layer of the ECG image ViT, with the color map denoting the similarity to LVSD phenotypes based on CLMBR-T. The plots are provided for illustration purposes and serve to visualize the complementary nature of the two approaches. *AI: artificial intelligence; CLMBR: clinical language model-based representation - transformer; ECG: electrocardiography; EHR: electronic health record; UMAP: Uniform Manifold Approximation and Projection; ViT: vision transformer; YNHHS: Yale-New Haven Health System.*

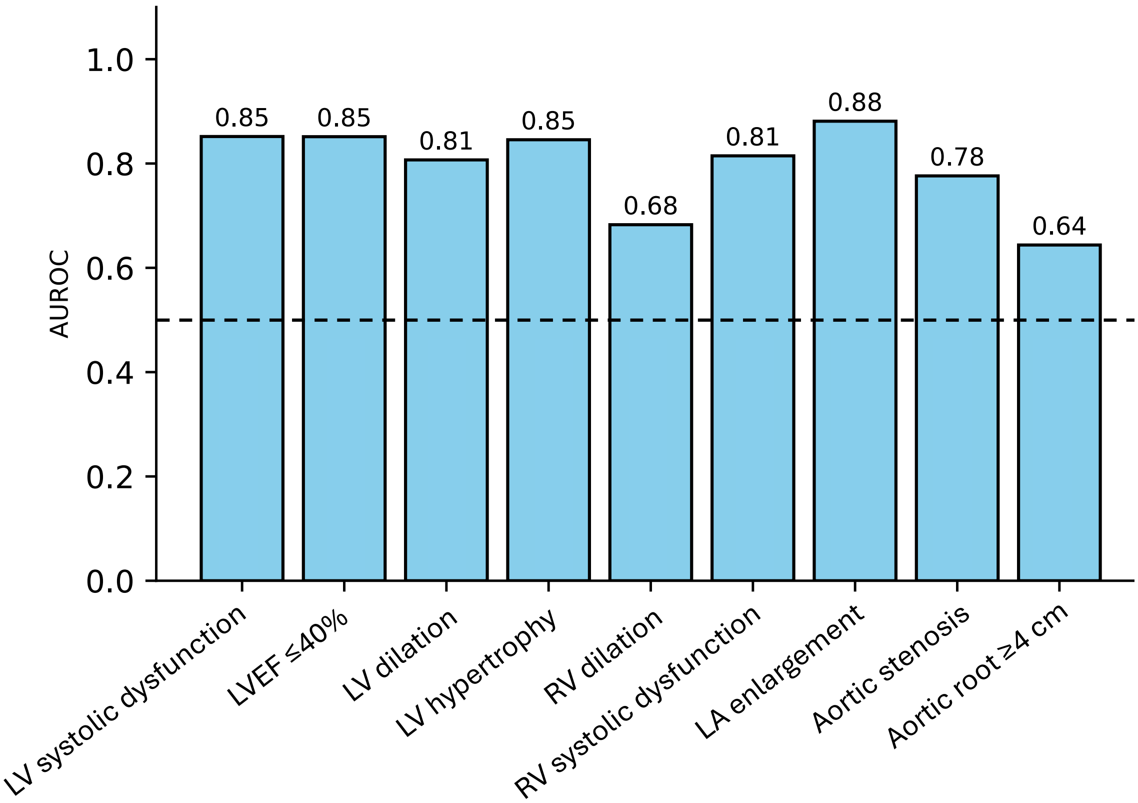


**Fig. S5 | Zero-Shot Transfer Performance of the ECG ViT Model in the UK Biobank Test Set.** Shown are the discrimination performances (area under the receiver-operating-characteristic curve [AUROC]) of a vision transformer (ViT)-based electrocardiogram (ECG) model for detecting structural and functional cardiac phenotypes in the UK Biobank cohort. Centroids for each phenotype were computed from the Yale New Haven Health System (YNHHS) training set and applied directly to the UK Biobank embeddings without retraining or fine-tuning. Each bar represents the mean AUROC for a given echocardiographic phenotype, with values annotated above the bars. Model performance remained robust across diverse SHD phenotypes, including left ventricular (LV) systolic dysfunction, reduced LV ejection fraction (LVEF ≤40%), increased LV size or wall thickness, right ventricular (RV) dysfunction or enlargement, left atrial (LA) enlargement, and aortic stenosis, but was lower for aortic root dilation. AUROC: area under the receiver-operating-characteristic curve; ECG: electrocardiogram; LA: left atrial; LV: left ventricular; LVEF: left ventricular ejection fraction; RV: right ventricular; SHD: structural heart disease; ViT: vision transformer; YNHHS: Yale New Haven Health System.

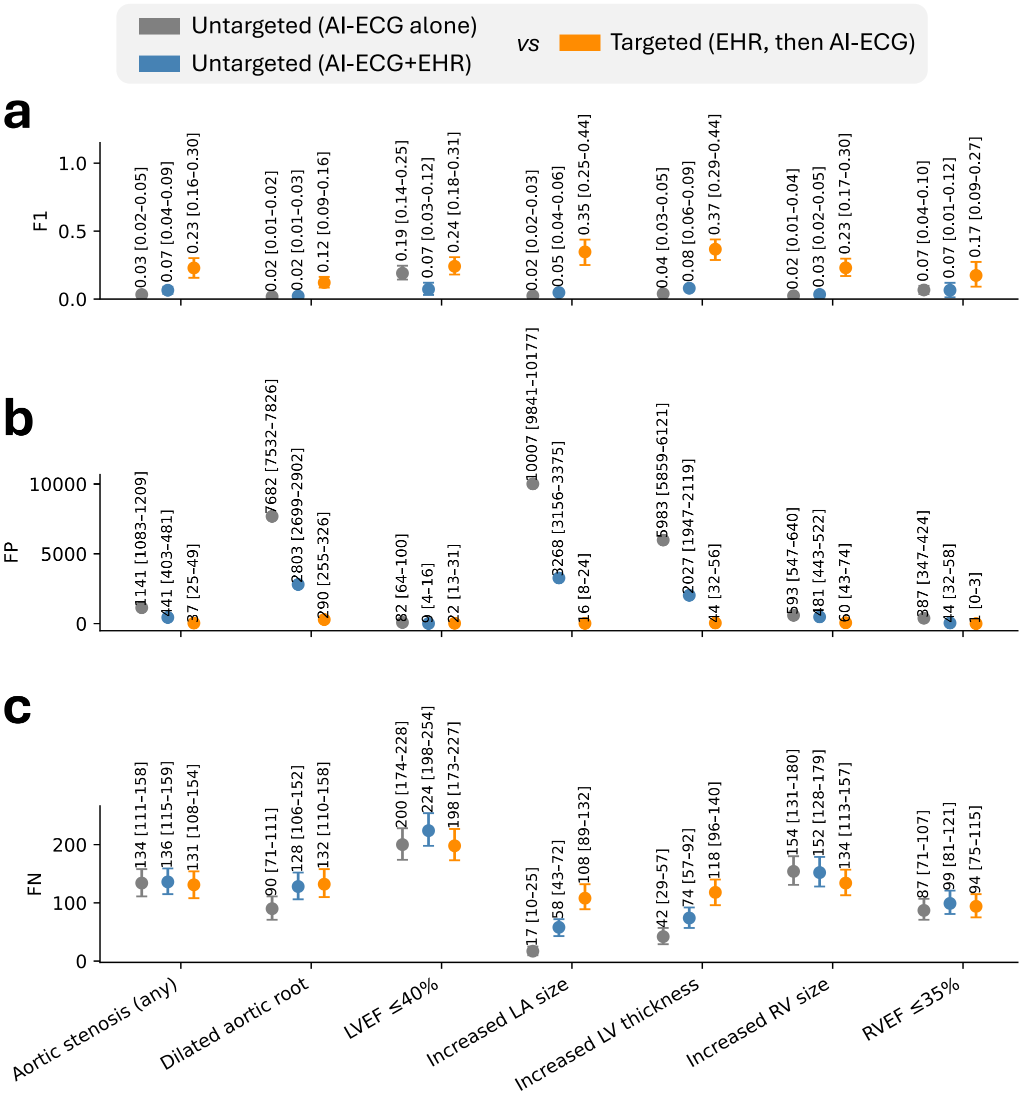


**Fig. S6 | Comparative performance of targeted and untargeted screening strategies for SHD detection in the UK Biobank.** Shown are absolute performance metrics for seven (7) representative cardiac phenotypes in the UK Biobank test cohort, comparing an untargeted image-only ECG model (gray), an untargeted multimodal model combining ECG and EHR embeddings (blue), and a targeted strategy in which EHR-derived embeddings identify high-risk individuals for subsequent ECG model application (orange). Each point represents the mean estimate from 500 paired bootstrap iterations, with vertical bars denoting 95% confidence intervals. (**a**) F1-scores, (**b**) absolute false-positive counts (FP), and (**c**) absolute false-negative counts (FN) for each phenotype and strategy. The targeted approach was associated with consistently higher F1-scores and fewer false positives compared with both untargeted strategies (95% confidence intervals of absolute difference in F1-scores and FP counts derived from bootstrapping with 500 replications were >0 for all pairwise comparisons). AI: artificial intelligence; EHR: electronic health record; ECG: electrocardiogram; FN: false negative; FP: false positive; F1: harmonic mean of precision and recall; LV: left ventricular; LVEF: left ventricular ejection fraction; RV: right ventricular; SHD: structural heart disease.

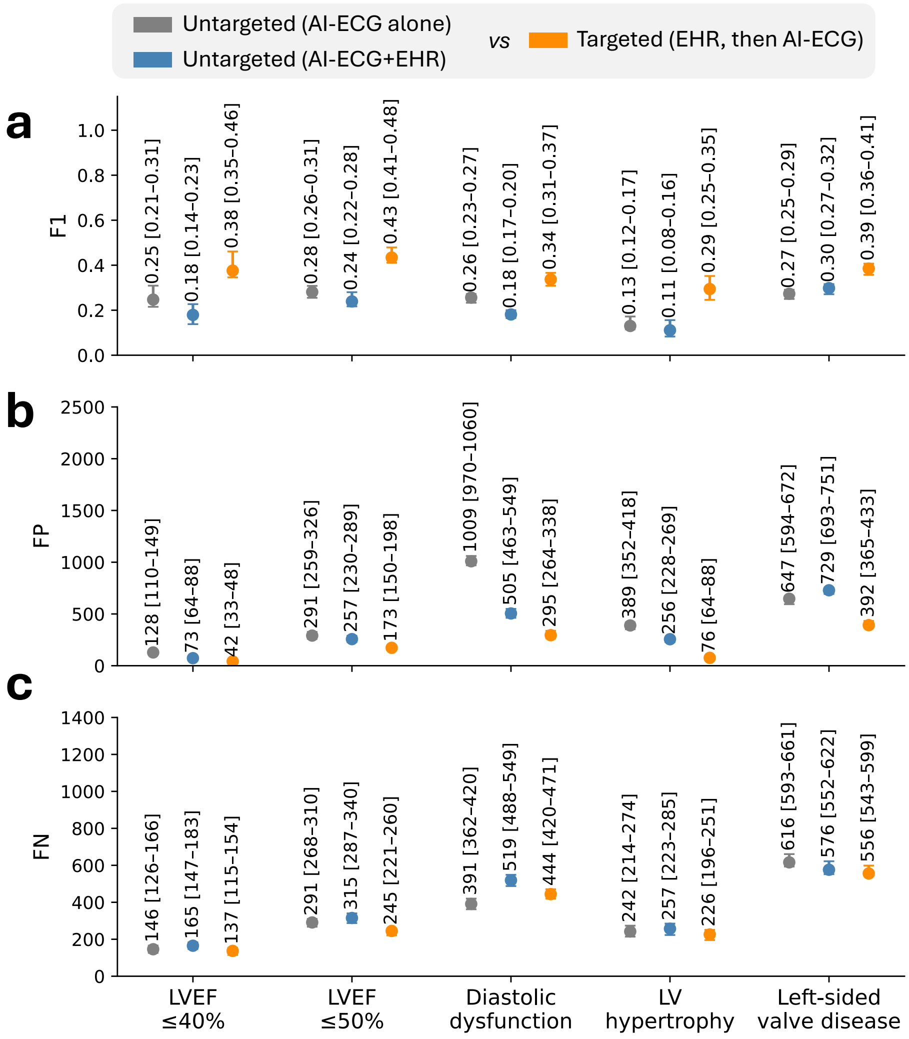


**Fig. S7 | Comparative performance of targeted and untargeted screening strategies for SHD detection in the MIMIC-IV subset**. Shown are absolute performance metrics for 5 representative cardiac phenotypes in the MIMIC-IV external test cohort, comparing an untargeted image-only ECG model (*gray*), an untargeted multimodal model combining ECG and EHR embeddings (*blue*), and a *targeted* strategy in which EHR-derived embeddings identify high-risk individuals for subsequent ECG model application (*orange*). Each point represents the mean value estimated from 500 bootstrap iterations, with vertical bars denoting 95% confidence intervals. (**a**) F1-scores; (**b**) Absolute numbers of false positives (FP); and (**c**) Absolute numbers of false negatives (FN) for each strategy and phenotype. The targeted approach is associated with a significant reduction in both FP counts and improved F1-scores compared with the untargeted approaches (95% confidence intervals of absolute difference in F1-scores and FP counts derived from bootstrapping with 500 replications were >0 for all pairwise comparisons). AI: artificial intelligence; EHR: electronic health record; ECG: electrocardiogram; FN: false negative; FP: false positive; F1: harmonic mean of precision and recall; LV: left ventricular; LVEF: left ventricular ejection fraction.

### Supplemental Tables

#### Table S1 | Definitions for structural heart disease (SHD) labels by echocardiography in YNHHS.

| Label | Positive class |
| --- | --- |
| Left ventricular systolic dysfunction | Moderate or severe (per the final report) |
| Left ventricular ejection fraction (LVEF) | ≤40% or ≤50% |
| Interventricular septum thickness at end-diastole (IVSd) | ≥1.3 cm or ≥1.5 cm |
| Left posterior wall thickness at end-diastole (LVPWd) | ≥1.3 cm or ≥1.5 cm |
| Left ventricular internal diameter at end-diastole (LVIDd) | ≥5.5 cm |
| Left ventricular internal diameter at end-systole (LVIDs) | ≥3.5 cm |
| E/e': ratio of early diastolic mitral inflow velocity to early diastolic mitral annulus velocity | ≥15 |
| Right ventricular systolic pressure (RVSP) | ≥40 or ≥50 mm Hg |
| Tricuspid annular plane systolic excursion (TAPSE) | ≤1.7 cm |
| Tricuspid annular systolic velocity (RV S') | ≤10 cm/sec |
| Aortic valve peak velocity (AV-Vmax) | ≥3 m/sec |
| Aortic root diameter | ≥4 cm |
| Left ventricular wall thickness | Moderate or severe (per the final report) |
| Left ventricular diastolic dysfunction | Moderate or severe (grade 2-3, per the final report) |
| Right ventricular enlargement | Moderate or severe (per the final report) |
| Right ventricular systolic dysfunction | Moderate or severe (per the final report) |
| Left atrial enlargement | Moderate or severe (per the final report) |
| Aortic stenosis | Severe |
| Aortic stenosis | Moderate or severe (per the final report) |
| Mitral stenosis | Any |
| Mitral regurgitation | Moderate or severe (per the final report) |
| Tricuspid regurgitation | Moderate or severe (per the final report) |

### Table S2 | Summary of CLMBR-T vocabulary and mapping to the cohorts used in this manuscript.

| Source ontology (knowledge graph) | Source | Original tokens | Mapped tokens | Cohorts with available data |
|---|---|---|---|---|
| **LOINC** | Logical Observation Identifiers Names and Codes | 37,590 | Selected inclusion of common laboratory measurements: complete blood count, comprehensive metabolic panel, brain natriuretic peptide measurements, troponin, thyroid function, lipid panel, glycated hemoglobin A1c → codes: "2028-9", "2075-0", "2085-9", "2089-1", "2093-3", "2160-0", "2345-7", "2571-8", "2885-2", "2951-2", "3016-3", "3094-0", "30934-4", "33762-6", "42637-9", "42757-5", "43396-1", "4544-3", "4548-4", "6598-7", "6690-2", "67151-1", "6768-6", "704-7", "706-2", "711-2", "713-8", "718-7", "731-0", "736-9", "742-7", "744-3", "751-8", "770-8", "777-3", "785-6", "786-4", "787-2", "788-0", "789-8", "10839-9", "1742-6", "1751-7", "17861-6", "1920-8", "1975-2"] | YNHHS, UKB, MIMIC-IV |
| **SNOMED** | Systematic Nomenclature of Medicine - Clinical Terms | 18,174 | Complete mapping performed | YNHHS, UKB, MIMIC-IV |
| **RxNorm** | RxNorm (NLM) | 4,678 | Complete mapping performed | YNHHS, UKB, MIMIC-IV |
| **CPT4** | Current Procedural Terminology version 4 (AMA) | 3,730 | Complete mapping performed | YNHHS, MIMIC-IV |
| **RxNorm Extension** | OMOP RxNorm Extension | 255 | Not mapped | |
| **ICD10PCS** | ICD-10 Procedure Coding System (CMS) | 233 | Complete mapping performed | YNHHS, MIMIC-IV |
| **ICD9Proc** | International Classification of Diseases, Ninth Revision, Clinical Modification, Volume 3 (NCHS) | 196 | Partial mapping performed | MIMIC-IV |
| **Cancer Modifier** | Diagnostic Modifiers of Cancer (OMOP) | 88 | Not mapped | |
| **HCPCS** | Healthcare Common Procedure Coding System (CMS) | 54 | Not mapped | |
| **ICDO3** | International Classification of Diseases for Oncology, Third Edition (WHO) | 52 | Not mapped | |
| **CVX** | CDC Vaccine Administered CVX (NCIRD) | 41 | Not mapped | |
| **Domain** | OMOP | 27 | Not mapped | |
| **Race** | Race and Ethnicity Code Set (USBC) | 5 | Complete mapping performed | YNHHS, UKB, MIMIC-IV |
| **OMOP Extension** | OMOP Extension (OHDSI) | 3 | Complete mapping performed | YNHHS, UKB, MIMIC-IV |
| **Gender** | OMOP Gender | 2 | Complete mapping performed | YNHHS, UKB, MIMIC-IV |
| **Ethnicity** | OMOP Ethnicity | 2 | Complete mapping performed | YNHHS, MIMIC-IV |
| **CMS Place of Service** | Place of Service Codes for Professional Claims (CMS) | 2 | Not mapped | |
| **Medicare Specialty** | Medicare provider/supplier specialty codes (CMS) | 1 | Not mapped | |
| **Condition Type** | OMOP | 1 | Complete mapping performed | YNHHS, UKB, MIMIC-IV |
| **CARE_SITE** | STANFORD_CUSTOM | 396 | Not applicable | |
| **Visit** | STANFORD_CUSTOM | 6 | Not applicable | |
| **Total counts** | - | 65,536 | 26,874 tokens mapped, including 26,828 [96.0%] out of 27,946 non-LOINC codes | |

**Abbreviations:** AMA: American Medical Association; CMS: Centers for Medicare & Medicaid Services; CLMBRT-: clinical language modeling based representations transformer; CPT4: Current Procedural Terminology version 4; HCPCS: Healthcare Common Procedure Coding System; ICD: International Classification of Diseases; ICDO3: International Classification of Diseases for Oncology, Third Edition; ICD9PCS: International Classification of Diseases, Ninth Revision, Clinical Modification, Volume 3; ICD10PCS: ICD-10 Procedure Coding System; LOINC: Logical Observation Identifiers Names and Codes; MIMIC: Medical Information Mart for Intensive Care; NCHS: National Center for Health Statistics; NCIRD: National Center for Immunization and Respiratory Diseases; NLM: National Library of Medicine; OMOP: Observational Medical Outcomes Partnership; SNOMED: Systematic Nomenclature of Medicine – Clinical Terms; WHO: World Health Organization; YNHHS: Yale-New Haven Health System.

### Table S3 | Examples of SNOMED codes mapped to key concepts for summary demographics.

| Concept | SNOMED Codes |
|---|---|
| Hypertension | ['83105008', '48146000', '199006004', '270440008', '697929007', '185716009', '66610008', '308502002', '308427006', '163028000', '713641000000103', '736286003', '268509003', '302192008', '220901000000101', '194767001', '1201005', '38341003', '1066941000000104', '401118009', '198997005', '73410007', '66052004', '199005000', '1066961000000103', '185723005', '161501007', '845891000000103', '194780003', '163027005', '194783001', '86234004', '275516004', '401117004', '193003', '199007008', '162659009', '308503007', '1083101000000106', '65443008', '863191000000102', '275944005', '185264001', '183856001', '48194001', '908651000000101', '31992008', '64715009', '59621000', '185718005', '1066971000000105', '36221001', '194788005', '846371000000103', '401048005', '170579000', '36315003', '185721007', '810981000000107', '6962006', '50490005', '908631000000108', '199008003', '713661000000102', '78975002', '89242004', '170578008', '194779001', '170577003', '123799005', '194781004', '843821000000102', '185722000', '54225002', '407567007', '170587004', '86041002', '185719002', '170586008', '95691008', '194785008', '70272006', '170588009', '56218007', '766211000000109', '185720008', '308116003', '843841000000109', '1066951000000101', '698640000', '96731000119100', '285831000119108', '371125006', '23717007', '39018007', '62240004', '198954000', '37618003', '198951008', '1474004', '726513006', '129151000119102', '198966006', '851071000000108', '248411000000105', '153891000119101', '31407004', '198949009', '29259002', '390885007', '71874008', '105651000119100', '140111000119107', '428575007', '40521000119100', '23786008', '96741000119109', '34694006', '10757441000119102', '123800009', '49220004', '81626002', '199003007', '712832005', '284961000119106', '698638005', '762463000', '198953006', '709881001', '5501000119106', '712487000', '96751000119106', '134377004', '74451002', '697930002', '237279007', '77970009', '286371000119107', '170571002', '703310005', '134378009', '198967002', '26078007', '417206009', '104931000119100', '59720008', '199000005', '765182005', '10562009', '39727004', '140121000119100', '198965005', '15781000119107', '67359005', '715280009', '59997006', '473392002', '31881008', '284991000119104', '198986005', '199002002', '307632004', '18416000', '16229371000119106', '198942000', '10752641000119102', '140101000119109', '161807003', '52698002', '82771000119102', '288250001', '8762007', '19769006', '118781000119108', '24042004', '429198000', '284981000119102', '398254007', '40511000119107', '48552006', '198984008', '10725009', '111438007', '285011000119108', '198944004', '132721000119104', '720568003', '117681000119102', '65402008', '96711000119105', '57684003', '23130000', '28119000', '541000119105', '285001000119105', '46764007', '16147005', '367390009', '698591006', '170601008', '198985009', '9901000', '8218002', '417322008', '492611000000102', '49102001', '78808002', '706882009', '140131000119102', '96701000119107', '153851000119106', '284971000119100', '41114007', '8501000119104', '14973001', '170572009', '198945003', '845331000000109', '285881000119109', '129181000119109', '198983002', '129161000119100', '247361000000100', '206596003', '198999008', '169465000', '128001000119105', '63287004', '10757481000119107', '198947006', '367821000119106', '237282002', '198968007', '95605009', '170574005', '49171007', '46481004', '170573004', '57873008', '707265005', '198952001', '72022006', '429457004', '84094009', '65518004', '198941007', '10757401000119104', '194791005', '443482000', '96721000119103', '427889009', '78544004', '73030000', '81363003', '66709007', '417287002', '69909000', '285841000119104', '395148004', '285871000119106', '871681000000102', '111411000119103', '35303009', '127991000119101', '285861000119100', '71421000119105', '704667004', '71701000119105', '285851000119102', '170591009', '397748008', '22966008', '15394000', '237281009', '198946002', '170590005', '766937004', '129171000119106', '221281000000109', '843851000000107', '692801000000103', '272821000000100', '221291000000106', '492621000000108', '253041000000107', '1051531000000104', '635071000000100', '478861000000109', '587111000000100', '247111000000104', '648001000000103', '599281000000106', '684221000000101', '892411000000107', '802941000000103', '1076211000000101', '845341000000100', '695711000000100', '672551000000109', '419571000000103', '471521000000108', '606221000000105', '272841000000107', '586581000000102', '599321000000103', '272801000000109', '703671000000107', '492631000000105', '414391000000105', '276789009', '695701000000102', '191241000000108', '565761000000106', '599261000000102', '846381000000101', '892431000000104', '587121000000106', '599341000000105', '11721000000102', '684211000000107', '713651000000103', '187811000000103', '194766005', '187801000000100', '77737007', '645721000000102', '948151000000100', '272781000000108', '766221000000103', '599291000000108', '674201000000103', '832151000000107', '292601000000100', '609021000000109', '635061000000107', '635101000000109', '250411000000105', '191231000000104', '412779008', '167031000000101', '883971000000102', '609011000000103', '250591000000100', '845901000000102', '802951000000100', '908661000000103', '892451000000106', '948161000000102', '843831000000100', '599311000000109', '272811000000106', '892471000000102', '40555009', '187791000000104', '308551004', '647991000000107', '698810000', '883981000000100', '606231000000107', '627791000000106', '3331000000104', '171791000000104', '608991000000100', '627801000000105', '662181000000100', '635081000000103', '908641000000104', '272791000000105', '599271000000109', '606201000000101', '565741000000105', '253221000000105', '11511004', '635091000000101', '544581000000102', '606211000000104', '194774006', '191251000000106', '692791000000102'] |
| Diabetes mellitus | ['112991000000101', '1142044000', '11530004', '1196922005', '1196923000', '1217044000', '1217068008', '1217674007', '1222660008', '1255271005', '127012008', '127013003', '127014009', '1481000119100', '190368000', '190369008', '190372001', '190388001', '190389009', '190390000', '201724008', '230577008', '237599002', '237601000', '237604008', '237612000', '237618001', '237619009', '268519009', '290002008', '313435000', '313436004', '314771006', '314893005', '314902007', '314903002', '314904008', '335621000000101', '368711000119106', '385041000000108', '395204000', '401110002', '408540003', '413183008', '420270002', '420825003', '420868002', '421075007', '421305000', '421437000', '421631007', '421750000', '421847006', '422014003', '422228004', '426705001', '426875007', '427089005', '44054006', '443694000', '444073006', '445353002', '46635009', '4855003', '49455004', '51002006', '5368009', '5969009', '609561005', '609562003', '609572000', '703136005', '703137001', '703138006', '70694009', '73211009', '737212004', '739681000', '75682002', '768792007', '768793002', '768794008', '816067005', '82581000119105', '8801005'] |
| Hypercholesterolemia | ['190774002', '238038003', '238040008', '238076009', '238077000', '238078005', '238079002', '238081000', '238083002', '238086005', '238088006', '267432004', '267434003', '267435002', '275598004', '299465007', '33513003', '34349009', '34528009', '397915002', '398036000', '398796005', '403829002', '403830007', '403831006', '445010006', '57218003', '767133009', '773726000'] |
| Chronic cardiac disease | ['41893002', '253706007', '253431009', '698377004', '281091000', '253274005', '253532004', '253289001', '70602002', '253318000', '54225002', '253677005', '52029003', '253407004', '251024009', '253343006', '253379003', '253678000', '314116003', '253592001', '233875006', '847041000000109', '38385001', '253531006', '204312002', '175042007', '175085003', '232731009', '253384009', '253676001', '232739006', '253650001', '837091000000100', '93353003', '394710008', '129574000', '253346003', '253322005', '253453006', '253628002', '253568009', '268184003', '426104007', '1755008', '76593002', '410023005', '253370004', '395105005', '308066006', '42343007', '174814003', '812001000000102', '85081000', '37104009', '414795007', '253401003', '4374004', '428558001', '253476009', '253512000', '194823009', '253629005', '277639002', '703518004', '425785006', '422348008', '253548003', '253273004', '253281003', '232735000', '253642005', '68466008', '253455004', '56675007', '111321007', '253305008', '717501000000108', '253637002', '426300009', '447672009', '253324006', '50570003', '233127003', '59631007', '253524007', '36969009', '281170005', '65340007', '175037004', '233871002', '253721001', '253374008', '253683008', '253578007', '275253006', '240567009', '15990001', '715971000000103', '86299006', '427109009', '253732001', '204297006', '57054005', '67278007', '253658008', '253412003', '54329005', '28574005', '253634009', '277192005', '253496001', '253280002', '77978002', '253441007', '194857001', '253590009', '414024009', '253477000', '82458004', '443502000', '233821000', '253518001', '194781004', '175076003', '253338004', '232723004', '253290005', '199221000000102', '233841005', '253415001', '253555001', '95268002', '204306007', '111322000', '175071008', '30288003', '253437008', '174822005', '233832000', '233843008', '88805009', '275521001', '42402006', '87648004', '199006004', '204296002', '204300001', '13213009', '123661007', '253508006', '253627007', '204443008', '253265008', '310490008', '253615009', '397193006', '253438003', '253707003', '86252004', '300995000', '73795002', '225566008', '233829003', '253472006', '253715000', '79009004', '253294001', '2583009', '253724009', '811901000000107', '315614006', '232722009', '174809002', '93271008', '84114007', '253607006', '253562005', '253359006', '40964007', '253264007', '253418004', '22462001', '253291009', '253563000', '253708008', '29928006', '253500004', '253633003', '781051000000108', '87837008', '253675002', '233831007', '175036008', '175047001', '253436004', '233840006', '447670001', '234010000', '6996004', '763641000000102', '36110001', '204311009', '265481001', '85232009', '253300003', '398754006', '253691004', '253353007', '233924009', '204395001', '234029006', '399211009', '32413006', '233929004', '253434001', '268180007', '253652009', '253465005', '253530007', '174826008', '315348000', '233138003', '18546004', '308065005', '195023001', '311796008', '83119008', '26900001', '233825009', '21981000', '253515003', '218728005', '90828009', '19092004', '717481000000104', '213152006', '174810007', '253668003', '253601007', '401251008', '194780003', '253643000', '170595000', '253725005', '416158002', '194767001', '253304007', '253593006', '253575005', '95234008', '61959006', '85053006', '253397006', '204357006', '194779001', '300920004', '416610007', '810681000000101', '206586007', '175041000', '194821006', '233846000', '23063005', '174815002', '253581002', '250983006', '253299006', '253569001', '175029007', '253657003', '716621000000101', '253286008', '253602000', '233826005', '194843003', '232717009', '275514001', '408471009', '28903008', '86234004', '161504004', '309814006', '253295000', '78069008', '253341008', '45237002', '41339005', '40250003', '384678000', '109428005', '521691000000100', '195025008', '253624000', '164869004', '271573009', '253336000', '175066001', '253534003', '240077004', '45471009', '341751000000103', '253689007', '94706008', '447698003', '253382008', '253566008', '253610004', '253414002', '286950001', '205769006', '253426007', '253357008', '92506005', '398274000', '247611000000109', '204431007', '445928005', '275215001', '161502000', '253360001', '83521008', '253481000', '22298006', '45492009', '232724005', '253507001', '203741000000101', '253663007', '43736008', '253556000', '253430005', '756003', '253527000', '384681005', '253335001', '253393005', '426911005', '199007008', '17024001', '195147006', '16972009', '253489003', '253480004', '27637000', '253395003', '253599005', '78643003', '46109009', '175061006', '389026000', '253387002', '233112003', '75372006', '232719007', '253419007', '441541008', '194809007', '253302006', '311792005', '253713007', '15459006', '232720001', '703272007', '286721000000101', '233835003', '253411005', '253381001', '111323005', '253717008', '253625004', '253432002', '161503005', '191372008', '253542002', '233877003', '253413008', '253458002', '253422009', '253482007', '204339005', '253621008', '253326008', '20738002', '19249002', '49584005', '204379001', '253720000', '311793000', '19242006', '164871004', '253345004', '232738003', '175045009', '232729000', '73699003', '274947007', '253288009', '401250009', '253714001', '90539001', '429589006', '253539008', '253391007', '401252001', '253711009', '253425006', '253484008', '170597008', '174808005', '253278008', '232726007', '204398004', '309518001', '204351007', '253378006', '36233006', '56501009', '253603005', '174830006', '253719006', '253586007', '281093002', '851521000000102', '853561000000109', '233823002', '12075007', '194845005', '36221001', '40541001', '109430007', '253445003', '232737008', '233836002', '253272009', '421704003', '427188004', '253553008', '268174004', '775171000000109', '233880002', '253405007', '233834004', '253546004', '813991000000101', '70422006', '253700001', '268185002', '174802006', '253283000', '736968006', '394659003', '775161000000102', '427886002', '401314000', '253376005', '253722008', '253559007', '264503005', '194802003', '399261000', '308068007', '253267000', '205834002', '412787009', '253565007', '194842008', '253420001', '253554002', '253574009', '410431009', '253547008', '395723006', '51442005', '315608004', '78495000', '717171000000109', '174833008', '7368005', '314207007', '715951000000107', '269299003', '253456003', '194849004', '233873004', '717541000000106', '286947004', '307140009', '253688004', '194868001', '407596008', '276514007', '253373002', '447671002', '315026000', '2829000', '253487001', '204352000', '253544001', '253632008', '64077000', '461105005', '264086008', '16567006', '175050003', '253389004', '90383006', '204362007', '253567004', '21470009', '60106004', '265467003', '253504008', '401303003', '83698002', '60232001', '275227003', '717531000000102', '175115008', '253452001', '287280009', '37034001', '78250005', '232712002', '253448001', '261756009', '253329001', '20529002', '27097002', '371068009', '194856005', '253511007', '10633002', '253703004', '1871002', '204456001', '247361000000100', '253672004', '161666008', '128404006', '253417009', '253270001', '415295002', '204399007', '253328009', '253543007', '253597007', '253660005', '253614008', '716211000000105', '253690003', '253520003', '5230009', '74218008', '253570000', '789621000000105', '304914007', '233828006', '204363002', '253443005', '195020003', '287277008', '253269002', '253600008', '420913000', '253510008', '174832003', '253523001', '253550006', '224161000000105', '10451007', '194867006', '195112003', '79955004', '204330009', '66052004', '233126007', '72352009', '315287002', '424045003', '204448004', '233837006', '253545000', '253402005', '716971000000109', '170594001', '233830008', '253495002', '204423002', '309519009', '426902005', '29843007', '253321003', '59877000', '19057007', '194863005', '253312004', '253271002', '253386006', '872361000000105', '3545003', '253529002', '811701000000105', '62335009', '64715009', '195021004', '91335003', '309634009', '253674003', '378121000000101', '28931004', '261538006', '40272001', '448302004', '253317005', '174803001', '204299009', '253499008', '253284006', '253609009', '315025001', '764851000000102', '233858000', '204378009', '253447006', '81990004', '253576006', '80387009', '253470003', '233838001', '253584005', '62067003', '253723003', '95441000', '253573003', '374231000000104', '253303001', '253435000', '253636006', '429639007', '15096009', '204354004', '253479002', '42531007', '394724007', '275516004', '195111005', '253277003', '75270000', '7438000', '253635005', '232921000000108', '32477003', '170596004', '253623006', '45227007', '204391005', '253297008', '936451000000108', '253451008', '915571000000102', '308117007', '253654005', '233844002', '55510008', '95237001', '233842003', '48520006', '85898001', '204457005', '303272002', '873881000000100', '233839009', '194865003', '253664001', '253366007', '253423004', '253644006', '60787001', '253460000', '7305005', '232728008', '26201005', '68092007', '174818000', '732230001', '253580001', '287283006', '253596003', '253591008', '48121000', '213037002', '253514004', '253498000', '253611000', '277638005', '719678003', '195027000', '736966005', '237227006', '35728003', '399020009', '54682008', '253275006', '253486005', '253564006', '68237008', '253588008', '427585005', '253704005', '717491000000102', '395704004', '253334002', '63042009', '204345002', '275511009', '253540005', '253552003', '253646008', '253356004', '253537005', '204464007', '24355008', '253377001', '164868007', '448794008', '253301004', '164870003', '736969003', '14886009', '408546009', '253579004', '28656008', '31413008', '93384001', '253375009', '164865005', '13689005', '253583004', '232736004', '390799004', '194866002', '253712002', '853591000000103', '26780008', '716091000000107', '717191000000108', '253716004', '204343009', '204394002', '66403007', '44777001', '66610008', '261755008', '58612006', '23685000', '253718003', '204370002', '93059006', '134401001', '14201006', '233827001', '253268005', '253388007', '401253006', '367363000', '233847009', '253421002', '233819005', '715981000000101', '253468007', '302943003', '174817005', '422293003', '53741008', '70211005', '204433005', '175038009', '253348002', '253582009', '253333008', '414089002', '232727003', '704207005', '426611007', '194828000', '195029002', '3546002', '54160000', '253536001', '275252001', '253337009', '253598002', '204397009', '161505003', '92960007', '195130005', '415070008', '253394004', '204463001', '175040004', '413838009', '67682002', '31019002', '39905002', '10273003', '62695002', '7484005', '251038002', '253571001', '204346001', '253594000', '195114002', '276517000', '281092007', '414545008', '253533009', '233845001', '175048006', '224111000000108', '262941008', '194858006', '174911007', '360371003', '63739005', '11101003', '91634006', '253446002', '253608001', '277994007', '204451006', '437746009', '253463003', '253494003', '204368006', '12770006', '1083131000000100', '90453003', '248461000000107', '387732009', '233878008', '253525008', '204427001', '70320004', '253439006', '233817007', '170593007', '213151004', '253354001', '253298003', '253457007', '413844008', '253467002', '253369000', '253287004', '204342004', '253655006', '233833005', '234132006', '26146002', '195030007', '390884006', '83105008', '253285007', '253492004', '253509003', '253293007', '63934006', '420300004', '23687008', '45503006', '253587003', '195032004', '698378009', '71892000', '253282005', '253649001', '416683003', '250942008', '253521004', '253656007', '314206003', '446221000', '223301000000106', '204467000', '250941001', '270510008', '195031006', '175039001', '253471004', '373093003', '72972005', '704097008', '253404006', '204319006', '87343002', '253279000', '253364005', '707828002', '253485009', '384677005', '253622001', '62377009', '174831005', '253342001', '253403000', '253310007', '297160003', '164867002', '25106000', '717111000000104', '253653004', '253276007', '253585006', '75398000', '13867009', '428411007', '253681005', '52035003', '36422005', '253392000', '253673009', '253450009', '194862000', '9909003', '224181000000101', '253680006', '253349005', '253710005', '253429000', '83799000', '397194000', '65547006', '59021001', '35928006', '253330006', '253399009', '253416000', '275216000', '253442000', '4557003', '233930009', '253396002', '29899005', '308118002', '253638007', '202231000000106', '175021005', '253513005', '253505009', '253551005', '704096004', '11614003', '233970002', '253327004', '36315003', '10818008', '277534001', '939881000000105', '66189004', '253682003', '390885007', '276516009', '72242008', '253440008', '425654008', '407597004', '699245006', '253631001', '253323000', '448619007', '236006000', '718556007', '285991000119100', '449536002', '204981000119101', '448646002', '720605009', '253538000', '715865008', '16440002', '429347004', '461345005', '1092801000119102', '447498008', '448919005', '472318007', '766883006', '449111002', '233711002', '1641000119107', '405598005', '725145002', '460307002', '1089471000000109', '93050005', '25544003', '29375001', '719824001', '448633007', '12367281000119109', '10519008', '448968005', '472100003', '449436000', '449455000', '460387004', '460590006', '24088005', '92969008', '472320005', '443391000', '11018701000119109', '459182005', '726083008', '196151000', '447669002', '96731000119100', '89461002', '47800001', '723304001', '448923002', '764697003', '447860000', '763279007', '233705000', '429809004', '759561000000108', '447913009', '418304008', '471841009', '449014008', '449178002', '460517008', '712866001', '449383004', '67782005', '720567008', '448824007', '72252007', '447845003', '15629741000119102', '51789008', '609155001', '447934002', '447703009', '448843007', '49699002', '60856006', '448819009', '447824005', '448645003', '448095003', '30277009', '264571006', '83014000', '787001', '445285002', '472315005', '447939007', '85503007', '707476006', '448947009', '12023003', '447928009', '253308005', '371806006', '703276005', '447668005', '285251000119101', '111319002', '449040002', '449560003', '471875005', '698296002', '703251009', '449385006', '728461000000101', '449031001', '429245005', '15960661000119107', '460592003', '448099009', '417996009', '67441000119101', '449512001', '78196008', '43910005', '448073002', '448066007', '472795006', '413995009', '17073005', '698740005', '759881000000102', '89222008', '445334007', '363125002', '471276003', '418044006', '44630002', '449123008', '460596000', '15964981000119104', '447998001', '443254009', '51201009', '2024009', '88445007', '91338001', '447988007', '448100001', '461107002', '399046008', '89687005', '449316002', '39202005', '79754008', '699297004', '30884007', '762253006', '472326004', '36133000', '85971001', '759761000000102', '460593008', '59063002', '89323001', '461359003', '443344007', '460913001', '339909005', '450311005', '461438006', '448007000', '448028009', '4313003', '449315003', '123660008', '448747000', '449351005', '89456004', '763834000', '448471006', '448497000', '448083003', '92944008', '15712841000119100', '15960981000119105', '360473004', '445284003', '726581000000107', '8501000119104', '447941008', '449538001', '449467003', '33644002', '448074008', '446916007', '445330003', '703164000', '95236005', '699298009', '445176007', '448624005', '41371000119100', '163309001', '448637008', '703274008', '459066006', '286011000119101', '449140004', '76931005', '359597003', '13453009', '445350004', '448475002', '108701000119102', '461383006', '448004007', '39987008', '284881000119108', '60514000', '10757401000119104', '128557009', '459062008', '449228007', '441530006', '286031000119106', '759931000000105', '472797003', '233708003', '471278002', '448092000', '721009008', '472820000', '12238111000119106', '204318003', '101281000119107', '723867002', '447673004', '80762004', '458088003', '233124005', '448098001', '448729006', '448629000', '448595006', '448060001', '448975006', '16228931000119102', '449551006', '428196007', '448105006', '726011000', '450302005', '448104005', '8712002', '460370001', '85041007', '472101004', '449492000', '707200002', '447874007', '240629003', '54830008', '703253007', '471890009', '364006', '728471000000108', '399228007', '64041007', '10335000', '111284004', '447838003', '473444001', '472796007', '46113002', '448181004', '120851000119104', '459180002', '15713081000119108', '31085000', '94703000', '449232001', '36471000', '461111008', '461433002', '703165004', '285981000119103', '703212004', '120891000119109', '448001004', '447861001', '736964008', '447689001', '81552002', '448320008', '232973007', '763778003', '371808007', '459183000', '66989003', '5203004', '84733009', '764965000', '123642008', '426263006', '448745008', '448304003', '59464004', '736962007', '16253001', '461629004', '63183009', '719451006', '449532000', '360481003', '448414008', '71603004', '15713161000119100', '448871001', '15712961000119108', '371811008', '717812000', '447932003', '93262004', '445131007', '414088005', '75942006', '30670000', '109429002', '449146005', '713523008', '71799002', '447850009', '447681003', '21057005', '445310002', '96751000119106', '413444003', '68602002', '460531008', '18192007', '448627003', '15964701000119109', '723448007', '253728007', '30496006', '722051004', '461331004', '80331001', '10930001', '448061002', '43121002', '46696008', '447997006', '719379001', '15544002', '461557000', '373131000', '264162009', '461112001', '447968008', '707478007', '405599002', '15629591000119103', '759771000000109', '15629541000119106', '21789005', '445106006', '448625006', '471288001', '448510002', '128584005', '370141003', '253641003', '15960141000119102', '445209005', '448920004', '447822009', '15712881000119105', '371862006', '448063004', '5148006', '18687009', '371809004', '263960005', '759731000000107', '447772003', '92830003', '57803009', '276526002', '764452004', '736973000', '153941000119100', '406428006', '54854003', '445027003', '371805006', '83978005', '448154004', '86995005', '446668003', '10091002', '93558006', '461110009', '128563000', '7169009', '204315000', '720639008', '447697008', '15960061000119102', '447937009', '471858004', '109431006', '449354002', '111285003', '194709000', '447938004', '460500001', '426012001', '41964001', '736963002', '1131009', '723993005', '278928000', '78862003', '461572001', '472853005', '371803003', '471292008', '447840008', '460906001', '723858002', '720610008', '460595001', '446657003', '471279005', '447812003', '12963004', '448575000', '50270004', '460594002', '70142008', '84725008', '447876009', '702424003', '59494005', '725416005', '233710001', '72481000119103', '449459006', '717943008', '449118008', '609507007', '471290000', '717859007', '448155003', '447701006', '86194008', '84871007', '471880001', '93076005', '724550005', '447661004', '1474004', '719400000', '92941000', '472803004', '74249003', '33258008', '253347007', '338892004', '460538002', '472327008', '284871000119105', '472787007', '64072006', '23341000119109', '54073003', '461587006', '448793002', '443725002', '457612009', '471293003', '735566005', '20005002', '15960381000119109', '111000119104', '703209002', '461577007', '119565001', '448827000', '419132001', '460597009', '445167000', '928341000000103', '74960003', '371812001', '471286002', '443343001', '698640000', '446670007', '371807002', '129582000', '39693003', '117051000119103', '759461000000109', '448278002', '449139001', '471291001', '128555001', '276518005', '268187005', '470752003', '723089002', '55565007', '447915002', '448357002', '123641001', '253698005', '448085005', '28284007', '90487008', '88318005', '724437007', '450310006', '448612003', '119564002', '40023000', '89819002', '460437005', '722461004', '253692006', '446326008', '449600009', '470758004', '78741000119103', '42481001', '461420008', '92923008', '447286000', '709981002', '447839006', '448303009', '449428009', '16063004', '472319004', '26865008', '1089441000000103', '447702004', '471287006', '460365008', '460588005', '81266008', '735581004', '460510005', '822501000000101', '449567000', '449568005', '128567004', '448604006', '119773008', '284861000119104', '15960581000119102', '128568009', '447285001', '448331000', '449563001', '448780007', '728481000000105', '735571003', '449353008', '445264002', '718713000', '78320000', '447284002', '18037001', '736972005', '15256002', '726704006', '448305002', '447571007', '449029005', '698594003', '448310003', '61490001', '448516008', '35577008', |

| | |
|---|---|
| | '447962009', '472783006', '442304009', '31080005', '448084009', '448097006', '449085001', '762251008', '461384000', '53703001', '472793004', '766976003', '447829000', '447667000', '443380007', '450305007', '472855003', '698638005', '70998009', '413905004', '58629009', '13963007', '449350006', '448161000', '459165008', '448328001', '448079003', '736970002', '448946000', '448630005', '470753008', '443389008', '448087002', '80479009', '736965009', '448082008', '447683000', '413439005', '384680006', '1082601000119104', '233601000000100', '449537006', '444354004', '93247001', '253371000', '721976003', '43176009', '763835004', '11433004', '109379000', '447827003', '93234006', '447283008', '443253003', '449596004', '763066009', '458039003', '447690005', '7991000119102', '446909006', '39724006', '459181003', '445002005', '447773008', '445298006', '92976003', '130541000119100', '703210007', '447318008', '398716006', '250999000', '447933008', '96711000119105', '33006007', '449015009', '162970000', '448681000', '46165000', '69609002', '253358003', '703245004', '41049003', '371804009', '471299004', '461104009', '448086006', '448308000', '15713041000119103', '46939000', '715626008', '39589002', '725027004', '759551000000105', '460312001', '717840005', '470757009', '736971003', '108421000119102', '91604001', '109432004', '707372009', '445435009', '92963009', '461376004', '90205004', '14323007', '415509000', '449511008', '447528003', '94702005', '472103001', '445001003', '445898001', '286071000119109', '65861005', '443379009', '59062007', '764955006', '232733007', '19036004', '473393007', '448118003', '62207008', '716740009', '15713121000119105', '448116004', '448356006', '428501002', '418551006', '448744007', '78031003', '721014007', '759741000000103', '718216009', '11468004', '461592008', '445235006', '82523003', '17531000119105', '93237004', '715535009', '448966009', '21234008', '736967001', '767311002', '445336009', '449456004', '448721009', '93064005', '447696004', '449086000', '163311005', '448059006', '10326007', '449025004', '448183001', '10757441000119102', '312591002', '759781000000106', '449398002', '93305002', '444217005', '53043001', '445003000', '461385004', '447664007', '8239009', '471301006', '36665001', '460937001', '445286001', '724208006', '718181001', '703213009', '85284003', '78485007', '26942004', '449005003', '472703008', '472786003', '449125001', '426856002', '445373007', '62820003', '445650008', '447780005', '697976003', '448080000', '96701000119107', '449441008', '735580003', '462172006', '153931000119109', '460930004', '92888002', '253679008', '39041004', '721013001', '459065005', '713689002', '763725002', '759851000000108', '719272007', '233707008', '703211006', '726571000000105', '460471001', '719380003', '74371005', '447274003', '719395001', '444176000', '253519009', '78609007', '139011000119104', '21430002', '26170003', '4501007', '10365005', '728391000000103', '28869005', '703244000', '123658006', '443330001', '53322007', '83898004', '461366002', '448887003', '405752007', '447919008', '52757001', '715987000', '163313008', '449593007', '448162007', '449016005', '763280005', '461109004', '762460002', '23311000119105', '11104006', '59554006', '34126003', '449158005', '128558004', '461421007', '79439001', '461414007', '93078006', '460581004', '718681002', '71346006', '724066002', '448472004', '53067009', '471297002', '449589001', '371810009', '93053007', '17759006', '448182006', '123656005', '13839000', '28381002', '791000119109', '724173009', '448746009', '445208002', '15712921000119103', '233123004', '460438000', '703356002', '449110001', '472317002', '460589002', '445453003', '472323007', '449124002', '41334000', '233125006', '15676002', '276637009', '74034002', '72011007', '448332007', '195137008', '1085021000119106', '47058000', '429559004', '471846004', '399216004', '450313008', '1321241000000105', '16855001', '67431000119105', '68800008', '45669002', '448724001', '429620002', '447929001', '5375005', '253730009', '49102001', '448309008', '448280008', '471277007', '360465008', '82522008', '447999009', '24584003', '461413001', '67696008', '728491000000107', '460524009', '359617009', '93347003', '471280008', '759891000000100', '397431004', '471274000', '718135001', '93069000', '74908007', '233712009', '472785004', '447903001', '461000119108', '1082611000119101', '44088000', '445436005', '234106000', '83130006', '410065004', '448027004', '67166004', '405276000', '119772003', '703519007', '163312003', '460545002', '720606005', '448064005', '60732002', '449539009', '707477002', '427919004', '96311000119109', '441481004', '449430006', '18932005', '461597002', '253697000', '460582006', '461382001', '419097006', '445351000', '450301003', '35719004', '461567005', '448876006', '91529004', '722919003', '8876004', '729581004', '58911001', '698592004', '448081001', '41514002', '67399008', '728401000000100', '767309006', '56309007', '700065003', '52985009', '449490008', '460591005', '448628008', '462174007', '719456001', '448120000', '5053004', '727221000000107', '15781000119107', '25559009', '94150003', '77970009', '232974001', '460380002', '718128009', '22966008', '450312003', '196152007', '448478000', '472102006', '93075009', '448790004', '96741000119109', '445607003', '109425008', '18919000', '449433008', '763615003', '448119006', '473362006', '768552007', '128556000', '96721000119103', '253577002', '92974000', '448153005', '450315001', '447901004', '721073008', '715655000', '724435004', '48872007', '449098005', '17718000', '445268004', '64615000', '12238151000119107', '55546004', '447849009', '233709006', '471885006', '443765003', '472769009', '93051009', '449442001', '40532006', '720748007', '421327009', '460923005', '447666009', '460586009', '264467005', '448115000', '444851003', '444961009', '762252001', '720609003', '700458001', '1954004', '36079008', '194750008', '728381000000100', '471289009', '471870000', '307311001', '63402005', '722095005', '92517006', '450308009', '722027009', '471298007', '74561007', '703275009', '759751000000100', '48431000', '418824004', '448103004', '448415009', '764521002', '443296008', '109426009', '449429001', '75409000', '703214003', '471285003', '447852001', '447289007', '448842002', '54636000', '28248000', '93287006', '446890001', '713405002', '460583001', '723333000', '75761004', '56941009', '703273002', '64862009', '82247006', '32912007', '448184007', '263944006', '448723007', '448634001', '448117008', '120861000119102', '56276002', '196153002', '448477005', '449595000', '448078006', '716773002', '86466006', '1086051000119107', '445270008', '450309001', '461381008', '727071000000103', '445236007', '449594001', '735565009', '448062009', '48447003', '15713201000119105', '52048004', '703385008', '766751007', '449138009', '759521000000100', '448330004', '448809003', '449032008', '450307004', '120881000119106', '447774002', '418461002', '449576007', '236004002', '449270002', '83291003', '31881008', '450314002', '77696009', '90202001', '472324001', '449133000', '44313006', '93445007', '460585008', '461436005', '17394001', '76257003', '405768001', '448072007', '461371009', '424404003', '10971000087107', '73660006', '449457008', '448844001', '448517004', '447700007', '445454009', '726335002', '448277007', '34342000', '95440004', '213215000', '84648007', '449587004', '762433009', '420006002', '448948004', '50275009', '461439003', '722206009', '763747002', '64749001', '443166007', '719839000', '462175008', '473443007', '449520004', '472807003', '447935001', '448823001', '450300002', '460587000', '359601003', '448528000', '445636003', '448611005', '75205009', '10757481000119107', '44993000', '736978009', '446659000', '445299003', '704242009', '29819009', '79152002', '236003008', '6022005', '724436003', '472801002', '109427000', '2816000', '446667008', '448208003', '61233003', '448075009', '448778001', '449190000', '460944005', '721010003', '472794005', '64627002', '449427004', '450306008', '46847001', '450304006', '759861000000106', '707371002', '713617008', '721978002', '447970004', '236005001', '448577008', '282006', '459164007', '447830005', '25263003', '719835006', '460584007', '449440009', '472316006', '716193004', '449188001', '759871000000104', '1089431000000107', '413994008', '472854004', '32574007', '449382009', '153951000119103', '728371000000102', '460375006', '430396006', '12363009', '723336008', '704241002', '91442002', '703258003', '233706004', '42866003', '449352003', '447665008', '95437004', '1089451000000100', '8209004', '12563008', '449559008', '449547009', '703252002', '447930006', '461435009', '431667009', '449379004', '722006004', '10674871000119105', '699668009', '253729004', '449087009', '42099002', '194741006', '735579001', '111283005', '445440001', '449479003', '703360004', '61012002', '720858001', '461434008', '263961009', '10190003', '472325000', '461562004', '449599006', '783611000000101', '194798004', '589911000000106', '251961000000106', '492711000000108', '538561000000104', '539061000000102', '205890008', '728421000000109', '565751000000108', '589871000000109', '320371000000101', '259501000000101', '589901000000109', '753551000000108', '223171000000101', '727621000000105', '677131000000105', '464231000000104', '259051000000108', '232971000000107', '726701000000105', '676131000000108', '368871000000102', '841141000000104', '320361000000108', '753561000000106', '381191000000102', '182551000000100', '639231000000108', '397041000000101', '32598000', '419231000000102', '232961000000100', '38801000000100', '175032005', '258871000000100', '64333001', '581511000000103', '578421000000109', '918151000000107', '536971000000101', '712421000000102', '233879000', '253041000000107', '539081000000106', '591181000000101', '627861000000109', '253560002', '297401000000100', '614081000000109', '726941000000105', '250471000000100', '89791000000100', '442511000000106', '274661000000100', '726881000000109', '659411000000100', '727921000000101', '175031003', '684221000000101', '599271000000109', '248531000000101', '728711000000106', '727521000000109', '251681000000108', '191651000000108', '751161000000102', '569961000000105', '728601000000102', '258811000000105', '676111000000100', '659421000000106', '204376008', '668811000000101', '297461000000101', '726671000000106', '253661009', '726491000000108', '727771000000100', '111282000', '444321000000101', '917861000000101', '223631000000109', '588811000000108', '727971000000102', '582901000000101', '338101000000107', '253666004', '237241000000107', '727241000000100', '128572008', '776291000000108', '656811000000106', '608451000000100', '194766005', '176331000000107', '507621000000100', '266242007', '1064941000000106', '713211000000101', '728151000000109', '726961000000106', '585651000000102', '249291000000108', '728821000000107', '253557009', '52295007', '448114001', '491601000000108', '727541000000102', '284831000119107', '728431000000106', '407605008', '612641000000100', '407607000', '82109007', '726591000000109', '38781000000101', '728141000000106', '425961000000106', '728291000000106', '37689002', '325431000000103', '259041000000105', '591161000000105', '316095000', '643821000000108', '194832006', '727981000000100', '727871000000105', '727111000000109', '88596007', '776301000000107', '727701000000108', '727891000000109', '890631000000101', '841981000000109', '223311000000108', '764861000000104', '586341000000100', '251971000000104', '373145006', '727641000000103', '728451000000104', '464561000000107', '752191000000102', '202251000000104', '853601000000109', '31420001', '668581000000106', '92518001', '259471000000108', '472771009', '727751000000109', '586381000000108', '258631000000103', '253572008', '750781000000101', '409621000000103', '223181000000104', '248701000000104', '212721000000104', '224061000000109', '432391000000106', '396561000000107', '207252009', '516941000000102', '224121000000102', '3192005', '517421000000104', '540201000000103', '885321000000101', '253101000000105', '400941000000105', '443626009', '555721000000106', '749771000000107', '894611000000100', '469611000000108', '783601000000103', '727901000000105', '491721000000105', '449829009', '250411000000105', '251025005', '728031000000101', '237281000000104', '258581000000100', '881331000000102', '687081000000103', '664221000000104', '232951000000103', '727951000000106', '416461000000103', '89600009', '469031000000107', '214611000000109', '516841000000106', '233661000000101', '873891000000103', '727991000000103', '29201009', '526811000000101', '492051000000105', '727491000000106', '589881000000106', '727631000000107', '728181000000103', '1033151000000100', '837101000000108', '811911000000109', '271441000', '698361000000107', '175009006', '924381000000104', '692851000000102', '583961000000105', '439211000000102', '692861000000104', '258621000000100', '519531000000108', '727531000000106', '581151000000108', '248511000000109', '595941000000109', '43395008', '367416001', '282811000000103', '297411000000103', '776251000000100', '259021000000103', '603901000000104', '274641000000101', '259821000000107', '727781000000103', '24080003', '259491000000107', '728341000000108', '381181000000104', '248521000000103', '891411000000100', '428191000000108', '752181000000104', '726411000000104', '657501000000108', '753501000000107', '224131000000100', '757071000000102', '253558004', '516951000000104', '373146007', '727711000000105', '727661000000102', '175022003', '321551000000106', '728701000000109', '259001000000107', '274931000000102', '451911000000108', '259481000000105', '274561000000104', '726931000000101', '568721000000106', '728051000000108', '728661000000103', '621571000000106', '259811000000101', '596001000000100', '274941000000106', '591061000000109', '728261000000100', '727911000000107', '170598003', '728021000000103', '789631000000107', '248381000000108', '709791000000102', '727101000000107', '175026000', '555691000000102', '753591000000100', '753511000000109', '581521000000109', '407608005', '409301000000104', '429371000000100', '128570000', '536981000000104', '233561000000100', '258851000000109', '517411000000105', '668801000000103', '253662002', '728161000000107', '894601000000102', '208101000000107', '726691000000105', '373144005', '253561003', '471711000000109', '526721000000100', '89781000000102', '728271000000107', '320351000000105', '286741000000108', '1086811000000105', '726921000000103', '374251000000106', '526781000000104', '253372007', '537001000000108', '1051071000000107', '811711000000107', '204310005', '853571000000102', '22111008', '727961000000109', '292721000000103', '728001000000107', '87064008', '603911000000102', '2180001', '175062004', '612101000000109', '312651000000106', '537011000000105', '753481000000103', '253701002', '728201000000104', '632061000000101', '834741000000101', '727061000000105', '603891000000100', '516971000000108', '831781000000107', '516961000000101', '414541000000105', '586361000000104', '605041000000104', '601601000000109', '728041000000105', '657511000000105', '562851000000107', '538891000000107', '248501000000107', '359620001', '608851000000103', '1720006', '259011000000109', '627871000000102', '713221000000107', '492671000000107', '643871000000107', '885311000000107', '569971000000103', '589951000000105', '442521000000100', '757101000000106', '727051000000107', '614071000000107', '361215006', '418901000000107', '771581000000100', '726471000000109', '726461000000102', '728351000000106', '654291000000107', '274671000000107', '251651000000102', '38842003', '595931000000100', '728671000000105', '781061000000106', '807951000000103', '712411000000108', '834611000000108', '847051000000107', '11511004', '77737007', '726551000000101', '1088021000000109', '752481000000106', '253709000', '258991000000107', '727841000000104', '507031000000105', '207253004', '8957000', '614131000000109', '175011002', '831691000000102', '70390005', '728321000000101', '763651000000104', '539031000000107', '402241000000101', '728121000000104', '249301000000107', '287287007', '251581000000102', '753571000000104', '728681000000109', '728011000000109', '369061000000108', '619801000000107', '307765003', '586391000000105', '1033161000000102', '749821000000107', '727861000000103', '224151000000107', '169761000000104', '92520003', '374241000000108', '38811000000103', '810691000000104', '727511000000103', '625741000000106', '488621000000105', '501871000000106', '683171000000106', '499461000000105', '586331000000109', '233671000000108', '461582000', '451021000000103', '214621000000103', '776261000000102', '939891000000107', '259791000000102', '727881000000107', '645721000000102', '629861000000100', '727851000000101', '750131000000104', '728591000000108', '443751000000108', '753541000000105', '630501000000105', '472770005', '654281000000105', '236701000000102', '709801000000103', '727691000000108', '516991000000107', '727131000000101', '726561000000105', '594421000000105', '917871000000108', '727581000000105', '258861000000107', '538591000000105', '750151000000106', '516981000000105', '233681000000105', '812011000000100', '259101000000108', '727141000000105', '232911000000102', '728281000000100', '188151000000107', '851531000000107', '517431000000102', '38791000000104', '765651000000100', '274951000000109', '390661000000109', '727681000000106', '772261000000103', '398561000000108', '526801000000103', '726651000000102', '657531000000102', '727551000000104', '233998004', '471531000000105', '259111000000105', '727791000000101', '89801000000101', '175033000', '175081007', '237271000000101', '692841000000100', '237291000000102', '684201000000105', '596331000000108', '728811000000101', '753491000000101', '175025001', '831761000000103', '259081000000102', '553051000000104', '727251000000102', '727721000000104', '251691000000105', '274651000000103', '207254005', '632031000000106', '728301000000105', '499471000000103', '728131000000102', '707421000000103', '33622007', '182131000000102', '37171002', '727611000000104', '251661000000104', '726951000000108', '753581000000102', '84537008', '465021000000107', '198868006', '676121000000106', '175008003', '627251000000105', '208051000000107', '538881000000105', '427608000', '211371000000105', '728411000000103', '253702009', '175030002', '259801000000103', '175007008', '589921000000100', '753231000000103', '599281000000106', '214151000000100', '593631000000107', '639641000000102', '251671000000106', '727561000000101', '532881000000107', '208091000000104', '207553000', '34644005', '412781000000102', '726971000000104', '512751000000100', '727031000000100', '286948009', '65181009', '259351000000106', '471851000000100', '728171000000100', '258601000000109', '727671000000109', '599261000000102', '915581000000100', '466061000000109', '463141000000100', '259341000000108', '728871000000106', '727651000000100', '526731000000103', '440211000000107', '407606009', '728831000000109', '195547003', '831771000000105', '274631000000105', '727211000000101', '595991000000104', '810701000000104', '614141000000100', '713201000000103', '492061000000108', '636881000000100', '752211000000103', '194815007', '727121000000103', '492041000000107', '62675008', '417023008', '491591000000102', '224101000000106', '562871000000103', '639401000000103', '233651000000104', '232901000000104', '536991000000102', '596021000000109', '727501000000100', '727731000000102', '224311000000105', '814001000000106', '582911000000104', '521701000000100', '492761000000105', '443284001', '259061000000106', '286731000000104', '236711000000100', '198731000000102', '208061000000105', '778201000000100', '728311000000107', '175024002', '381171000000101', '373139003', '841991000000106', '258591000000103', '526791000000102', '251951000000108', '581531000000106', '41702007', '586351000000102', '20606002', '727591000000107'] |
| **Chronic Respiratory Disease** | ['196133001', '776981000000103', '711379004', '213223003', '196135008', '22607003', '233675009', '716241000000106', '68328006', '23315001', '425748003', '195953005', '233769008', '63480004', '36696005', '196190001', '88039007', '67569000', '196049002', '293991000000106', '86092005', '783631000000109', '196021009', '85407005', '198901000000105', '187233002', '90117007', '13394002', '195989002', '7063008', '47386001', '87433001', '195951007', '233724002', '233725001', '105977003', '866901000000103', '195957006', '68333005', '86555001', '427123006', '233661002', '313296004', '25897000', '62371005', '115666004', '51577008', '367542003', '195984007', '51615001', '233700005', '44274007', '820341000000109', '233759002', '29422001', '90610005', '233733000', '426705001', '233673002', '195959009', '473202005', '790301000000101', '10692761000119107', '196138005', '196001008', '266368002', '13645005', '233758005', '716281000000103', '195963002', '233633008', '233694004', '26427008', '1066841000000108', '233749003', '4981000', '232658009', '111292008', '233738009', '235978006', '233692000', '196027008', '236302005', '47515009', '276537001', '52333004', '361195001', '718241000000107', '195985008', '69339004', '789661000000102', '857811000000102', '233763009', '1259003', '233774000', '16003001', '37471005', '266356006', '233761006', '24369008', '427022004', '526091000000100', '73144008', '233701009', '18690003', '40122008', '196017002', '233756009', '266404004', '414087000', '313297008', '40100001', '16838000', '398726004', '371611000000107', '84409004', '713731000000102', '233628009', '857661000000104', '390941006', '196047000', '10713006', '267276009', '266366003', '232657004', '811961000000106', '87909002', '515631000000107', '77690003', '1066281000000104', '827571000000106', '383611000000102', '805261000000101', '48188009', '233771008', '42680007', '10785007', '7343008', '277656005', '275503004', '736283006', '375911000000102', '233631005', '716358000', '18121009', '428173007', '45145000', '17996008', '192658007', '190909002', '90623003', '404807005', '196046009', '38729007', '162974009', '277486008', '8247009', '196026004', '857791000000103', '704123000', '526071000000104', '196052005', '401184000', '85761009', '390891009', '408501008', '805002', '190905008', '233672007', '233727009', '78723001', '859041000000103', '233674008', '73448002', '196053000', '232660006', '233696002', '233751004', '86204009', '61937009', '233768000', '79958002', '20322005', '716901000000101', '7548000', '760601000000107', '23022004', '939431000000108', '12295008', '394702007', '46847001', '195958001', '51277007', '719218000', '233764003', '233755008', '85438006', '233659006', '195990006', '64631008', '74417001', '196137000', '233726000', '198411000000102', '80825009', '196019004', '233702002', '58691003', '285381006', '196136009', '445090007', '81423003', '42402006', '44165003', '394703002', '129451001', '233698001', '233757000', '66987001', '12088005', '700250006', '233667003', '741056003', '13151001', '760621000000103', '185086009', '233713004', '205237003', '39871006', '360470001', '8549006', '67242002', '33325001', '233664005', '14700006', '233730002', '77593006', '266355005', '50589003', '196051003', '233770009', '270473001', '426896000', '233750003', '375851000000108', '847091000000104', '1066851000000106', '74015002', '196125002', '398640008', '233634002', '429332008', '198401000000104', '48347002', '233632003', '233615002', '155597006', '49840000', '49691004', '233772001', '717521000000104', '233699009', '56968009', '810951000000101', '848431000000106', '89549007', '135836000', '232659001', '233630006', '892321000000109', '1066231000000103', '17385007', '233762004', '44549008', '401185004', '71193007', '233753001', '47895001', '7457009', '233728004', '723245007', '120639003', '50076003', '52571006', '233697006', '16623004', '196028003', '84004001', '19274004', '716311000000100', '233695003', '438773007', '717021000000106', '233666007', '233665006', '233677001', '233629001', '413845009', '233663004', '704121003', '40527005', '1066821000000101', '277485007', '361194002', '170628003', '86638007', '61233003', '599006', '233767005', '897311000000101', '715069001', '826111000000109', '233748006', '233703007', '233627004', '313299006', '1092361000119109', '51068008', '86649001', '46207001', '408681003', '427046006', '240387006', '427445002', '129452008', '41381004', '233620002', '40640008', '763888005', '707507003', '19076009', '200049006', '196040003', '278484009', '420544002', '426696003', '88687001', '196152007', '53084003', '59940009', '87153008', '59475000', '301004001', '23958009', '366971000119100', '707508008', '725052002', '427670006', '102361000119104', '233604007', '10625271000119109', '124691000119101', '128711000119106', '233613009', '70756004', '441658007', '71727007', '196151000', '10625431000119105', '724500003', '34004002', '10625671000119106', '57463004', '28791000119105', '195886008', '142931000119100', '707766007', '28295001', '83608006', '16846004', '277869007', '10625711000119105', '425996009', '181007', '195888009', '430395005', '707551007', '301002002', '233609002', '445096001', '206286008', '52500008', '31898008', '328661000119108', '186195008', '39172002', '200050006', '58890000', '46970008', '725415009', '10625111000119106', '716088000', '714203003', '722913002', '721095007', '425464007', '7678002', '417688002', '415125002', '64880000', '195896004', '737181009', '409664000', '233607000', '12310001', '59773008', '276693005', '196112005', '236793007', '10625151000119107', '426437004', '1092951000119106', '195902009', '72854003', '416916004', '312342009', '720469008', '64917006', '206289001', '276695003', '37711000', '408680002', '15708009', '724499007', '415126001', '75426006', '60805002', '44547005', '195900001', '45556008', '233617005', '301001009', '240391001', '78895009', '10625351000119105', '3487004', '12181002', '233731003', '10624991000119103', '34020007', '1033111000000104', '63607006', '196034005', '233619008', '708537005', '240741002', '737183007', '22343003', '733453005', '233623000', '385093006', '10625591000119108', '737180005', '71926009', '735465007', '195911009', '733051000', '213152006', '191727003', '89687005', '11944003', '10625751000119106', '106001000119101', '233622005', '308906005', '430969000', '10625471000119108', '10672271000119100', '186177005', '713525001', '83271005', '45312009', '123587001', '762271004', '276692000', '10625231000119106', '206283000', '64030005', '186203002', '10625071000119104', '722912007', '72656004', '233618000', '186175002', '233624006', '441590008', '707553005', '143111000119103', '187196002', '42004004', '195878008', '233608005', '186204008', '80602006', '233606009', '319841000119107', '55679008', '713544008', '233716007', '80614003', '196032009', '123588006', '123590007', '66110007', '31561003', '206287004', '2523007', '196035006', '83457000', '57702005', '2912004', '65141002', '89099002', '84753008', '442094008', '86680006', '407671000', '707552000', '67525007', '198014000', '762269004', '233714005', '10625831000119107', '421508002', '10625311000119109', '240629003', '38976008', '471272001', '47082005', '240871004', '408679000', '85469005', '18354001', '72270005', '707373004', '57686001', '10625391000119100', '10613001', '7361000175106', '401000119107', '278516003', '236807008', '765275003', '314978007', '240742009', '396285007', '708026002', '708030004', '86294001', '32139003', '10692721000119102', '12571000132104', '762270003', '708031000', '81554001', '195909000', '10625191000119102', '196115007', '1751000119100', '196033004', '430476004', '233715006', '81164001', '736056000', '40786001', '123589003', '10625511000119104', '699014000', '206284006', '427121008', '66429007', '724498004', '38699009', '111900000', '233717003', '446946005', '195904005', '200052003', '32204007', '425793006', '41269000', '187052004', '35031000119100', '441942006', '447006007', '206285007', '713526000', '233610007', '233621003', '716198008', '266370006', '371072008', '707433009', '302913000', '10625631000119108', '426853005', '301000005', '80003002', '240747003', '700252003', '31920006', '429271009', '266350000', '697923008', '32544004', '86263001', '64479007', '735466008', '47938003', '713084008', '195908008', '442025000', '366981000119102', '64667001', '41997000', '737182002', '438764004', '70036007', '60485005', '13217005', '76090006', '19325002', '233691007', '421671002', '1087061000119106', '405569006', '422588002', '233760007', '75570004', '10625031000119102', '421047005', '409665004', '22754005', '425558002', '293241000119100', '40218008', '67782005', '41207000', '61884008', '28122003', '277844007', '38534008', '707449006', '700251005', '154283005', '196037003', '1240551000000105', '35339003', '51530003', '33548005', '50804000', '200051005', '64936001', '704120002', '240635003', '420787001', '195889001', '68409003', '16311000119108', '707503004', '26511004', '195881003', '196153002', '445378003', '186193001', '396286008', '300999006', '233625007', '12381000132107', '704345008', '707435002', '1082721000119101', '737184001', '196116008', '64703005', '301003007', '123591006', '35037009', '186194007', '720401009', '233614003', '428697002', '10625551000119103', '32286006', '700249006', '207331000000102', '207321000000104', '169631000000104', '649131000000101', '432151000000100', '226461000000108', '313433007', '427069009', '414882007', '487261000000103', '564541000000102', '456031000000108', '890871000000103', '174221000000108', '432461000000108', '781661000000103', '64912000', '644271000000105', '778131000000103', '811971000000104', '523671000000103', '195738009', '626081000000109', '414341000000100', '186726005', '57716000', '88693009', '579411000000108', '213431000000100', '1033131000000107', '566801000000107', '192681000000108', '412776001', '419861000000108', '823141000000101', '21072009', '834861000000109', '827581000000108', '939441000000104', '375871000000104', '535011000000101', '383621000000108', '45157009', '196038008', '195920000', '42964004', '718251000000105', '944961000000107', '174231000000105', '454431000000109', '420245002', '47839005', '16810008', '820351000000107', '1033121000000105', '233689004', '174211000000102', '601611000000106', '111282000', '807871000000104', '419331000000108', '83316006', '832441000000100', '627021000000105', '621061000000108', '325371000000107', '389077007', '169651000000106', '866911000000101', '579421000000102', '526101000000108', '685151000000101', '429381000000103', '789761000000107', '250521000000103', '554071000000106', '31886003', '84234002', '397991000000107', '807851000000108', '233723008', '312403005', '526111000000105', '670641000000109', '1033141000000103', '339601000000108', '375921000000108', '948111000000104', '604421000000104', '944971000000100', '526211000000102', '848441000000102', '113071000000105', '1761000119103', '397190009', '834851000000106', '186191004', '213421000000102', '122971000000108', '649171000000104', '294011000000104', '586721000000108', '807391000000107', '707021000000109', '686551000000103', '57086000', '37721008', '645421000000106', '760611000000109', '707001000000100', '113061000000103', '338131000000101', '441141000000108', '113051000000101', '204574007', '616161000000107', '823151000000103', '169761000000104', '169641000000108', '60696005', '789751000000109', '656891000000102', '789671000000109', '540151000000108', '540141000000105', '807651000000109', '468671000000107', '400631000000109', '649141000000105', '781591000000108', '568721000000106', '390151000000103', '528511000000103', '626071000000107', '276694004', '658391000000105', '123620007', '240311002', '213401000000106', '523661000000105', '389075004', |

| | |
|---|---|
| | '187042003', '213411000000108', '834491000000101', '111885006', '412891000000108', '649201000000103', '611551000000109', '491091000000105', '123592004', '28168000', '707011000000103', '74455006', '897321000000107', '192671000000106', '68759006', '196161007', '857671000000106', '412775002', '253151000000106', '689401000000108', '515641000000103', '195905006', '781651000000101', '338121000000103', '325401000000109', '286451000000100', '645401000000102', '707891000000102', '630531000000104', '579661000000108', '60837001', '195882005', '706991000000106', '948121000000105', '17365008', '255621000000109', '555141000000100', '915671000000101', '564621000000105', '397981000000105', '478351000000102', '807891000000100', '601591000000103', '586341000000100', '781711000000106', '129458007', '469801000000105', '294001000000101', '255641000000102', '113031000000108', '89991000000102', '471471000000106', '617941000000107', '391131001', '187061004', '564611000000104', '636501000000107', '113041000000104', '338241000000100', '390971000000109', '790311000000104', '375861000000106', '81638006', '204572006', '368861000000109', '442341000000102', '810721000000108', '233693005', '783641000000100', '375931000000105', '255631000000106', '826121000000103', '312602005', '857801000000104', '554081000000108', '196009005', '812311000000102', '831811000000105', '292571000000107', '652331000000107', '89087009', '409081000000107', '654461000000102', '409811000000108', '832111000000108', '50648007', '15341009', '807861000000106', '396284006', '760631000000101', '57541005', '550501000000102', '413846005', '451271000000107', '649151000000108', '859051000000100'] |
| **Heart Failure** | ['101281000119107', '10335000', '10633002', '111283005', '120851000119104', '120861000119102', '120881000119106', '120891000119109', '128404006', '13839000', '153931000119109', '153941000119100', '153951000119103', '15629541000119106', '15629591000119103', '15629741000119102', '15781000119107', '15964661000119102', '15964701000119109', '161505003', '16838951000119100', '194767001', '194779001', '194781004', '195111005', '195112003', '195114002', '206586007', '23341000119109', '233924009', '236003008', '25544003', '275514001', '309634009', '314206003', '359617009', '360371003', '364006', '367363000', '371862006', '40541001', '410431009', '417996009', '418304008', '42343007', '424404003', '426012001', '426263006', '426611007', '43736008', '44088000', '441481004', '441530006', '44313006', '443253003', '443254009', '443343001', '443344007', '446221000', '45650007', '46113002', '471880001', '48447003', '49584005', '5148006', '5375005', '56675007', '66989003', '67431000119105', '67441000119101', '698296002', '698594003', '703272007', '703273002', '703274008', '703275009', '703276005', '717840005', '71892000', '722095005', '72481000119103', '74960003', '78862003', '788950000', '79955004', '80479009', '82523003', '83105008', '83291003', '84114007', '85232009', '871617000', '88805009', '89555002', '898208007', '90727007', '92506005', '96311000119109'] |
| **Cancer** | ['1014611000000103', '1025781000000106', '103682005', '103685007', '103686008', '103688009', '103689001', '103690005', '103691009', '105111000119109', '1051141000000100', '105121000119102', '1054001000000101', '10639003', '10708511000119108', '10745291000119103', '10749871000119100', '107581000119103', '107591000119100', '107601000119107', '107691000119101', '107751000119102', '107771000119106', '107781000119109', '107791000119107', '1078881000119102', '1078901000119100', '1078931000119107', '1078961000119104', '1079091000119100', '1079101000119105', '1079111000119108', '1079131000119103', '1079151000119109', '1079161000119106', '1079171000119100', '1079191000119104', '1080111000119108', '1080151000119109', '1080161000119106', '1080191000119104', '1080231000119108', '1080241000119104', '1080261000119100', '1080341000119105', '1080941000119109', '1080981000119104', '1081021000119109', '1081551000119106', '1081561000119108', '1081711000119104', '1081751000119103', '1081781000119105', '1081811000119107', '1082071000119102', '1082101000119106', '1082191000119100', '1082221000119106', '1082281000119105', '1082291000119108', '1082301000119109', '1082311000119107', '1090081000000106', '1090091000000108', '1090101000000100', '1090121000000109', '1090131000000106', '1090141000000102', '1090151000000104', '1090161000000101', '1090171000000108', '1090181000000105', '1090191000000107', '1090201000000109', '1090211000000106', '1090221000000100', '1090231000000103', '1090241000000107', '1090251000000105', '1090261000000108', '1090271000000101', '1090281000000104', '1090291000000102', '1090301000000103', '1090311000000101', '1090321000000107', '1090821000000101', '1090871000000102', '1090881000000100', '1090891000000103', '1090901000000102', '1090911000000100', '1090921000000106', '1090931000000108', '1090941000000104', '1090951000000101', '1090961000000103', '1090971000000105', '1090981000000107', '1090991000000109', '1091861000000100', '1091891000000106', '1091921000000103', '109264009', '109267002', '109347009', '109348004', '109349007', '109356001', '109357005', '109358000', '109359008', '109366009', '109367000', '109368005', '109369002', '109370001', '109371002', '109374005', '109378008', '109383000', '109384006', '109385007', '109386008', '109388009', '109389001', '109390005', '109391009', '109392002', '109821008', '109822001', '109823006', '109824000', '109828002', '109830000', '109831001', '109832008', '109833003', '109834009', '109835005', '109836006', '109837002', '109838007', '109839004', '109840002', '109841003', '109842005', '109843000', '109844006', '109847004', '109848009', '109851002', '109853004', '109854005', '109855006', '109856007', '109858008', '109870007', '109874003', '109875002', '109876001', '109878000', '109879008', '109880006', '109882003', '109885001', '109886000', '109887009', '1098981000119101', '109911004', '109912006', '109915008', '109918005', '109919002', '109921007', '109923005', '109925003', '109927006', '109931000', '109933002', '109937001', '109939003', '109941002', '109943004', '109945006', '109947003', '109948008', '109951001', '109962001', '109964000', '109965004', '109966003', '109967007', '109968002', '109969005', '109970006', '109971005', '109972003', '109975001', '109976000', '109977009', '109978004', '109979007', '109980005', '109982002', '109984001', '109985000', '109988003', '109989006', '109991003', '109992005', '109994006', '109995007', '109996008', '109998009', '110000005', '110002002', '110004001', '110005000', '110006004', '110007008', '110013004', '11010461000119101', '110457005', '110458000', '110459008', '110460003', '11073003', '112241002', '112677002', '112679004', '112683004', '112684005', '112685006', '112687003', '115223002', '115232000', '115241005', '115244002', '115245001', '115246000', '116381000119105', '11671000', '116811000119106', '116821000119104', '118599009', '118600007', '118601006', '118602004', '118605002', '118606001', '118607005', '118608000', '118609008', '118610003', '118611004', '118612006', '118613001', '118614007', '118615008', '118617000', '118618005', '119247004', '119248009', '12060004', '12240951000119107', '12240991000119102', '12246561000119101', '12301000132103', '12302002', '12311000132101', '12323008', '123313007', '12341000132100', '12351000132102', '12354007', '123631000119103', '123842006', '123845008', '12622007', '126885006', '12690005', '126906006', '127066000', '127070008', '127216000', '127220001', '127221002', '127225006', '127580003', '128041000119107', '128087008', '128462008', '128465005', '128466006', '128632008', '128645007', '128726006', '128734000', '128735004', '128749008', '128750008', '128753005', '128756002', '128770002', '128771003', '128772005', '128773000', '128774006', '128775007', '128776008', '128783001', '128791005', '128792003', '128794002', '128796000', '128798004', '128799007', '128800006', '128801005', '128802003', '128803008', '128805001', '128806000', '128807009', '128808004', '128812005', '128813000', '128814006', '128815007', '128816008', '128817004', '128818009', '128819001', '128820007', '128821006', '128822004', '128823009', '128824003', '128825002', '128826001', '128827005', '128828000', '128829008', '128830003', '128831004', '128832006', '128833001', '128834007', '128841001', '128858006', '128874001', '128875000', '128883006', '128884000', '128900005', '128910001', '128911002', '128912009', '128913004', '128916007', '128918008', '128919000', '128920006', '128921005', '128922003', '128923008', '128924002', '128929007', '128930002', '128931003', '128932005', '128933000', '128934006', '128935007', '129000002', '12943006', '13048006', '13238004', '13351431000119102', '133531000119104', '133751000119102', '133871000119103', '133881000119100', '134160009', '134166003', '134174002', '134176000', '134177009', '134178004', '134179007', '134180005', '134181009', '134182002', '134312002', '134332001', '134421000', '13583002', '13875003', '14269005', '14317002', '1443001', '14494009', '14537002', '145831000119103', '147101000119108', '147131000119101', '14990007', '15118008', '15176003', '1556006', '15619004', '15635721000119108', '15635801000119106', '15674004', '15949004', '15956181000119102', '15956341000119105', '15956381000119100', '16034002', '16090008', '16260631000119101', '1651000119109', '1661000119106', '166851000000104', '1671000119100', '167934009', '1681000119102', '16893006', '1691000119104', '16974005', '169931000000107', '1701000119104', '170941000000100', '170961000000104', '170981000000108', '17264009', '17302008', '174581000000106', '175601000000101', '175621000000105', '175641000000103', '177281002', '17788007', '18105004', '18121000119104', '183649005', '18450009', '184741000119107', '184781000119102', '184871000119108', '184881000119106', '184891000119109', '18588008', '186709004', '186722007', '186723002', '186725009', '187600004', '187601000', '187604008', '187606005', '187607001', '187608006', '187613005', '187614004', '187622006', '187624007', '187631006', '187633009', '187634003', '187635002', '187637005', '187640005', '187641009', '187644001', '187652003', '187653008', '187658004', '187659007', '187660002', '187661003', '187662005', '187666008', '187675005', '187681002', '187682009', '187683004', '187685006', '187688008', '187692001', '187693006', '187694000', '187697007', '187698002', '187700006', '187701005', '187702003', '187708004', '187709007', '187716008', '187722004', '187723009', '187724003', '187725002', '187726001', '187727005', '187732006', '187733001', '187734007', '187736009', '187737000', '187738005', '187740000', '187741001', '187742008', '187752007', '187757001', '187760008', '187767006', '187769009', '187773007', '187775000', '187776004', '187777008', '187784000', '187786003', '187791002', '187792009', '187793004', '187794005', '187798008', '187801002', '187803004', '187804005', '187807003', '187808008', '187809000', '187810005', '187811009', '187814001', '187821001', '187822008', '187824009', '187828007', '187829004', '187830009', '187831008', '187833006', '187834000', '187835004', '187836003', '187838002', '187841006', '187842004', '187843009', '187844003', '187845002', '187846001', '187852000', '187853005', '187854004', '187857006', '187861000', '187862007', '187864008', '187865009', '187866005', '187868006', '187869003', '187870002', '187881004', '187894002', '187895001', '187900002', '187903000', '187906008', '187916000', '187917009', '187918004', '187920001', '187925006', '187926007', '187927003', '187929000', '187932002', '187937008', '187938003', '187939006', '187940008', '187941007', '187942000', '187943005', '187944004', '187945003', '187946002', '187947006', '187948001', '187949009', '187950009', '187952001', '187956003', '187957007', '187959005', '187967002', '187968007', '187969004', '187970003', '187972006', '187973001', '187974007', '187975008', '187976009', '187977000', '187978005', '187979002', '187987001', '187988006', '187989003', '187991006', '187992004', '187993009', '187994003', '187995002', '187996001', '187997005', '187999008', '188000002', '188001003', '188002005', '188003000', '188004006', '188005007', '188006008', '188009001', '188010006', '188013008', '188015001', '188016000', '188017009', '188019007', '188020001', '188021002', '188022009', '188023004', '188029000', '188030005', '188032002', '188033007', '188034001', '188038003', '188040008', '188042000', '188044004', '188045003', '188046002', '188049009', '188050009', '188051008', '188052001', '188054000', '188055004', '188060000', '188061001', '188062008', '188063003', '188064009', '188065005', '188067002', '188068007', '188069004', '188070003', '188071004', '188072006', '188073001', '188074007', '188075008', '188076009', '188077000', '188085009', '188089003', '188090007', '188091006', '188095002', '188099008', '188100000', '188102008', '188103003', '188107002', '188110009', '188119005', '188121000', '188122007', '188125009', '188127001', '188132000', '188133005', '188135003', '188138001', '188147009', '188151006', '188152004', '188153009', '188154003', '188155002', '188156001', '188157005', '188159008', '188163001', '188168005', '188176007', '188177003', '188180002', '188183000', '188184006', '188188009', '188189001', '188190005', '188191009', '188192002', '188193007', '188195000', '188196004', '188198003', '188204000', '188208002', '188209005', '188211001', '188219004', '188220005', '188230001', '188234005', '188235006', '188239000', '188240003', '188241004', '188242006', '188243001', '188244007', '188245008', '188247000', '188250002', '188251003', '188252005', '188253000', '188256008', '188261005', '188263008', '188264002', '188265001', '188266000', '188268004', '188269007', '188272000', '188273005', '188274004', '188280007', '188281006', '188282004', '188283009', '188285002', '188286001', '188287005', '188289008', '188290004', '188292007', '188293002', '188295009', '188296005', '188297001', '188298006', '188301005', '188302003', '188307009', '188308004', '188312005', '188313000', '188315007', '188317004', '188318009', '188319001', '188321006', '188322004', '188323009', '188324003', '188325002', '188326001', '188327005', '188339002', '188340000', '188353002', '188361007', '188366002', '1884006', '188434008', '188445006', '188454009', '188458007', '188459004', '188462001', '188469005', '188471005', '188478004', '188487008', '188489006', '188492005', '188493000', '188498009', '188500005', '188501009', '188502002', '188503007', '188504001', '188505000', '188506004', '188507008', '188510001', '188511002', '188512009', '188513004', '188514005', '188515006', '188516007', '188517003', '188521005', '188523008', '188524002', '188526000', '188527009', '188528004', '188529007', '188531003', '188534006', '188536008', '188537004', '188538009', '18854008', '188541000', '188544008', '188547001', '188548006', '188551004', '188554007', '188558005', '188559002', '188561006', '188562004', '188565002', '188566001', '188567005', '188568000', '188569008', '188570009', '188572001', '188575004', '188576003', '188577007', '188578002', '188579005', '188580008', '188582000', '188585003', '188586002', '188587006', '188589009', '188590000', '188591001', '188592008', '188593003', '188609000', '18861007', '188612002', '188613007', '188619006', '188623003', '188624009', '188627002', '188630009', '188631008', '188632001', '188633006', '188634000', '188635004', '188636003', '188637007', '188640007', '188641006', '188642004', '188645002', '188648000', '188649008', '188651007', '188656002', '188657006', '188658001', '188660004', '188662007', '188663002', '188664008', '188665009', '188666005', '188667001', '188668006', '188669003', '188672005', '188674006', '188675007', '188676008', '188678009', '188679001', '188691005', '188718006', '188725004', '188726003', '188728002', '188729005', '188732008', '188733003', '188734009', '188736006', '188737002', '188738007', '188741003', '188744006', '188745007', '188746008', '188748009', '188754005', '188756007', '188759000', '188768003', '188770007', '189509003', '189510008', '189551005', '189552003', '189553008', '189555001', '189562005', '189563000', '189573003', '189576006', '189584005', '189585006', '189587003', '189590009', '189593006', '189596003', '189600008', '189601007', '189603005', '189605003', '189606002', '189607006', '189608001', '189609009', '189611000', '189612007', '189615009', '189619003', '189621008', '189623006', '189624000', '189626003', '189627007', '189628002', '189629005', '189630000', '189631001', '189632008', '189633003', '189634009', '189636006', '189640002', '189642005', '189643000', '189649001', '189652009', '189659000', '189661009', '189662002', '189663007', '189666004', '189667008', '189669006', '189670007', '189671006', '189672004', '189673009', '189679008', '189681005', '189682003', '189685001', '189687009', '189692006', '189699002', '189700001', '189702009', '189709000', '189711009', '189713007', '189715000', '189747005', '189754004', '189756002', '189757006', '189761000', '189764008', '189765009', '189777004', '189779001', '189790006', '189796000', '189797009', '189802003', '189805001', '189806000', '189808004', '189813000', '189815007', '189818009', '189819001', '189826001', '189830003', '189832006', '189835008', '189837000', '189838005', '189840000', '189841001', '189843003', '189849004', '189850004', '189857001', '189871009', '189881008', '189882001', '189884000', '189885004', '189886003', '189889005', '189912009', '189916007', '189919000', '189920006', '189921005', '189922003', '189923008', '189926000', '189927009', '189928004', '189930002', '189932005', '189933000', '189934006', '189935007', '189939001', '189957000', '189960007', '189961006', '189962004', '189963009', '189966001', '189967005', '189968000', '189969008', '189970009', '189971008', '189972001', '189973006', '189974000', '189975004', '189976003', '189977007', '189978002', '189980008', '189981007', '189982000', '189984004', '189985003', '189988001', '189989009', '189990000', '189991001', '189993003', '189995005', '189996006', '189997002', '189998007', '189999004', '190001002', '190002009', '190004005', '190006007', '190007003', '190008008', '190009000', '190011009', '190012002', '190013007', '190016004', '190018003', '190019006', '190022008', '190023003', '190024009', '190025005', '190026006', '190028007', '190029004', '190030009', '190031008', '190032001', '190033006', '190034000', '190035004', '190036003', '190038002', '190039005', '190040007', '190043009', '190045002', '190046001', '190047005', '190048000', '190049008', '190050008', '190051007', '190052000', '190053005', '190054004', '190055003', '190056002', '190057006', '190063002', '190067001', '190069003', '190070002', '190073000', '190075007', '190079001', '190080003', '190086009', '190090006', '190091005', '190096000', '190098004', '190100004', '190102007', '190103002', '190105009', '190107001', '190108006', '190110008', '190111007', '190112000', '190115003', '190120003', '190122006', '190123001', '190127000', '190131006', '190132004', '190133009', '190135002', '190137005', '190138000', '190139008', '190140005', '190141009', '190143007', '190144001', '190145000', '190146004', '190147008', '190148003', '190149006', '190151005', '190162005', '190164006', '190165007', '190166008', '190167004', '190222001', '190229005', '190817009', '190818004', '190955000', '191333009', '19134004', '192581000000107', '1929004', '19340000', '19467007', '19756007', '198651000000100', '19897006', '19906005', '20224008', '203436008', '20380000', '20447006', '20667008', '208041000119100', '2092003', '20955008', '21008007', '21165006', '21326004', '213981000000105', '2142002', '21589007', '21708004', '21851000119103', '21912003', '21964009', '21968007', '22197008', '2221008', '22217002', '2227007', '22331004', '22694002', '230156002', '231829006', '231831002', '231832009', '231833004', '231834005', '231835006', '232075002', '233693005', '233940007', '23444003', '234577004', '234941000119100', '235965006', '235966007', '236512004', '236513009', '236811000119101', '237252008', '23746000', '237759003', '237833006', '237865009', '238636003', '238637007', '239297008', '239940004', '24007003', '240163000', '240531002', '24072005', '24111000119105', '241861008', '2424003', '242862004', '242951000119108', '24604009', '24653009', '248455008', '25050002', '252999006', '253000007', '253001006', '253003009', '253014007', '253017000', '253018005', '253026002', '253032007', '253036005', '253039003', '253042009', '253046007', '253056006', '253063006', '253071005', '253072003', '253074002', '253075001', '253076000', '253077009', '253092005', '253093000', '253094006', '253096008', '25370001', '254042000', '254383006', '254389005', '254390001', '254393004', '254398008', '254402004', '254404003', '254408000', '254412006', '254417000', '254423005', '254424004', '254425003', '254427006', '254431000', '254434008', '254435009', '254436005', '254437001', '254441002', '254445006', '254450000', '254454009', '254457002', '254459004', '254462001', '254465004', '254466003', '254474002', '254478004', '254481009', '254484001', '254503007', '254509006', '254513004', '254517003', '254520006', '254523008', '254526000', '254530002', '254535007', '254539001', '254543002', '254547001', '254549003', '254551004', '254553001', '254555008', '254557000', '254559002', '254561006', '254563009', '254564003', '254567005', '254570009', '254582000', '254586002', '254601002', '254609000', '254611009', '254612002', '254619006', '254620000', '254622008', '254625005', '254626006', '254627002', '254628007', '254629004', '254630009', '254631008', '254632001', '254633006', '254634000', '254635004', '254637007', '254638002', '254639005', '254645002', '254650008', '254651007', '254652000', '254653005', '254654004', '254655003', '254701007', '254702000', '254703005', '254707006', '254708001', '254709009', '254710004', '254711000', '254712007', '254713002', '254714008', '254726003', '254727007', '254730000', '254731001', '254732008', '254733003', '254734009', '254748009', '254764001', '254771006', '254792006', '254794007', '254797000', '254800003', '254820002', '254824006', '254828009', '254829001', '254837009', '254838004', '254839007', '254840009', '254841008', '254843006', '254844000', '254849005', '254850005', '254851009', '254852002', '254853007', '254855000', '254856004', '254860001', '254861002', '254863004', '254869000', '254870004', '254871000', '254872007', '254874008', '254876005', '254877001', '254878006', '254886006', '254887002', '254888007', '254889004', '254893005', '254895003', '254896002', '254897006', '254898001', '254900004', '254904008', '254908006', '254909003', '254912000', '254915003', '254916002', '254917006', '254918001', '254934003', '254938000', '254940005', '254948003', '254950006', '254955001', '254969001', '254972008', '254973003', '254974009', '254975005', '254980001', '254983004', '254986007', '254987003', '254988008', '254989000', '254990009', '254993006', '254994000', '254995004', '254996003', '255003007', '255004001', '255008003', '255012009', '255014005', '255015006', '255016007', '255017003', '255021005', '255028004', '255029007', '255030002', '255031003', '255032005', '255035007', '255037004', '255044008', '255052006', '255056009', '255066001', '255067005', '255068000', '255069008', '255070009', '255071008', '255072001', '255073006', '255074000', '255075004', '255077007', '255078002', '255081007', '255083005', '255084004', '255086002', '255087006', '255088001', '255090000', '255091001', '255093003', '255096006', '255100007', '255101006', '255102004', '255106001', '255107005', '255108000', '255109008', '255110003', '255111004', '255112006', '255114007', '255115008', '255116009', '255117000', '255118005', '255119002', '255121007', '255123005', '255124004', '255127006', '255128001', '255129009', '255131000', '255191003', '26019009', '26138003', '26211003', '26328002', '26888009', '269459004', '269460009', '269463006', '269464000', '269466003', '269467007', '269468002', '269469005', '269473008', '269475001', '269476000', '269478004', '269499001', '269501009', '269503007', '269506004', '269507008', '269508003', '269509006', '269510001', '269515006', '269516007', '269533000', '269543002', '269544008', '269567005', '269569008', '269578002', '269579005', '269580008', '269581007', '269616004', '269617008', '27092008', '271323007', '271385002', '271467005', '271468000', '271486002', '271568003', '271943005', '271944004', '2735009', '274084007', '274085008', '274087000', '274088005', '27461004', '274902006', '274904007', '274905008', '274907000', '275265005', '275266006', '275394001', '275395000', '275396004', '275397008', '275399006', '275419009', '275464006', '275490009', '275524009', '276419004', '276420005', '276499003', '276611000000108', '276738009', '276750003', '276751004', '276797002', '276799004', '276803003', '276804009', '276808007', '276809004', '276810009', '276811008', '276815004', '276821000', '276822007', '276826005', '276827001', '276828006', '276829003', '276836002', '276860003', '276870001', '276876007', '276952000', '276953005', '276954004', '276962007', '276975007', '277156006', '277461004', '277473004', '277474005', '277505007', '277530005', '277543005', '277545003', '277549009', '277550009', '277551008', '277567002', '277568007', '277569004', '277570003', '277571004', '277572006', '277573001', '277574007', '277575008', '277576009', '277577000', '277579002', '277580004', '277587001', '277589003', '277597005', '277600006', '277601005', '277602003', '277604002', '277608004', '277609007', '277610002', '277611003', '277612005', '277613000', '277614006', '277615007', '277616008', '277617004', '277618009', '277619001', '277622004', '277623009', '277624003', '277625002', '277626001', '277627005', '277628000', '277629008', '277632006', '277637000', '277641001', '277642008', '277643003', '277648007', '277651000', '277653002', '277654008', '277664004', '277782009', '277984002', '277985001', '277993001', '278024000', '278042005', '278043000', '278044006', '278046008', '278050001', '278051002', '278052009', '278053004', '278054005', '278055006', '278060005', '278065000', '278189009', '278190000', '278411006', '278433008', '278453007', '27849002', '278491007', '28047004', '28054005', '280959007', '281560004', '281562007', '281563002', '281564008', '281565009', '281566005', '281702006', '28173006', '28325008', '28351005', '285307007', '285308002', '285309005', '285310000', '285312008', '285420006', '285421005', '285422003', '285423008', '285424002', '285426000', '285428004', '285430002', '285432005', '28558000', '285598005', '285603002', '285604008', '285605009', '285606005', '285607001', '285608006', '285609003', '285610008', '285611007', '285612000', '285613005', '285614004', '285615003', '285616002', '285617006', '285618001', '285619009', '285631006', '285633009', '285634003', '285635002', '285637005', '285638000', '285639008', '285640005', '285641009', '285642002', '285643007', '285644001', '285645000', '285763005', '285766002', '285768001', '285769009', '285776004', '285839005', '28655007', '286887005', '286889008', '286890004', '286891000', '286892007', '286893002', '286894008', '286895009', '286896005', '286897001', '286899003', '286900008', '286902000', '28950004', '28953002', '292551000000103', '29370006', '29421000119105', '294301000000103', '294311000000101', '294321000000107', '29792007', '2985005', '300987004', '300988009', '301036008', '301756000', '302815008', '302816009', '302817000', '302835009', '302837001', '302840001', '302841002', '302842009', '302845006', '302847003', '302848008', '302849000', '302851001', '302852008', '302855005', '302856006', '30289006', '303012000', '303017006', '303055001', '303056000', '303057009', '303194003', '303201005', '30383009', '30440004', '304545002', '30546008', '306870009', '30713000', '307216009', '307219002', '307226002', '307340003', '307341004', '307502000', '307576001', '307592006', '307593001', '307599002', '307601000', '307603002', '307608006', '307609003', '307610008', '307617006', '307622006', '307623001', '307624007', '307625008', '307633009', '307634003', '307635002', '307636001', '307637005', '307646004', '307647008', '307649006', '307650006', '307651005', '307652003', '307726001', '308121000', '30924005', '309245001', '309431000000107', '30962008', '309831004', '3102004', '31047003', '310498001', '310504009', '310526005', '310599006', '31131002', '311779007', '31186001', '312104005', '312111009', '312112002', '312113007', '312114001', '312115000', '312939009', '312949007', '313248004', '313249007', '313250007', '313353007', '313354001', '313355000', '313356004', '313357008', '313417009', '313427003', '313428008', '313429000', '313430005', '313431009', '31396002', '314191009', '314408000', '314418005', '31470003', '314922006', '314923001', '314924007', '314925008', '314926009', '314927000', '314929002', '314930007', '314931006', '314934003', '314951005', '314952003', '314953008', '314954002', '314955001', '314960002', '314961003', '314962005', '314963000', '314964006', '314965007', '314966008', '314967004', '314968009', '314969001', '314970000', '314973003', '314974009', '314975005', '314976006', '314987003', '314988008', '314989000', '314990009', '314991008', '314992001', '314993006', '314994000', '314995004', '314996003', '314997007', '314998002', '314999005', '315000005', '315001009', '315002002', '315003007', '315004001', '315005000', '315006004', '315007008', '315008003', '315009006', '315058005', '31671006', '3172003', '32280000', '32456001', '32512003', '326072005', '326681000000109', '326691000000106', '326701000000106', '326711000000108', '32913002', '32968003', '33176006', '33681003', '337231000000108', '337241000000104', '337251000000101', '337261000000103', '337671000000108', '337681000000105', '337691000000107', '337701000000107', '338011000000108', '342511000000104', '342561000000102', '342571000000109', '34360000', '351461000119100', '352001000119100', '352071000119105', '352201000119105', '352231000119103', '352301000119103', '352311000119100', '35232005', '352321000119107', '35262004', '35287006', '353561000119103', '353661000119102', '353671000119108', '353741000119106', '354002', '354351000119105', '354361000119107', '354621000119105', '354641000119104', '354651000119102', '354671000119106', '354681000119109', '35562000', '35601003', '35868009', '359626007', '359631009', '359636004', '359640008', '359648001', '359777006', '359780007', '359782004', '359785002', '359987004', '360331008', '36310008', '363346000', '363348004', '363349007', '363350007', '363351006', '363352004', '363353009', '363354003', '363355002', '363357005', '363358000', '363359008', '363360003', '363363001', '363364007', '363365008', '363366009', '363367000', '363368005', '363369002', '363370001', '363372009', '363373004', '363374005', '363375006', '363376007', '363377003', '363378008', '363379000', '363380002', '363381003', '363382005', '363383000', '363384006', '363385007', '363386008', '363387004', '363388009', '363389001', '363390005', '363391009', '363392002', '363393007', '363394001', '363395000', '363396004', '363397008', '363398003', '363399006', '363400004', '363401000', '363402007', '363403002', '363404008', '363405009', '363406005', '363407001', '363408006', '363409003', '363410008', '363411007', '363412000', '363413005', '363414004', '363415003', '363416002', '363417006', '363418001', '363419009', '363420003', '363421004', '363422006', '363423001', '363424007', '363425008', '363426009', '363427000', '363428005', |

| | |
|---|---|
| | '363429002', '363430007', '363431006', '363432004', '363433009', '363434003', '363435002', '363436001', '363437005', '363438000', '363439008', '363440005', '363441009', '363443007', '363444001', '363445000', '363446004', '363447008', '363449006', '363450006', '363451005', '363452003', '363453008', '363454002', '363456000', '363457009', '363458004', '363459007', '363460002', '363461003', '363462005', '363463000', '363464006', '363465007', '363466008', '363467004', '363468009', '363469001', '363470000', '363471001', '363473003', '363474009', '363475005', '363476006', '363477002', '363478007', '363481002', '363482009', '363483004', '363484005', '363485006', '363486007', '363487003', '363488008', '363489000', '363490009', '363491008', '363492001', '363493006', '363494000', '363495004', '363496003', '363497007', '363498002', '363499005', '363500001', '363501002', '363502009', '363503004', '363504005', '363505006', '363506007', '363507003', '363508008', '363509000', '363510005', '363514001', '363515000', '363516004', '363517008', '363518003', '363745004', '36425007', '36449005', '366059005', '367051000000101', '36741007', '368671000000103', '368681000000101', '368691000000104', '369448007', '369449004', '369450004', '369451000', '369452007', '369453002', '369454008', '369455009', '369456005', '369457001', '369458006', '369459003', '369460008', '369461007', '369463005', '369464004', '369465003', '369466002', '369467006', '369468001', '369469009', '369470005', '369471009', '369472002', '369473007', '369474001', '369475000', '369476004', '369477008', '369478003', '369479006', '369480009', '369481008', '369482001', '369483006', '369484000', '369485004', '369486003', '369487007', '369488002', '369489005', '369490001', '369491002', '369492009', '369493004', '369494005', '369495006', '369496007', '369497003', '369498008', '369499000', '369500009', '369501008', '369502001', '369503006', '369504000', '369505004', '369506003', '369507007', '369508002', '369509005', '369510000', '369511001', '369512008', '369513003', '369514009', '369515005', '369516006', '369517002', '369518007', '369519004', '369520005', '369521009', '369522002', '369523007', '369524001', '369525000', '369526004', '369527008', '369528003', '369529006', '369530001', '369531002', '369532009', '369533004', '369534005', '369535006', '369536007', '369538008', '369540003', '369541004', '369542006', '369543001', '369544007', '369545008', '369546009', '369547000', '369548005', '369549002', '369550002', '369551003', '369552005', '369553000', '369554006', '369555007', '369556008', '369557004', '369558009', '369559001', '369560006', '369561005', '369562003', '369563008', '369564002', '369565001', '369566000', '369567009', '369568004', '369569007', '369570008', '369571007', '369572000', '369573005', '369574004', '369575003', '369576002', '369577006', '369578001', '369579009', '369580007', '369581006', '369582004', '369583009', '369584003', '369585002', '369586001', '369587005', '369588000', '369589008', '369590004', '369591000', '369592007', '369593002', '369594008', '369595009', '369596005', '369597001', '369598006', '369599003', '369600000', '369601001', '369602008', '369603003', '369604009', '369605005', '369606006', '369607002', '369608007', '369609004', '369610009', '369748008', '369775001', '369777009', '370511006', '37064009', '370967009', '370987005', '371012000', '371134001', '37138001', '371962007', '371963002', '371964008', '371965009', '371966005', '371967001', '371968006', '371969003', '371970002', '371971003', '371972005', '371973000', '371974006', '371975007', '371976008', '371977004', '371978009', '371979001', '371980003', '371981004', '371982006', '371983001', '371984007', '371985008', '371986009', '371987000', '371988005', '371989002', '371990006', '371991005', '371992003', '371993008', '371994002', '371995001', '371996000', '371997009', '371998004', '371999007', '372000001', '372001002', '372002009', '372003004', '372004005', '372005006', '372006007', '372007003', '372008008', '372009000', '372010005', '372011009', '372012002', '372013007', '372014001', '372015000', '372016004', '372017008', '372018003', '372019006', '372020000', '372021001', '372022008', '372023003', '372024009', '372025005', '372026006', '372027002', '372028007', '372030009', '37206003', '372062007', '372063002', '372064008', '372065009', '372087000', '372092003', '372093008', '372094002', '372095001', '372096000', '372097009', '372098004', '372099007', '372100004', '372101000', '372103002', '372104008', '372105009', '372106005', '372107001', '372108006', '372110008', '372111007', '372112000', '372113005', '372115003', '372116002', '372119009', '372120003', '372121004', '372122006', '372123001', '372124007', '372125008', '372126009', '372127000', '372128005', '372129002', '372130007', '372131006', '372133009', '372135002', '372136001', '372137005', '372138000', '372139008', '372140005', '372141009', '372142002', '372143007', '372151005', '372152003', '372156000', '372244006', '373080008', '373081007', '373082000', '373083005', '373088001', '373089009', '373090000', '373091001', '373168002', '373627005', '37373007', '373888000', '37810007', '384951004', '385377005', '385478001', '38549000', '388650006', '389216001', '389231000000105', '389331000000104', '389361000000109', '389491000000107', '38958001', '389731000000103', '389741000000107', '389751000000105', '389761000000108', '389771000000101', '389781000000104', '389831000000109', '389851000000102', '389861000000104', '389871000000106', '389881000000108', '39000009', '39005004', '390261000000107', '390271000000100', '390281000000103', '390391000000109', '390401000000107', '390651000000106', '390781000000108', '39086001', '390901000000101', '391381000000100', '391691000000109', '392061000000102', '39274007', '393563007', '394932008', '395099008', '395671000000107', '396121000000105', '396131000000107', '396161000000102', '396181000000106', '396198006', '396201000000105', '396241000000108', '396491000000102', '396611000000109', '396651000000108', '396671000000104', '396701000000100', '396711000000103', '396731000000106', '396761000000101', '396771000000108', '396781000000105', '396791000000107', '396801000000106', '396811000000108', '396821000000102', '396891002', '396892009', '397008008', '397009000', '397011009', '397015000', '397079001', '397080003', '397081004', '397121000000107', '397141000000100', '397340004', '397341000', '397342007', '397343002', '397344008', '397345009', '397346005', '397347001', '397348006', '397349003', '397350003', '397351004', '397352006', '397355008', '397356009', '397357000', '397358005', '397379005', '397400006', '397421000000102', '397431000000100', '397451000000107', '397467006', '397468001', '397469009', '397621000000100', '397631000000103', '397641000000107', '397801000000108', '39781001', '397811000000105', '39795003', '398271000000103', '398601000000108', '398623004', '398670003', '398691000000101', '398701000000101', '398903003', '39896009', '399068003', '399081000000101', '399211000000101', '399326009', '399399005', '399456004', '399475009', '399477001', '399479003', '399490008', '399491000000106', '399501000000100', '399521000', '399552004', '399590005', '399607007', '399634005', '399644007', '399648005', '399660006', '399712003', '399884001', '399887008', '399897004', '399934007', '399954008', '399968001', '399969009', '400001003', '400011005', '400057007', '400058002', '400074007', '400094003', '400122007', '400151006', '400173004', '400174005', '400175006', '400180002', '401121000000109', '401123009', '401221000000100', '401241000000107', '401561000000106', '40244008', '402494003', '402495002', '402496001', '402497005', '402498000', '402499008', '402500004', '402501000', '402502007', '402503002', '402504008', '402505009', '402506005', '402507001', '402508006', '402509003', '402510008', '402511007', '402512000', '402513005', '402514004', '402515003', '402516002', '402517006', '402518001', '402519009', '402520003', '402521004', '402522006', '402523001', '402524007', '402525008', '402526009', '402527000', '402528005', '402529002', '402530007', '402531006', '402532004', '402533009', '402534003', '402535002', '402536001', '402537005', '402538000', '402539008', '402540005', '402541009', '402542002', '402543007', '402544001', '402545000', '402546004', '402547008', '402558004', '402559007', '402560002', '402561003', '402562005', '402563000', '402564006', '402636006', '402652009', '402761000000108', '402815007', '402816008', '402817004', '402818009', '402819001', '402820007', '402871009', '402873007', '402876004', '402879006', '402880009', '402881008', '402882001', '402910001', '402911002', '402912009', '40312006', '403451000000108', '403458008', '403468003', '403711001', '403712008', '403713003', '403714009', '403729006', '403742006', '403889000', '403890009', '403891008', '403892001', '403893006', '403894000', '403895004', '403896003', '403897007', '403898002', '403899005', '403900000', '403901001', '403902008', '403903003', '403904009', '403905005', '403906006', '403909004', '403910009', '403911000000104', '403911008', '403912001', '403913006', '403914000', '403915004', '403916003', '403917007', '403918002', '403919005', '403920004', '403921000', '403922007', '403923002', '403924008', '403925009', '403926005', '403927001', '403929003', '403939009', '403940006', '403941005', '403942003', '403943008', '403944002', '403946000', '403947009', '403948004', '403949007', '403950007', '403951006', '403954003', '403955002', '403961000000102', '403977003', '403978008', '403979000', '403986008', '403996004', '404008001', '404009009', '404010004', '404014008', '404015009', '404016005', '404017001', '404031000000109', '404037002', '404038007', '404039004', '404040002', '404041003', '404045007', '404046008', '404047004', '404051002', '404052009', '404053004', '404054005', '404055006', '404056007', '404067008', '404068003', '404069006', '404070007', '404071006', '404072004', '404073009', '404077005', '404079008', '404080006', '404081005', '404087009', '404088004', '404089007', '404090003', '404091004', '404092006', '404093001', '404094007', '404103007', '404104001', '404105000', '404106004', '404107008', '404108003', '404109006', '40411000', '404110001', '404111002', '404112009', '404113004', '404114005', '404115006', '404116007', '404117003', '404118008', '404119000', '404120006', '404121005', '404122003', '404123008', '404124002', '404125001', '404126000', '404127009', '404128004', '404129007', '404130002', '404131003', '404132005', '404133000', '404134006', '404135007', '404136008', '404137004', '404138009', '404139001', '404140004', '404141000', '404142007', '404143002', '404144008', '404145009', '404146005', '404147001', '404148006', '404149003', '404150003', '404151004', '404152006', '404153001', '404154007', '404155008', '404156009', '404157000', '404169008', '404172001', '404664002', '405546008', '405822008', '405843009', '405943005', '405945003', '4079000', '408281000000107', '408321000000104', '408331000000102', '408351000000109', '408381000000103', '408401000000103', '408491000000105', '408591000000106', '408621000000109', '408642003', '408643008', '408644002', '408645001', '408646000', '408647009', '408648004', '408649007', '408661000000101', '408771000000104', '408791000000100', '408801000000101', '408811000000104', '408831000000107', '408851000000100', '408861000000102', '408871000000109', '408881000000106', '408911000000106', '409151000000100', '409171000000109', '409231000000104', '409471000000101', '409561000000107', '409571000000100', '409651000000108', '409691000000100', '409701000000100', '409831000000100', '409841000000109', '410571000000108', '410671000000109', '411551000000106', '411561000000109', '411991000000107', '412761000000106', '412771000000104', '413389003', '413440007', '413441006', '413442004', '413443009', '413445002', '413446001', '4135001', '413526008', '413527004', '413537009', '413587002', '413616009', '413656006', '413737006', '413742003', '413743008', '413836008', '413840004', '413841000', '413842007', '413843002', '413847001', '413961000000100', '413990004', '414141000000102', '414166008', '414451000000109', '414553000', '414628006', '414654003', '414655002', '414676007', '414711000000102', '414780005', '414785000', '414791003', '415109007', '415110002', '415111003', '415112005', '415113000', '415156005', '415177008', '415283002', '415284008', '415285009', '415286005', '415287001', '41529000', '415461000000108', '415701004', '41607009', '416274001', '416501000000103', '416669000', '416712009', '416769008', '416842003', '416901000000108', '416901002', '417181009', '417417007', '417554000', '417570003', '417681000000107', '418152006', '418265009', '418372008', '418529003', '418628003', '418789003', '418829009', '419018000', '419035001', '419052002', '419094004', '41919003', '419240004', '419283005', '419386004', '419392005', '419427007', '419471000000104', '419563005', '419586003', '419591000000104', '419641000000108', '419662008', '419770008', '419842002', '419879004', '420028002', '420063007', '420301000', '420302007', '420441000000109', '420519005', '420524008', '420631000000103', '420788006', '420890002', '421246008', '421249001', '421283008', '421414006', '421630008', '421658000', '421686008', '421835000', '421918000', '42194009', '421980000', '422052002', '42215000', '422238009', '422282000', '422350000', '422378004', '422541001', '422572002', '422599000', '422676009', '422691006', '422736007', '422758009', '422782004', '422833009', '422853008', '422886007', '422968005', '4230004', '423005002', '423032007', '423038006', '423050000', '423106003', '423121009', '423158009', '423189008', '423195009', '423280002', '423284006', '423294001', '423295000', '423318000', '423325007', '423348002', '423349005', '423384009', '423424005', '423425006', '423447006', '423463003', '423464009', '423468007', '423494003', '423506005', '423535002', '423595004', '423600008', '423607006', '423610004', '423615009', '423627007', '423673009', '423691004', '423699002', '423700001', '423708008', '423746001', '423793008', '423807009', '423812005', '423829008', '423896007', '42392001', '423973006', '423987006', '424052001', '424132000', '424151006', '424190005', '424260006', '424276002', '424302003', '424334007', '424413001', '424440001', '424487008', '424549003', '424621000000108', '424631000000105', '424666005', '424671000000107', '424721000000102', '424771000000103', '424779008', '424849005', '424887002', '424938000', '424952003', '424954002', '424970000', '425037004', '425048006', '425066001', '425121000000100', '425127006', '425131000000103', '425148008', '425161000000108', '425178004', '425191000000102', '425211000000103', '425221000000109', '425225007', '425230006', '425231005', '425251000000104', '425261000000101', '425303004', '425376008', '425511000000100', '425521000000106', '425531000000108', '425535000', '425541000000104', '425657001', '425688002', '425749006', '425869007', '425941000000105', '425941003', '425951000000108', '426031000000107', '426071002', '426124006', '426191000000107', '426217000', '426248008', '426336007', '426370008', '426642002', '426711000000102', '426721000000108', '426741000000101', '426751000000103', '426885008', '426955004', '426964009', '427038005', '427051000000102', '427056005', '427141000000104', '427141003', '427151000000101', '427492003', '427642009', '427658007', '427685000', '428031000000108', '428061005', '428100006', '428281000', '428301000000104', '428322007', '428411000000106', '428905002', '428971000000106', '429033009', '429114002', '429221000000106', '429391000000101', '430338009', '4305004', '430556008', '430621000', '430841000000108', '430991000000109', '431396003', '43149009', '431561000000103', '431831000000105', '432082001', '432271000000102', '432328008', '43296007', '433067002', '433141000000108', '433531000000109', '437201000000108', '437231000000102', '437751000000102', '437761000000104', '437771000000106', '437781000000108', '437801000000109', '437811000000106', '437821000000100', '437871000000101', '438191000000105', '438231000000101', '438251000000108', '438271000000104', '438291000000100', '438321000000105', '438331000000107', '438351000000100', '438401000000106', '438671000000101', '438681000000104', '438691000000102', '438701000000102', '438946002', '439141000000106', '439151000000109', '439161000000107', '439171000000100', '439271000000100', '439291000000106', '439478008', '439740005', '43985008', '440173001', '440397000', '440422002', '440501006', '440525003', '440527006', '440781000000105', '440791000000107', '441161000000109', '441313008', '441535001', '441559006', '441962003', '442537007', '44255004', '442591000000102', '442771000000109', '442921000000108', '443136000', '443144000', '443250000', '443261008', '443333004', '443439001', '443487006', '443488001', '443489009', '443493003', '443501000000104', '443520009', '443565000', '443621000000109', '443641000000102', '443648003', '443675005', '443677002', '443679004', '443719001', '443757001', '443936004', '443937008', '443951000000105', '443961001', '444134008', '444596001', '444597005', '444604002', '444712000', '44474009', '444910004', '444911000', '445105005', '445227008', '445238008', '445269007', '4529004', '445406001', '445448008', '445681000000106', '445691000000108', '445925008', '446022000', '446189008', '446578007', '446643000', '446688004', '446807009', '446939001', '447011009', '447100004', '447109003', '447266004', '447389009', '447390000', '447416008', '447656001', '447658000', '44769000', '447705002', '447706001', '447707005', '447708000', '447710003', '447712006', '447730004', '447738006', '447757002', '447766003', '447781009', '447782002', '447783007', '447784001', '447785000', '447792005', '447800002', '447804006', '447805007', '447806008', '447882007', '447883002', '447885009', '447886005', '447949005', '447951009', '447989004', '448139005', '448165009', '448212009', '448213004', '448214005', '448215006', '448216007', '448217003', '448218008', '448220006', '448221005', '448229007', '448231003', '448233000', '448248006', '448250003', '448254007', '448257000', '448258005', '448259002', '448269008', '448273006', '448274000', '448296006', '448298007', '448299004', '448300007', '448314007', '448315008', '448317000', '448319002', '448354009', '448371005', '448372003', '448376000', '448377009', '448378004', '448384001', '448386004', '448387008', '448388003', '448401007', '448408001', '448435005', '448436006', '448447004', '448449001', '448450001', '448451002', '448465000', '448468003', '448509007', '448553002', '448554008', '448555009', '448558006', '448559003', '448560008', '448561007', '448607004', '448609001', '448663003', '448664009', '448665005', '448666006', '448668007', '448669004', '448670003', '448672006', '448673001', '448674007', '448675008', '448709005', '448710000', '448711001', '448712008', '448738008', '448774004', '448775003', '448776002', '448863000', '448864006', '448865007', '448867004', '448868009', '448882009', '448922007', '448930008', '448931007', '448952004', '448954003', '448988009', '448989001', '448990005', '448992002', '448993007', '448994001', '448995000', '449034009', '449053004', '449054005', '449055006', '449058008', '449059000', '449063007', '449065000', '449066004', '449067008', '449072004', '449073009', '449074003', '449075002', '449077005', '449096009', '449100005', '449101009', '449108003', '449153001', '449156009', '449173006', '449176003', '449177007', '449206007', '449207003', '449208008', '449211009', '449216004', '449217008', '449218003', '449219006', '449220000', '449221001', '449222008', '449223003', '449224009', '449248000', '449253005', '449254004', '449259009', '449260004', '449267001', '449268006', '449269003', '449292003', '449293008', '449294002', '449295001', '449307001', '449308006', '449309003', '449318001', '449377002', '449386007', '449416001', '449417005', '449418000', '449419008', '449420002', '449421003', '449472007', '449487002', '449497006', '449578008', '449627008', '449628003', '449630001', '449631002', '449632009', '449633004', '449634005', '449635006', '449636007', '449637003', '449731000000104', '449741000000108', '449751000000106', '449791000000103', '449803009', '449881000000109', '449891000000106', '450041000000102', '450051000000104', '450141000000101', '450171000000107', '450181000000109', '450191000000106', '450201000000108', '45024009', '450611000000107', '450851000000105', '450861000000108', '450902001', '450907007', '450908002', '450909005', '450910000', '450911001', '450913003', '450914009', '450915005', '450916006', '450917002', '450920005', '450928003', '450929006', '450935006', '450937003', '450943001', '450944007', '450949002', '450950002', '450951003', '450952005', '450953000', '450954006', '450955007', '450956008', '450958009', '450959001', '450971000000109', '450981000000106', '450991000000108', '451111000000105', '451121000000104', '451131000000102', '451141000000106', '45221000119105', '453011000000105', '45410002', '454411000000101', '454781000000102', '455391000000100', '455551000000102', '45572000', '455771000000103', '455781000000101', '456171000000109', '456651000000108', '457051000000103', '457421000000102', '457571000000102', '45881000', '4590003', '461431000000101', '461841000000107', '461871000000101', '461881000000104', '461891000000102', '461901000000101', '461931000000107', '462251000000102', '462341000000107', '462371000000101', '462381000000104', '462391000000102', '462401000000104', '462451000000103', '462461000000100', '462491000000106', '462711000000105', '462951000000106', '462961000000109', '462971000000102', '463091000000106', '463221000000103', '463341000000103', '463371000000109', '463381000000106', '463391000000108', '46347006', '464031000000108', '464261000000109', '464771000000108', '46585005', '466481000000108', '466491000000105', '466521000000108', '467071000000100', '46710009', '467201000000100', '46732000', '46744002', '467881000000109', '467891000000106', '468291000000108', '468681000000109', '468691000000106', '46923007', '469881000000100', '470351000000104', '47107000', '471231000000108', '471831000000107', '471861000000102', '472871000000100', '472951000000104', '472961000000101', '473031000000100', '473041000000109', '473419009', '477471000000101', '477491000000102', '477501000000108', '477531000000102', '477541000000106', '477571000000100', '477841000000109', '477851000000107', '477861000000105', '477871000000103', '478021000000105', '478041000000103', '478081000000106', '478091000000108', '478111000000103', '478121000000109', '478131000000106', '478151000000104', '478171000000108', '478201000000109', '478211000000106', '478231000000103', '478611000000108', '478871000000102', '478881000000100', '478981000000107', '479011000000106', '479021000000100', '479031000000103', '4797003', '480061000000108', '480071000000101', '48460009', '48554007', '48952003', '49430005', '4950009', '49937004', '50007008', '50102004', '50150000', '50422007', '5052009', '50542000', '50813003', '51092000', '51217003', '51465000', '51549004', '51757004', '52040006', '52178006', '52220008', '52248008', '5257006', '531581000000101', '532061000000105', '53237008', '533371000000109', '533381000000106', '533391000000108', '533411000000108', '533431000000100', '533471000000103', '533491000000104', '533951000000104', '534631000000101', '534641000000105', '534651000000108', '534661000000106', '534671000000104', '534681000000102', '534691000000100', '534781000000105', '534791000000107', '534801000000106', '534811000000108', '534821000000102', '534831000000100', '534841000000109', '534851000000107', '535821000000103', '536091000000103', '53618008', '536201000000102', '53654007', '536941000000107', '537911000000109', '537921000000103', '537931000000101', '537941000000105', '537951000000108', '537961000000106', '537981000000102', '537991000000100', '538001000000109', '53801007', '538011000000106', '538021000000100', '538031000000103', '538041000000107', '538081000000104', '538091000000102', '538111000000107', '539391000000105', '539511000000109', '539521000000103', '539531000000101', '539541000000105', '539571000000104', '539581000000102', '53968002', '539701000000104', '539711000000102', '539731000000105', '539741000000101', '540241000000100', '540251000000102', '540261000000104', '540281000000108', '540291000000105', '540301000000109', '540311000000106', '540321000000100', '54087003', '541001000000104', '541011000000102', '541021000000108', '541031000000105', '541111000000101', '541121000000107', '541131000000109', '54124005', '541651000000105', '541661000000108', '541671000000101', '541691000000102', '541701000000102', '541711000000100', '541731000000108', '541741000000104', '541881000000103', '541891000000101', '542031000000104', '542881000000106', '543361000000101', '543371000000108', '543381000000105', '543391000000107', '543401000000105', '543411000000107', '543421000000101', '543431000000104', '54443001', '545241000000108', '546321000000103', '546351000000108', '54666007', '546691000000106', '546701000000106', '546821000000109', '546831000000106', '546841000000102', '546851000000104', '546861000000101', '546871000000108', '546881000000105', '547121000000104', '547131000000102', '547141000000106', '547151000000109', '547161000000107', '54734006', '548991000000100', '549521000000102', '549531000000100', '549541000000109', '549551000000107', '549561000000105', '549571000000103', '549581000000101', '549601000000105', '549611000000107', '549621000000101', '550081000000103', '550091000000101', '550101000000109', '550111000000106', '55020008', '55045006', '55150002', '552141000000106', '552161000000107', '55320002', '553641000000108', '555731000000108', '555741000000104', '555761000000103', '556021000000102', '556031000000100', '556041000000109', '556051000000107', '556061000000105', '556071000000103', '556141000000105', '556151000000108', '556161000000106', '556171000000104', '556221000000107', '557081000000104', '557091000000102', '557101000000105', '557121000000101', '557131000000104', '557141000000108', '557151000000106', '557161000000109', '557171000000102', '557491000000109', '557501000000103', '557641000000101', '557651000000103', '557661000000100', '557671000000107', '557691000000106', '558011000000107', '558021000000101', '559181000000106', '55921005', '55937004', '559871000000103', '559881000000101', '561431000000107', '561451000000100', '561461000000102', '561471000000109', '561481000000106', '561581000000107', '561591000000109', '56346006', '56422000', '56484001', '564881000000107', '564891000000109', '564901000000105', '564911000000107', '564921000000101', '564931000000104', '565581000000109', '565591000000106', '565601000000100', '56565002', '565911000000106', '565961000000108', '565971000000101', '566041000000100', '566521000000100', '566531000000103', '566541000000107', '566551000000105', '567711000000101', '569271000000107', '569281000000109', '569291000000106', '569301000000105', '569311000000107', '569321000000101', '569331000000104', '569351000000106', '56944001', '5701003', '570611000000103', '570621000000109', '570661000000101', '571181000000107', '571191000000109', '57141000', '572191000000103', '572201000000101', '572591000000105', '572601000000104', '573341000000101', '573361000000100', '573381000000109', '573431000000103', '573441000000107', '573471000000101', '573481000000104', '573521000000104', '573621000000105', '573631000000101', '573641000000105', '573651000000108', '573661000000106', '573671000000104', '574411000000109', '574421000000103', '575491000000101', '575501000000107', '575511000000109', '575521000000103', '57596004', '57622002', '578161000000107', '578171000000100', '578251000000103', '578281000000109', '578331000000104', '578391000000103', '578441000000102', '578451000000104', '578481000000105', '578501000000101', '579201000000107', '579211000000109', '579221000000103', '579231000000101', '579241000000105', '579251000000108', '579261000000106', '579281000000102', '579291000000100', '579301000000101', '579311000000104', '579321000000103', '58069009', '58248003', '58477004', '58961005', '59238007', '59367005', '5958006', '59583009', '60346004', '60620005', '60718004', '609515005', '61217001', '61291000119103', '61301000119102', '61471000119100', '61493004', '615431000000101', '615441000000105', '620431000000103', '620441000000107', '62064005', '6219000', '62383007', '62497000', '6250003', '62621002', '62681000', '63211008', '63264007', '633321000000101', '633331000000104', '63364005', '63373002', '63449009', '634561000000104', '634581000000108', '634591000000105', '634601000000104', '634611000000102', '634621000000108', '634631000000103', '634641000000101', '634911000000105', '634921000000104', '634931000000102', '634951000000109', '634961000000107', '634971000000100', '634981000000103', '634991000000101', '635001000000108', '635011000000105', '635021000000104', '635031000000102', '6357002', '63634009', '636471000000100', '636481000000103', '6381009', '638271000000102', '64000002', '640511000000108', '64059005', '6426003', '645061000000108', '64524002', '64575004', '648451000000106', '6492006', '65140001', '65278006', '65399007', '65646006', '657351000000103', '657371000000107', '657401000000109', '659491000000109', '662421000000106', '662431000000108', '662441000000104', '662451000000101', '663631000000101', '663641000000105', '663651000000108', '663661000000106', '66445009', '66515009', '666471000000109', '666481000000106', '666491000000108', '666501000000102', '666511000000100', '666541000000104', '668601000000102', '669301000000101', '670121000000109', '67280001', '673771000000103', '675601000000100', '67811000119102', '67821000119109', '67830002', '67831000119107', '67841000119103', '681601000119101', '681621000119105', '681631000119108', '681721000119103', '68358000', '683991000119103', '684911000119105', '68614005', '687911000000109', '687921000000103', '687931000000101', '688151000000106', '68827007', '68880006', '689131000000106', '68979007', '69028005', '690661000000104', '69077002', '691151000000104', '691161000000101', '69317001', '69408002', '69515008', '695601000000109', '697551000000102', '697993003', '698011002', '698040004', '698041000', '698042007', '698043002', '698044008', '698045009', '698048006', '698151000000107', '698161000000105', '698181000000101', '698200003', '698255000', '698285005', '698286006', '698287002', '698288007', '698646006', '699028006', '699229009', '699317002', '699318007', '699354006', '699355007', '699356008', '699357004', '699358009', '699657009', '699658004', '699659007', '699704002', '699818003', '700064004', '700423003', '700488005', '7010000', '70179006', '702346005', '702368000', '702369008', '702391001', '702392008', '702405001', '702446006', '702467006', '702476004', '702741000000102', '702785000', '702786004', '702977001', '703228009', '703230006', '703387000', '703429003', '703541000000100', '703545003', '703549009', '703577000', '703595002', '703596001', '703609007', '703615007', '703617004', '703618009', '703625002', '703626001', '703650004', '703655009', '703692009', '703693004', '703694005', '703699000', '703700004', '703816006', '703818007', '703819004', '703820005', '703821009', '703822002', '704152002', '704441000000101', '704551000000104', '704561000000101', '705061009', '705141000000109', '705145009', '705176003', '70555003', '70600005', '706970001', '707146006', '707337006', '707339009', '707342003', '707343008', '707344002', '707345001', '707346000', '707347009', '707348004', '707349007', '707350007', '707353009', '707354003', '707355002', '707356001', '707357005', '707358000', '707359008', '707360003', '707361004', '707362006', '707377003', '707378008', '707379000', '707380002', '707383000', '707384006', '707385007', '707386008', '707388009', '707389001', '707390005', '707391009', '707392002', '707393007', '707394001', '707395000', '707396004', '707397008', '707398003', '707399006', '707400004', '707401000', '707402007', '707403002', '707404008', '707405009', '707406005', '707407001', '707408006', '707409003', '707410008', '707411007', '707421004', '707422006', '707423001', '707424007', '707425008', '707426009', '707427000', '707429002', '707430007', '707451005', '707452003', '707453008', '707454002', '707455001', '707456000', '707457009', '707458004', '707460002', '707464006', '707465007', '707466008', '707467004', '707468009', '707469001', |

| | |
|---|---|
| | '707470000', '707471001', '707472008', '707473003', '707475005', '707479004', '707481002', '707482009', '707483004', '707484005', '707485006', '707486007', '707487003', '707489000', '707490009', '707491008', '707492001', '707493006', '707494000', '707495004', '707528007', '707529004', '707531008', '707532001', '707535004', '707537007', '707538002', '707539005', '707575007', '707576008', '707579001', '707580003', '707581004', '707582006', '707583001', '707584007', '707585008', '707586009', '707587000', '707588005', '707589002', '707590006', '707591005', '707592003', '707593008', '707595001', '707596000', '707627009', '707628004', '707660007', '707662004', '707664003', '707670009', '707671008', '707672001', '707673006', '707674000', '707686002', '707697002', '707703001', '707704007', '707705008', '707761000000104', '708054009', '708504008', '708921005', '708937004', '708971008', '708979005', '709031009', '70910003', '709190005', '709191009', '709285002', '709471005', '709517003', '709830006', '710195004', '710196003', '71109004', '71111000119109', '711414003', '711701000000107', '712525007', '712750007', '712837004', '712849003', '713038003', '713189001', '713290004', '713293002', '713325002', '713327005', '713483007', '713516007', '713572001', '713573006', '713574000', '713609000', '713646001', '713718006', '713897006', '71390001', '714251006', '714463003', '71447003', '715215007', '715412008', '715414009', '715664005', '715901002', '715903004', '715904005', '715950008', '716179006', '716274007', '716318002', '716586009', '716588005', '716593008', '716647001', '716648006', '716649003', '716651004', '716652006', '716653001', '716654007', '716657000', '716659002', '716788007', '716789004', '716855006', '716857003', '716859000', '717731002', '717734005', '717735006', '717736007', '717916003', '717921000', '717922007', '717968005', '718195003', '718200007', '718220008', '718604008', '718721006', '719052006', '71985006', '719952009', '720006006', '720346009', '720587009', '721302006', '721303001', '721304007', '721305008', '721306009', '721308005', '721310007', '721311006', '721313009', '721314003', '721540005', '721541009', '721546004', '721547008', '721548003', '721549006', '721555001', '721556000', '721557009', '721558004', '721559007', '721560002', '721561003', '721562005', '721563000', '721567004', '721568009', '721569001', '721574009', '721575005', '721576006', '721577002', '721578007', '721579004', '721580001', '721602000', '721603005', '721604004', '721605003', '721606002', '721607006', '721617001', '721618006', '721619003', '721620009', '721621008', '721622001', '721623006', '721624000', '721625004', '721626003', '721627007', '721628002', '721629005', '721630000', '721632008', '721633003', '721634009', '721635005', '721636006', '721637002', '721638007', '721639004', '721640002', '721641003', '721642005', '721643000', '721644006', '721645007', '721669006', '721670007', '721671006', '721672004', '721673009', '721695008', '721696009', '721697000', '721698005', '721699002', '721700001', '721701002', '721709000', '721716004', '721718003', '721725005', '72174007', '721762007', '722103009', '722112006', '722229001', '722425009', '722444002', '722445001', '722509001', '722510006', '722511005', '722512003', '722513008', '722514002', '722515001', '722516000', '722517009', '722518004', '722519007', '722524005', '722527003', '722528008', '722529000', '722530005', '722533007', '722542000', '722543005', '722665003', '722666002', '722670005', '722671009', '722672002', '722673007', '722674001', '722676004', '722677008', '722678003', '722679006', '722680009', '722681008', '722682001', '722683006', '722684000', '722685004', '722686003', '722687007', '722688002', '722707001', '722712000', '722714004', '722718001', '722795004', '722827008', '722828003', '722832009', '722953004', '722954005', '723076008', '723077004', '723173003', '723182009', '723265000', '723281005', '723301009', '723843005', '723844004', '723845003', '723846002', '723847006', '723848001', '723849009', '723850009', '723851008', '723852001', '723853006', '723854000', '723855004', '723856003', '723879002', '723887001', '723889003', '724056005', '724058006', '724059003', '724060008', '724467001', '724468006', '724536000', '724537009', '724552002', '724553007', '724554001', '724644005', '724645006', '724647003', '724648008', '724649000', '724650000', '724805000', '724806004', '724866001', '72495009', '725390002', '725391003', '725437002', '726019003', '726652005', '726653000', '726654006', '726721002', '72891000119103', '72907003', '732201008', '732968009', '733064004', '733081008', '733123006', '733134009', '733135005', '733136006', '733144006', '733162002', '733163007', '733343005', '733344004', '733345003', '733346002', '733347006', '733348001', '733349009', '733350009', '733351008', '733352001', '733353006', '733354000', '733355004', '733356003', '733357007', '733358002', '733359005', '733360000', '733361001', '733362008', '733470002', '733471003', '733348003', '733598001', '733602004', '733603009', '733607005', '733608000', '733627006', '733834006', '733843002', '733844008', '733848006', '733860007', '733881007', '733895005', '733906003', '733912008', '733913003', '733917002', '733926004', '734014001', '734015000', '734033006', '734035004', '734038002', '734044003', '734066005', '734067001', '734068006', '734069003', '734072005', '734076008', '734086009', '734087000', '734093008', '734095001', '734099007', '734134003', '734141009', '734142002', '734522002', '734524001', '734933008', '73506006', '735332000', '735385007', '735450006', '735678002', '735679005', '735680008', '735735001', '735757008', '735916009', '735917000', '735918005', '735919002', '735920008', '735921007', '736322001', '73676002', '737058005', '737308008', '737309000', '737310005', '737311009', '737312002', '737313007', '738527001', '738770003', '7391000119103', '739301006', '74042001', '74053007', '74153005', '74189002', '74279005', '74409009', '74532006', '74654000', '74739000', '75339006', '754351000000100', '754361000000102', '754371000000109', '75622000', '756621000000106', '757251000000101', '757351000000109', '757711000000109', '757831000000107', '757841000000103', '75931002', '760591000000101', '76060004', '760641000000105', '760951000000102', '760981000000108', '761958009', '762315004', '762457009', '762458004', '762665002', '762690000', '762691001', '763063001', '763064007', '76312009', '763131005', '763309005', '763408003', '763409006', '763477007', '763479005', '763666008', '763719001', '763771009', '763796007', '763884007', '76422004', '764694005', '764735002', '764737005', '764845008', '764846009', '764847000', '764855007', '764856008', '764938007', '764940002', '764951002', '764952009', '764961009', '764990003', '764991004', '765094003', '765095002', '765136002', '765143008', '765328000', '76564002', '765740002', '765741003', '76594008', '766045006', '766046007', '766048008', '766247009', '766248004', '766757006', '766758001', '766759009', '766930002', '766935007', '766978002', '766979005', '766980008', '766981007', '767047005', '767444009', '767448007', '767511000000109', '76817009', '76909002', '770531000000106', '77381001', '77430005', '77455004', '77659000', '778041000000106', '778061000000107', '778071000000100', '778081000000103', '778091000000101', '778101000000109', '77865003', '77870005', '78010001', '78197004', '78303004', '78411000119107', '78453009', '78838008', '79143006', '79281000119106', '80061003', '80091008', '804391000000100', '80440003', '80570006', '80727009', '80914001', '812871000000103', '813671000000107', '814201000000103', '8145008', '814851000000101', '814981000000100', '815011000000107', '815031000000104', '815091000000103', '815101000000106', '815121000000102', '815131000000100', '815141000000109', '815161000000105', '815191000000104', '815271000000105', '815311000000105', '815361000000107', '815451000000102', '815671000000108', '815681000000105', '81622000', '81920005', '820601000000103', '821881000000100', '821891000000103', '82267002', '82501000119102', '82546001', '82591005', '82711006', '83118000', '83217000', '832521000000108', '832531000000105', '832561000000100', '832581000000109', '832591000000106', '832601000000100', '832611000000103', '832621000000109', '832631000000106', '832641000000102', '832651000000104', '832661000000101', '832671000000108', '832681000000105', '832691000000107', '832811000000104', '832821000000105', '832911000000106', '83292005', '832921000000100', '833001000000103', '833021000000107', '833031000000109', '833061000000104', '833081000000108', '833091000000105', '833101000000102', '833111000000100', '833121000000106', '833141000000104', '833171000000105', '833331000000106', '833341000000102', '833351000000104', '833361000000101', '833371000000108', '833381000000105', '833391000000107', '833401000000105', '833411000000107', '833431000000104', '833441000000108', '833451000000106', '833461000000109', '833471000000102', '833481000000100', '833491000000103', '833501000000109', '833511000000106', '833521000000100', '833531000000103', '833541000000107', '833551000000105', '833571000000101', '833581000000104', '833591000000102', '833601000000108', '833611000000105', '833621000000104', '833631000000102', '833641000000106', '833691000000101', '834551000000108', '834561000000106', '834571000000104', '835009', '83950009', '84290008', '84350008', '84427001', '84631004', '84633001', '84664004', '847481000000109', '847631000000107', '847651000000100', '847691000000108', '847701000000108', '847741000000106', '849141000000100', '855061000000104', '855201000000106', '85530002', '85654004', '857651000000102', '86049000', '86293007', '863131000000103', '863141000000107', '863741000000108', '863751000000106', '863761000000109', '863771000000102', '863781000000100', '863791000000103', '86562005', '867931000000106', '867941000000102', '87091000119101', '87111000119109', '87121000119102', '87151000119105', '87163000', '8734000', '87364003', '875231000000102', '875381000000107', '875391000000109', '87737001', '87992000', '88195001', '88252006', '88253001', '882791000000109', '88334008', '88400008', '884601000000103', '884611000000101', '88591002', '887521000000104', '887531000000102', '88897007', '8900005', '89277004', '89487002', '89623007', '89706008', '89740008', '89880005', '900006', '90120004', '90282004', '90725004', '91031000119108', '91041000119104', '91061000119100', '91081000119109', '91113000', '91131000119109', '91151000119103', '91161000119101', '91171000119107', '91281000119103', '915007', '916281000000108', '918261000000103', '91854005', '91855006', '91856007', '91857003', '91858008', '91859000', '91860005', '91861009', '92508006', '92509003', '92510008', '92511007', '92512000', '92513005', '92514004', '92515003', '92516002', '92526009', '92527000', '92528005', '92573003', '92581002', '92604004', '92661009', '92675002', '92683008', '92811003', '92812005', '92813000', '92814006', '92817004', '92818009', '9294008', '93133006', '93134000', '93135004', '93136003', '93137007', '93138002', '93139005', '93140007', '93141006', '93142004', '93143009', '93144003', '93145002', '93146001', '93147005', '93148000', '93149008', '93150008', '93151007', '93152000', '93169003', '93182006', '93183001', '93184007', '93185008', '93186009', '93187000', '93188005', '93189002', '93190006', '93191005', '93192003', '93193008', '93194002', '93195001', '93196000', '93197009', '93198004', '93199007', '93200005', '93201009', '93202002', '93203007', '93204001', '93205000', '93206004', '93207008', '93208003', '93209006', '93210001', '93211002', '93212009', '93213004', '93214005', '93215006', '93216007', '93217003', '93218008', '93219000', '93220006', '93221005', '93222003', '93223008', '93224002', '93225001', '93226000', '93227009', '93228004', '93229007', '93230002', '93450001', '93451002', '93487009', '93488004', '93489007', '93492006', '93493001', '93494007', '93495008', '93496009', '93497000', '93498005', '93500006', '93501005', '93505001', '93506000', '93507009', '93509007', '93510002', '93514006', '93515007', '93516008', '93518009', '93519001', '93520007', '93521006', '93522004', '93523009', '93524003', '93525002', '93526001', '93527005', '93528000', '93530003', '93531004', '93532006', '93533001', '93534007', '93536009', '93537000', '93541001', '93542008', '93543003', '93545005', '93546006', '93547002', '93548007', '93549004', '93550004', '93551000', '93552007', '93554008', '93555009', '93636004', '93637008', '93638003', '93639006', '93640008', '93641007', '93642000', '93643005', '93644004', '93645003', '93646002', '93647006', '93648001', '93649009', '93650009', '93651008', '93652001', '93653006', '93654000', '93655004', '93657007', '93658002', '93659005', '93660000', '93661001', '93662008', '93663003', '93665005', '93667002', '93669004', '93670003', '93671004', '93672006', '93674007', '93676009', '93677000', '93679002', '93680004', '93681000', '93682007', '93683002', '93684008', '93686005', '93687001', '93689003', '93690007', '93692004', '93693009', '93694003', '93695002', '93696001', '93697005', '93698000', '93699008', '93700009', '93701008', '93703006', '93704000', '93705004', '93706003', '93707007', '93708002', '93709005', '93710000', '93711001', '93712008', '93713003', '93714009', '93715005', '93716006', '93717002', '93720005', '93721009', '93722002', '93723007', '93724001', '93725000', '93726004', '93727008', '93728003', '93729006', '93730001', '93731002', '93732009', '93733004', '93734005', '93737003', '93738008', '93739000', '93740003', '93741004', '93742006', '93743001', '93744007', '93745008', '93746009', '93747000', '93748005', '93749002', '93751003', '93753000', '93754006', '93755007', '93756008', '93757004', '93759001', '93760006', '93761005', '93763008', '93764002', '93766000', '93767009', '93768004', '93769007', '93770008', '93771007', '93772000', '93773005', '93775003', '93776002', '93777006', '93778001', '93779009', '93781006', '93782004', '93783009', '93784003', '93786001', '93787005', '93788000', '93789008', '93792007', '93793002', '93796005', '93797001', '93798006', '93799003', '93800004', '93801000', '93802007', '93803002', '93804008', '93806005', '93807001', '93808006', '93809003', '93812000', '93814004', '93815003', '93816002', '93817006', '93818001', '93820003', '93821004', '93824007', '93825008', '93826009', '93827000', '93828005', '93829002', '93830007', '93831006', '93832004', '93833009', '93834003', '93835002', '93836001', '93837005', '93839008', '93840005', '93841009', '93842002', '93843007', '93844001', '93845000', '93846004', '93848003', '93849006', '93850006', '93851005', '93852003', '93854002', '93857009', '93858004', '93860002', '93861003', '93862005', '93864006', '93865007', '93866008', '93867004', '93868009', '93870000', '93871001', '93872008', '93874009', '93875005', '93876006', '93878007', '93879004', '93880001', '93882009', '93883004', '93884005', '93885006', '93886007', '93887003', '93888008', '93889000', '93890009', '93891008', '93892001', '93893006', '93894000', '93898002', '93899005', '93900000', '93901001', '93902008', '93903003', '93904009', '93905005', '93906006', '93907002', '93908007', '93909004', '93910009', '93911008', '93912001', '93913006', '93914000', '93915004', '93916003', '93917007', '93918002', '93920004', '93922007', '93923002', '93924008', '93925009', '93926005', '93927001', '93928006', '93929003', '93930008', '93931007', '93932000', '93933005', '93934004', '93936002', '93939009', '93940006', '93941005', '93942003', '93943008', '93944002', '93945001', '93946000', '93947009', '93948004', '93950007', '9395006', '93951006', '93952004', '93953009', '93955002', '93956001', '93957005', '93958000', '93959008', '93960003', '93961004', '93962006', '93963001', '93964007', '93965008', '93966009', '93967000', '93968005', '93969002', '93970001', '93971002', '93973004', '93974005', '93975006', '93976007', '93977003', '93978008', '93979000', '93980002', '93981003', '93982005', '93983000', '93984006', '93985007', '93986008', '93987004', '93989001', '93990005', '93991009', '93992002', '93993007', '93994001', '93995000', '93996004', '93997008', '93998003', '94000008', '94001007', '94002000', '94003005', '94004004', '94005003', '94006002', '94007006', '94008001', '94010004', '94011000', '94012007', '94013002', '94014008', '94015009', '94016005', '94017001', '94018006', '94019003', '94020009', '94021008', '94022001', '94023006', '94024000', '94025004', '94026003', '94027007', '94028002', '94029005', '94030000', '94031001', '94032008', '94033003', '94034009', '94035005', '94036006', '94037002', '94038007', '94039004', '94040002', '94041003', '94042005', '94043000', '94044006', '94045007', '94046008', '94047004', '94048009', '94049001', '94050001', '94051002', '94052009', '94053004', '94054005', '94055006', '94056007', '94057003', '94058008', '94059000', '94060005', '94061009', '94062002', '94063007', '94066004', '94067008', '94068003', '94069006', '94071006', '94072004', '94073009', '94075002', '94076001', '94077005', '94078000', '94079008', '94080006', '94081005', '94082003', '94083008', '94084002', '94085001', '94086000', '94087009', '94089007', '94090003', '94092006', '94093001', '94094007', '94096009', '94097000', '94098005', '94099002', '94101009', '94102002', '94103007', '94104001', '94105000', '94107008', '94108003', '94109006', '94110001', '94111002', '94112009', '94113004', '94115006', '94116007', '94117003', '94118008', '94120006', '94121005', '94122003', '94123008', '94124002', '94125001', '94126000', '94128004', '94129007', '94131003', '94132005', '94133000', '94134006', '94135007', '94138009', '94139001', '94140004', '94142007', '94143002', '94144008', '94145009', '94148006', '94149003', '94152006', '94153001', '94154007', '94155008', '94156009', '94157000', '94158005', '94159002', '94160007', '94161006', '94162004', '94163009', '94164003', '94165002', '94166001', '94167005', '94168000', '94169008', '94170009', '94171008', '94172001', '94173006', '94175004', '94176003', '94177007', '94178002', '94179005', '94180008', '94181007', '94182000', '94183005', '94184004', '94185003', '94186002', '94187006', '94189009', '94190000', '94191001', '94192008', '94193003', '94194009', '94195005', '94196006', '94197002', '94198007', '94200001', '94201002', '94202009', '94203004', '94204005', '94205006', '94206007', '94207003', '94208008', '94209000', '94210005', '94211009', '94212002', '94213007', '94214001', '94215000', '94217008', '94218003', '94219006', '94220000', '94221001', '94222008', '94224009', '94225005', '94226006', '94227002', '94228007', '94229004', '94230009', '94231008', '94232001', '94233006', '94234000', '94235004', '94236003', '94237007', '94238002', '94239005', '94240007', '94241006', '94242004', '94243009', '94244003', '94245002', '94246001', '94247005', '94248000', '94249008', '94250008', '94252000', '94253005', '94254004', '94255003', '94256002', '94257006', '94258001', '94259009', '94260004', '94261000', '94262007', '94263002', '94264008', '94265009', '94266005', '94267001', '94268006', '94269003', '94270002', '94271003', '94272005', '94273000', '94274006', '94275007', '94276008', '94277004', '94278009', '94279001', '94280003', '94281004', '94282006', '94283001', '94284007', '94285008', '94286009', '94287000', '94288005', '94289002', '94290006', '94291000119103', '94291005', '94292003', '94293008', '94294002', '94295001', '94296000', '94297009', '94298004', '94299007', '94300004', '94301000', '94302007', '94303002', '94304008', '94305009', '94306005', '94307001', '94308006', '94309003', '94310008', '94311007', '94312000', '94313005', '94314004', '94315003', '94316002', '94317006', '94318001', '94319009', '94320003', '94321004', '94322006', '94323001', '94324007', '94325008', '94326009', '94327000', '94328005', '94329002', '94330007', '94331006', '94332004', '94333009', '94334003', '94335002', '94336001', '94337005', '94338000', '94339008', '94340005', '94341009', '94342002', '94343007', '94344001', '94345000', '94346004', '94347008', '94348003', '94349006', '94350006', '94351005', '94352003', '94353008', '94354002', '94355001', '94356000', '94357009', '94358004', '94359007', '94360002', '94361000000105', '94361003', '94362005', '94363000', '94364006', '94365007', '94366008', '94367004', '94368009', '94369001', '94370000', '94371001', '94372008', '94373003', '94374009', '94375005', '94376006', '94377002', '94378007', '94379004', '94380001', '94381002', '94382009', '94383004', '94384005', '94385006', '94386007', '94387003', '94388008', '94389000', '94390009', '94391008', '94392001', '94393006', '94394000', '94395004', '94396003', '94397007', '94398002', '94399005', '94400003', '94401004', '94402006', '94403001', '94404007', '94405008', '94406009', '94407000', '94408005', '94409002', '94410007', '94411006', '94412004', '94413009', '94414003', '94415002', '94416001', '94417005', '94418000', '94419008', '94420002', '94421003', '94422005', '94423000', '94424006', '94425007', '94426008', '94427004', '94428009', '94429001', '94430006', '94431005', '94432003', '94433008', '94434002', '94435001', '94436000', '94437009', '94439007', '94440009', '94441008', '94442001', '94443006', '94444000', '94445004', '94446003', '94447007', '94448002', '94449005', '94450005', '94451009', '94452002', '94453007', '94454001', '94455000', '94456004', '94457008', '94458003', '94459006', '94460001', '94461002', '94462009', '94463004', '94464005', '94465006', '94466007', '94467003', '94468008', '94469000', '94470004', '94471000', '94472007', '94473002', '94474008', '94475009', '94476005', '94477001', '94478006', '94479003', '94480000', '94481001', '94482008', '94483003', '94484009', '94485005', '94486006', '94487002', '94488007', '94489004', '94490008', '94491007', '94492000', '94493005', '94494004', '94495003', '94496002', '94497006', '94498001', '94499009', '94500000', '94501001', '94502008', '94503003', '94504009', '94505005', '94506006', '94507002', '94508007', '94509004', '94510009', '94511008', '94512001', '94513006', '94514000', '94515004', '94516003', '94517007', '94518002', '94519005', '94520004', '94521000', '94522007', '94523002', '94524008', '94525009', '94526005', '94527001', '94528006', '94529003', '94530008', '94531007', '94532000', '94533005', '94534004', '94535003', '94536002', '94537006', '94538001', '94539009', '94540006', '94541005', '94542003', '94543008', '94544002', '94545001', '94546000', '94547009', '94548004', '94549007', '94550007', '94551006', '94552004', '94553009', '94554003', '94555002', '94556001', '94557005', '94558000', '94559008', '94560003', '94561004', '94562006', '94563001', '94564007', '94565008', '94566009', '94567000', '94568005', '94569002', '94570001', '94571002', '94572009', '94573004', '94574005', '94575006', '94576007', '94577003', '94578008', '94579000', '94580002', '94581003', '94582005', '94583000', '94584006', '94585007', '94586008', '94587004', '94588009', '94589001', '94590005', '94591009', '94592002', '94593007', '94594001', '94595000', '94596004', '94597008', '94598003', '94599006', '94600009', '94601008', '94602001', '94603006', '94604000', '94605004', '94606003', '94607007', '94608002', '94609005', '94610000', '94611001', '94612008', '94613003', '94614009', '94615005', '94616006', '94617002', '94618007', '94619004', '94620005', '94621009', '94622002', '94623007', '94624001', '94625000', '94626004', '94627008', '94628003', '94629006', '94630001', '94631002', '94632009', '94633004', '94634005', '94635006', '94636007', '94637003', '94638008', '94639000', '94640003', '94641004', '94642006', '94643001', '94644007', '94645008', '94646009', '94647000', '94648005', '94649002', '94650002', '94651003', '94652005', '94653000', '94654006', '94655007', '94656008', '94657004', '94658009', '94659001', '94660006', '94661005', '94662003', '94663008', '94664002', '94665001', '94666000', '94667009', '94668004', '94669007', '94670008', '94671007', '94672000', '94673005', '94674004', '94675003', '94676002', '94677006', '94678001', '94679009', '94680007', '94681006', '94682004', '94683009', '94686001', '94687005', '94688000', '94690004', '94704006', '94707004', '94708009', '94709001', '94710006', '94711005', '94712003', '94714002', '94715001', '94716000', '94718004', '94719007', '95186006', '95187002', '95188007', '95192000', '95193005', '95194004', '95209008', '95210003', '95214007', '95224004', '95225003', '95226002', '95230004', '95231000', '95256006', '95257002', '95258007', '95259004', '95260009', '95261008', '95262001', '95263006', '95264000', '9541000119105', '956331000000107', '956341000000103', '956351000000100', '956361000000102', '956371000000109', '956381000000106', '956391000000108', '956401000000106', '956411000000108', '956421000000102', '956431000000100', '956441000000109', '956451000000107', '956461000000105', '956471000000103', '956481000000101', '956511000000107', '956521000000101', '956531000000104', '956541000000108', '956551000000106', '956561000000109', '9618003', '96281000119107', '96291000119105', '966311000000107', '966321000000101', '96981000119102', '97361000119109', '9801004', '9903002', '99101000119101', '99111000119103', '99121000119105', '99131000119108', '999000'] |
| **Stroke** | ['64009001', '230718007', '237702003', '277325005', '195200006', '230704000', '195236009', '270907008', '230695002', '195216008', '230693009', '231231000000107', '275526006', '195201005', '230694003', '195232006', '195234007', '810991000000109', '195160000', '195189003', '206196005', '276219001', '195241001', '230700009', '240313004', '277328007', '195209007', '277319006', '75543006', '1386000', '450410005', '230713003', '7713009', '519751000000106', '230698000', '195212005', '413102000', '230708002', '277315000', '230701008', '206398001', '230696001', '432504007', '390936003', '307766002', '277324009', '195190007', '71444005', '195199008', '62914000', '230712008', '195185009', '95454007', '195213000', '195154000', '230690007', '195211003', '230709005', '230707007', '206188000', '30400005', '306803007', '195243003', '288276001', '20908003', '373606000', '1032761000000105', '233983001', '195235008', '230706003', '277322008', '195182007', '95460007', '125081000119106', '195210002', '195167002', '276722003', '307767006', '276648002', '274100004', '195169004', '195180004', '450418003', '195231004', '275434003', '20059004', '195240000', '699270006', '39925003', '266253001', '195168007', '70611002', '112901000000108', '308067002', '56453003', '277299009', '812041000000104', '213208008', '230692004', '72986009', '230691006', '230711001', '195183002', '195165005', '95457000', '307363008', '213209000', '277303004', '195186005', '277286006', '277320000', '277329004', '75038005', '277330009', '195230003', '195233001', '38023001', '56267009', '281240008', '230714009', '21454007', '186893003', '195155004', '230710000', '49422009', '195217004', '195239002', '277316004', '230715005', '230699008', '52201006', '291111000119105', '90165008', '291411000119104', '126011000119107', '690201000119109', '42970005', '275363001', '703156006', '722628000', '329451000119104', '206576006', '329561000119101', '735115007', '431266005', '699429007', '12237951000119105', '292861000119106', '34135005', '703184001', '291571000119106', '127308007', '3681008', '97531000119106', '722623009', '254775002', '408664007', '734961002', '734963004', '17667005', '735131004', '276649005', '65587001', '195176009', '192763001', '281864001', '88747000', '34181000119102', '26954004', '62973004', '16002031000119102', '110997000', '329371000119101', '127311008', '230223008', '290641000119107', '95453001', '127310009', '441887006', '291091000119102', '674361000119104', '428268007', '291371000119100', '738779002', '209978003', '723141009', '16000391000119104', '762633000', '762652006', '672531000119106', '89142007', '762648006', '291531000119108', '4332009', '703313007', '200330000', '703207000', '330791000119108', '80901002', '312586003', '73173006', '291621000119109', '43216008', '590005', '230724001', '723857007', '149821000119103', '702374000', '292621000119100', '734959006', '703208005', '397809001', '133991000119109', '95456009', '142851000119103', '79341000119107', '724993002', '63795001', '762632005', '716051003', '287731003', '87020000', '230716006', '230732009', '276284000', '722615007', '18058007', '15978631000119109', '206395003', '69178005', '16418006', '237867001', '722619001', '78477003', '16023991000119104', '329491000119109', '276278003', '90520006', '209956005', '73439007', '111601003', '192757005', '723138000', '290631000119103', '450376009', '1089421000000105', '16371781000119100', '230702001', '111615001', '1055001', '425932008', '276650005', '936271000000100', '28318001', '721412003', '33331000119103', '291631000119107', '291351000119109', '276269009', '674401000119108', '12237911000119109', '286742002', '425957003', '266257000', '106016005', '9901000119100', '14070001', '6729006', '276282001', '141151000119101', '443929000', '59648004', '281865000', '276277008', '1078001000000105', '16000351000119109', '204497008', '195205001', '698838008', '329421000119107', '722631004', '34501004', '292631000119102', '762651004', '87253004', '703163006', '276271009', '111677000', '425642008', '15710721000119104', '413758000', '192753009', '118961000119101', '698627005', '111593004', '674121000119107', '42994005', '16218291000119100', '705130002', '450425005', '21202004', '86182004', '48601002', '302880005', '65189006', '40276003', '724425005', '290931000119108', '230703006', '45502001', '15742000', '703267003', '90099008', '444172003', '291511000119103', '200332008', '16061002', '16026991000119107', '65084004', '65312002', '441960006', '672461000119104', '762630002', '70607008', '111297002', '95458005', '674161000119102', '703161008', '276276004', '230221005', '672511000119101', '276221006', '18761000119108', '206417009', '43658003', '89980009', '16000511000119103', '302878004', '141811000119106', '62702001', '442668000', '1131000119105', '297176007', '276281008', '723139008', '253194008', '7931000119101', '88922007', '78569004', '195206000', '302881009', '428561000', '15982271000119104', '674391000119106', '442097001', '371041009', '209958006', '762649003', '408665008', '737232003', '16024031000119100', '724994008', '111673001', '111668007', '710575003', '195229008', '276651009', '1430001', '722930000', '95459002', '140281000119108', '60706008', '442181008', '45639009', '427020007', '140711000119106', '276280009', '703216001', '91601000119109', '293831000119105', '28048009', '24654003', '206194008', '111679002', '82999001', '276275000', '32728005', '37955001', '192771002', '724357007', '672561000119103', '25004000', '230717002', '35486000', '73020009', '16000431000119109', '1089411000000104', '690051000119100', '698839000', '200258006', '118951000119103', '88755007', '703268008', '734397002', '329361000119107', '315048006', '720854004', '291721000119102', '81520002', '733199002', '88032003', '449020009', '460880006', '230224002', '25133001', '140921000119102', '14309005', '703217005', '276594006', '722581000', '690351000119103', '128609009', '54265003', '416792008', '291481000119105', '329671000119106', '40135004', '87555007', '722643003', '111611005', '451037005', '722582007', '430959006', '95461006', '276652002', '192767000', '422504002', '703311009', '111298007', '690341000119100', '192756001', '37943007', '262949005', '285171000119104', '46758000', '734396006', '56384000', '29635000', '277313007', '77498000', '116288000', '713265001', '302909007', '20276007', '209947002', '73390009', '735114006', '722620007', '16024151000119105', '33301000119105', '93468003', '6666006', '724097003', '5251007', '441894009', '145741000119101', '16024111000119109', '330421000119102', '721979005', '230287006', '330411000119109', '15985031000119102', '71266000', '672441000119103', '95644001', '192760003', '126941005', '40161000119102', '230735006', '87551000119101', '734879002', '724428007', '111597003', '293811000119100', '32112006', '700467001', '672571000119109', '95830009', '87345009', '7819003', '68267002', '34191000119104', '23276006', '302904002', '234142008', '690271000119104', '12853006', '48518008', '80758005', '84216001', '703312002', '441735003', '95455008', '291581000119109', '703266007', '722909009', '71074007', '724429004', '16260551000119106', '451039008', '723085008', '426033005', '64775002', |

| | |
|---|---|
| | '472746006', '18751000119106', '12242711000119109', '12367511000119101', '28155008', '63323000', '734964005', '106021000119105', '29264003', '672521000119108', '429993008', '371121002', '103761000119107', '18322005', '200331001', '14246007', '69458009', '307362003', '725132001', '15978431000119106', '290791000119105', '75138007', '276706004', '209940000', '722633001', '78879009', '230720005', '734965006', '315047001', '10349009', '440140008', '57998008', '315046005', '329571000119107', '292671000119104', '722630003', '427065003', '128218002', '192759008', '200260008', '722632006', '301765007', '14977000', '52902005', '672551000119100', '703158007', '276283006', '70861009', '291521000119105', '442676003', '40549004', '133981000119106', '285161000119105', '206397006', '451038000', '84792001', '721328009', '25772007', '276274001', '329461000119102', '276220007', '276273007', '15988351000119101', '329431000119105', '234149004', '442024001', '292691000119103', '38453000', '304831001', '291591000119107', '59633005', '329651000119102', '16023911000119108', '276647007', '16024271000119107', '302213007', '206419007', '276286003', '111605007', '609382000', '724424009', '444657001', '18605003', '288723005', '441526008', '99051000119101', '732923001', '137991000119103', '230731002', '722580004', '75507000', '704079000', '48248005', '230220006', '703193000', '141281000119101', '140911000119109', '371040005', '35672006', '206396002', '26205001', '292681000119101', '723084007', '329481000119106', '111681000', '102831000119104', '192765008', '290581000119101', '723082006', '91216000', '441630004', '18485009', '427296003', '23671000119107', '23808003', '141831000119101', '737231005', '451035002', '734960001', '690331000119109', '262955000', '23819000', '427432001', '87937009', '13289004', '6594005', '118971000119107', '735132006', '237701005', '70936005', '261808007', '15710641000119100', '29322000', '291665000', '127312001', '141821000119104', '703206009', '690071000119109', '230723007', '705128004', '276285004', '262940009', '716745004', '429743002', '361000119103', '92341000119107', '200333003', '718551002', '206195009', '291121000119103', '35386004', '141091000119105', '73308006', '58020007', '703174002', '703220002', '262717000', '143521000119103', '46421000119102', '703176000', '21258007', '230725000', '690171000119105', '277196008', '42429001', '703218000', '330081000119108', '442733008', '297138001', '722906002', '55382008', '426788002', '722929005', '192754003', '36179005', '192764007', '722575008', '722584008', '73192008', '2495006', '441759008', '78757009', '45659008', '111028009', '722583002', '4262001', '127309004', '329641000119104', '315049003', '292661000119105', '703226008', '16907002', '99451000119105', '330091000119106', '442212003', '15705007', '266254007', '43262000', '703180005', '426651005', '230225001', '276270005', '230722002', '206191000', '426814001', '441991000', '206399009', '724427002', '292851000119109', '734384004', '230285003', '722624003', '15978471000119109', '703205008', '230523009', '230730001', '50751005', '37134004', '699706000', '291681000119108', '61687004', '433183000', '276224003', '762629007', '77768006', '128171000119104', '15982311000119104', '15988391000119106', '723083001', '134771000119108', '290621000119101', '722703002', '192755002', '425882004', '724426006', '724779000', '230738008', '230222003', '29941000119105', '703215002', '26021000119107', '90178008', '40450001', '705129007', '16026951000119102', '192769002', '15710681000119105', '262952002', '230719004', '66976009', '702575003', '734880004', '108691000119102', '186317009', '703221003', '674381000119108', '192761004', '43602006', '230721009', '111299004', '186831000119104', '55734000', '204501003', '371050006', '430947007', '722629008', '69116000', '8269002', '28366008', '301764006', '297157005', '12242751000119105', '61642003', '713081000', '234005004', '734383005', '291711000119109', '57981008', '442617003', '148871000119109', '111671004', '128608001', '713082007', '429235008', '291541000119104', '209987007', '140881000119109', '140701000119108', '703219008', '16002111000119106', '277311009', '200259003', '426107000', '441529001', '428241007', '632611000000107', '269147009', '706691000000101', '209981008', '417791000000102', '619831000000101', '1032681000000109', '491301000000100', '14936004', '670211000000106', '670161000000101', '439121000000104', '586561000000106', '529421000000101', '262954001', '670221000000100', '670231000000103', '426661000000105', '63590009', '195226001', '532071000000103', '209943003', '471931000000103', '876451000000104', '402531000000107', '397591000000103', '632631000000104', '710053005', '692701000000109', '878921000000101', '272701000000103', '878911000000107', '262739003', '65971006', '206673009', '269146000', '856891000000108', '209959003', '209944009', '209982001', '431501000000102', '402521000000105', '876441000000102', '415151000000100', '615841000000107', '419711000000101', '614911000000103', '78658006', '209979006', '431511000000100', '619811000000109', '682621000000105', '262950005', '288272004', '209961007', '1508000', '532081000000101', '632601000000105', '527981000000104', '431611000000101', '206624000', '530681000000107', '530691000000109', '431981000000102', '262951009', '670171000000108', '857261000000101', '664231000000102', '631871000000105', '586571000000104', '812051000000101', '508311000000103', '584201000000101', '478941000000104', '530711000000106', '632641000000108', '82894007', '307213001', '467951000000108', '510141000000103', '670251000000105', '444151000000109', '532091000000104', '7602002', '209995006', '28837001', '192781000000100', '670191000000107', '209938005', '444531000000107', '467401000000101', '1032691000000106', '419691000000103', '509371000000101', '758321000000100', '567211000000105', '530721000000100', '584151000000106', '858181000000100', '209960008', '450951000000100', '69533002', '209963005', '469471000000100', '13410009', '530701000000109', '685631000000102', '398283005', '471041000000108', '584161000000109', '401103008', '411911000000103', '413751000000100', '891101000000105', '1051081000000109', '670181000000105', '874711000000100', '419701000000103', '1032771000000103', '410064000', '586551000000108', '856601000000100', '832161000000105', '209980009', '401961000000103', '756921000000102', '632261000000107', '209976004', '467911000000109', '528471000000107', '670261000000108', '509351000000105', '209933001', '639511000000102', '632621000000101', '206418004', '581761000000107', '700251000000105', '313242003', '209941001', '831191000000105', '632651000000106', '209942008', '670271000000101'] |
| **Chronic kidney disease** | ['864311000000105', '865861000000101', '236538004', '276983001', '71192002', '866991000000105', '236566007', '866951000000102', '278691000', '236565006', '233580005', '236571000', '175901007', '236589004', '236567003', '233578004', '844661000000109', '216933008', '236564005', '847791000000101', '238316008', '233576000', '276883000', '19765000', '233577009', '233583007', '87235005', '236579003', '233469007', '236551009', '236558003', '236562009', '236570004', '288182009', '846761000000109', '180273006', '233495009', '233482004', '233592005', '236138007', '233581009', '236576005', '238315007', '865781000000101', '216932003', '233491000', '843691000000100', '427053002', '233585000', '236584009', '872481000000106', '233483009', '216878005', '182753006', '70536003', '302497006', '180277007', '233476002', '866041000000104', '236578006', '233496005', '175873005', '865981000000103', '866061000000103', '233587008', '233484003', '182750009', '236546003', '236552002', '161665007', '236588007', '238314006', '236549005', '105502003', '236556004', '238321006', '846731000000104', '238322004', '438546008', '225230008', '180272001', '265763003', '27929005', '233575001', '32599008', '175779004', '233579007', '846671000000106', '238319001', '269698004', '225231007', '236436003', '867011000000102', '865761000000105', '175777002', '236548002', '711411006', '236563004', '108241001', '233480007', '236583003', '847881000000107', '430332005', '852981000000100', '233477006', '864271000000105', '846461000000102', '233481006', '866001000000102', '236577001', '277011002', '847811000000100', '175780001', '233582002', '282348002', '438548009', '867051000000103', '711413009', '236539007', '236580000', '82801007', '271418008', '88351001', '251859005', '175788008', '236543006', '233468004', '236575009', '236555000', '427992007', '233485002', '865841000000102', '236435004', '233478001', '872461000000102', '238323009', '276998004', '236557008', '236573002', '265538001', '865801000000100', '236540009', '854131000000107', '854161000000102', '236569000', '33461007', '238317004', '233590002', '276897008', '846801000000104', '233487005', '236544000', '236574008', '236559006', '236547007', '236561002', '233589006', '175902000', '233584001', '853651000000105', '236581001', '426340003', '313030004', '233486001', '853631000000103', '236614007', '449288005', '236542001', '236554001', '233588003', '853021000000108', '236582008', '236587002', '236434000', '866021000000106', '866081000000107', '265764009', '236550005', '236560001', '236572007', '277010001', '129214009', '233586004', '429451003', '46177005', '699235009', '233488000', '867031000000105', '428648006', '236541008', '715743002', '708931003', '381000124105', '765479007', '73257006', '233595007', '473397008', '405331002', '57274006', '68341005', '233574002', '708933000', '443143006', '300478000', '699070001', '431873008', '331000124109', '405419000', '698074000', '14130001000004103', '719501003', '676002', '11000771000119104', '14684005', '11932001', '704667004', '765478004', '621000124102', '90791000119104', '704308002', '708930002', '90771000119100', '233573008', '429075005', '127991000119101', '461000124106', '844011000000103', '251869004', '442326005', '440597004', '431000124102', '341000124104', '444887005', '702635003', '78922001', '34897002', '232999000', '3257008', '451000124109', '571000124100', '581000124102', '233572003', '251285001', '473193004', '428937001', '471000124104', '611000124105', '601000124107', '443596009', '175899003', '213150003', '591000124104', '341939001', '708934006', '182749009', '365399009', '473398003', '125141000119102', '286371000119107', '708932005', '11000731000119102', '128001000119105', '428982002', '406168002', '67970008', '702634004', '441000124107', '714749008', '855161000000103', '866011000000100', '266901000000105', '844671000000102', '366961000000102', '856221000000106', '260981000000102', '244811000000102', '523481000000103', '857581000000107', '316054003', '866741000000102', '856931000000103', '1050711000000105', '834931000000108', '467521000000106', '219601000000106', '622431000000104', '236568008', '523471000000100', '327751000000105', '225371000000104', '856201000000102', '847891000000109', '640091000000107', '404471000000102', '856191000000104', '364571000000105', '866051000000101', '866961000000104', '853661000000108', '876311000000107', '855901000000104', '780091000000107', '865851000000104', '843701000000100', '398481000000100', '640081000000105', '853641000000107', '866761000000101', '342071000000104', '640141000000101', '864281000000107', '233181000000101', '864321000000104', '266921000000101', '856691000000107', '855271000000103', '231691000000106', '231631000000105', '399241000000100', '855091000000105', '637691000000101', '261001000000101', '830901000000105', '872491000000108', '364591000000109', '523461000000107', '288004005', '867061000000100', '231721000000102', '866031000000108', '830871000000105', '260991000000100', '643501000000102', '225441000000104', '643511000000100', '865871000000108', '853031000000105', '855951000000103', '856211000000100', '637681000000103', '180268000', '834961000000103', '426361000000104', '426351000000102', '410511000000103', '855481000000100', '857241000000102', '856161000000105', '291421000000102', '856681000000105', '856701000000107', '866751000000104', '698810000', '371181000000109', '236537009', '856231000000108', '847821000000106', '622441000000108', '938461000000107', '866771000000108', '261991000000109', '429211000000100', '261981000000107', '284451000000108', '856711000000109', '855231000000100', '637641000000106', '865791000000104', '438503005', '48994000', '309819001', '938451000000109', '640131000000105', '830931000000104', '266931000000104', '830881000000107', '872471000000109', '857311000000108', '865811000000103', '236553007', '847801000000102', '866071000000105', '266911000000107', '855521000000100', '867001000000104', '364701000000105', '866091000000109', '867041000000101', '371191000000106', '857631000000109', '267261000000102', '525981000000106', '855861000000106', '342221000000108', '830891000000109', '223178003', '398471000000102', '637661000000107', '857571000000105', '867021000000108', '523491000000101', '706901000000104', '310435001', '865991000000101', '865771000000103'] |

### Table S4 | Discrimination performance with targeted versus untargeted AI-ECG screening in YNHHS (temporal validation set).

| Label | Strategy | Prevalence | F1 | Sensitivity | Specificity | PPV | NPV | FN | FP |
|---|---|---|---|---|---|---|---|---|---|
| Aortic root ≥ 4 cm | Untargeted (AI-ECG alone) | 0.076 | 0.262 [0.230–0.290] | 0.431 [0.382–0.480] | 0.848 [0.838–0.858] | 0.188 [0.162–0.212] | 0.948 [0.941–0.955] | 215 [185–242] | 704 [657–752] |
| | Untargeted (AI-ECG + EHR) | | 0.304 [0.270–0.336] | 0.450 [0.400–0.501] | 0.876 [0.866–0.885] | 0.229 [0.199–0.257] | 0.951 [0.945–0.958] | 208 [180–235] | 572 [533–622] |
| | Targeted (EHR, then AI-ECG) | | 0.564 [0.523–0.605] | 0.614 [0.570–0.661] | 0.954 [0.948–0.960] | 0.523 [0.475–0.569] | 0.968 [0.963–0.973] | 146 [125–167] | 212 [186–241] |
| Aortic valve peak velocity ≥ 3 m/sec | Untargeted (AI-ECG alone) | 0.022 | 0.168 [0.121–0.221] | 0.305 [0.219–0.399] | 0.949 [0.942–0.955] | 0.116 [0.082–0.158] | 0.984 [0.980–0.987] | 73 [57–91] | 245 [216–277] |
| | Untargeted (AI-ECG + EHR) | | 0.205 [0.129–0.276] | 0.190 [0.116–0.264] | 0.985 [0.982–0.989] | 0.222 [0.140–0.311] | 0.982 [0.978–0.986] | 85 [69–103] | 70 [53–88] |
| | Targeted (EHR, then AI-ECG) | | 0.757 [0.685–0.824] | 0.610 [0.521–0.700] | 1.000 [1.000–1.000] | 1.000 [1.000–1.000] | 0.991 [0.989–0.994] | 41 [30–54] | 0 - |
| Aortic regurgitation (moderate/severe) | Untargeted (AI-ECG alone) | 0.032 | 0.156 [0.117–0.197] | 0.234 [0.173–0.300] | 0.941 [0.935–0.948] | 0.117 [0.086–0.153] | 0.974 [0.969–0.978] | 134 [110–158] | 309 [275–342] |
| | Untargeted (AI-ECG + EHR) | | 0.170 [0.122–0.221] | 0.183 [0.128–0.238] | 0.968 [0.963–0.973] | 0.159 [0.111–0.210] | 0.973 [0.968–0.977] | 143 [121–167] | 169 [144–197] |
| | Targeted (EHR, then AI-ECG) | | 0.619 [0.551–0.679] | 0.543 [0.469–0.612] | 0.993 [0.991–0.995] | 0.720 [0.633–0.800] | 0.985 [0.981–0.988] | 80 [64–100] | 37 [25–49] |
| Aortic stenosis (severe) | Untargeted (AI-ECG alone) | 0.008 | 0.091 [0.030–0.160] | 0.140 [0.044–0.244] | 0.985 [0.982–0.988] | 0.067 [0.021–0.121] | 0.993 [0.991–0.995] | 37 [27–50] | 83 [66–100] |
| | Untargeted (AI-ECG + EHR) | | 0.138 [0.049–0.226] | 0.140 [0.049–0.247] | 0.993 [0.991–0.995] | 0.136 [0.046–0.250] | 0.993 [0.991–0.995] | 37 [26–49] | 38 [26–50] |
| | Targeted (EHR, then AI-ECG) | | 0.838 [0.730–0.922] | 0.721 [0.575–0.855] | 1.000 [1.000–1.000] | 1.000 [1.000–1.000] | 0.998 [0.996–0.999] | 12 [6–20] | 0 - |
| E/e’ ≥ 15 | Untargeted (AI-ECG alone) | 0.093 | 0.398 [0.361–0.432] | 0.571 [0.518–0.623] | 0.867 [0.858–0.877] | 0.305 [0.274–0.339] | 0.952 [0.945–0.959] | 173 [146–197] | 523 [484–560] |
| | Untargeted (AI-ECG + EHR) | | 0.431 [0.389–0.472] | 0.439 [0.389–0.488] | 0.939 [0.931–0.945] | 0.423 [0.374–0.470] | 0.942 [0.935–0.950] | 226 [197–254] | 241 [214–270] |
| | Targeted (EHR, then AI-ECG) | | 0.663 [0.622–0.701] | 0.596 [0.546–0.645] | 0.979 [0.975–0.983] | 0.748 [0.704–0.790] | 0.959 [0.953–0.966] | 163 [138–189] | 81 [65–97] |
| Left ventricular ejection fraction ≤40% | Untargeted (AI-ECG alone) | 0.067 | 0.545 [0.504–0.586] | 0.548 [0.499–0.596] | 0.967 [0.962–0.972] | 0.541 [0.497–0.593] | 0.968 [0.963–0.972] | 163 [139–189] | 168 [142–192] |
| | Untargeted (AI-ECG + EHR) | | 0.537 [0.494–0.578] | 0.471 [0.419–0.522] | 0.980 [0.975–0.983] | 0.625 [0.573–0.677] | 0.963 [0.958–0.967] | 191 [167–219] | 102 [84–124] |
| | Targeted (EHR, then AI-ECG) | | 0.744 [0.702–0.783] | 0.598 [0.547–0.650] | 0.999 [0.998–1.000] | 0.982 [0.962–0.996] | 0.972 [0.967–0.976] | 145 [122–171] | 4 [1–8] |
| Left ventricular ejection fraction ≤50% | Untargeted (AI-ECG alone) | 0.112 | 0.579 [0.545–0.609] | 0.587 [0.551–0.626] | 0.944 [0.938–0.951] | 0.571 [0.533–0.609] | 0.947 [0.941–0.954] | 252 [220–284] | 269 [238–300] |
| | Untargeted (AI-ECG + EHR) | | 0.570 [0.536–0.605] | 0.497 [0.458–0.535] | 0.969 [0.964–0.974] | 0.669 [0.627–0.712] | 0.938 [0.932–0.945] | 307 [274–342] | 150 [127–174] |
| | Targeted (EHR, then AI-ECG) | | 0.676 [0.643–0.707] | 0.548 [0.507–0.585] | 0.991 [0.988–0.993] | 0.884 [0.853–0.916] | 0.945 [0.938–0.952] | 276 [241–310] | 44 [31–56] |
| Interventricular septal diameter ≥1.3 cm | Untargeted (AI-ECG alone) | 0.093 | 0.324 [0.293–0.355] | 0.432 [0.390–0.472] | 0.873 [0.864–0.882] | 0.259 [0.230–0.288] | 0.937 [0.929–0.944] | 272 [241–306] | 593 [549–634] |
| | Untargeted (AI-ECG + EHR) | | 0.348 [0.316–0.382] | 0.432 [0.390–0.478] | 0.892 [0.883–0.901] | 0.291 [0.260–0.325] | 0.939 [0.931–0.945] | 272 [242–305] | 504 [463–548] |
| | Targeted (EHR, then AI-ECG) | | 0.557 [0.515–0.596] | 0.507 [0.464–0.553] | 0.968 [0.963–0.973] | 0.618 [0.568–0.665] | 0.950 [0.944–0.956] | 236 [208–267] | 150 [126–175] |
| Interventricular septal diameter ≥1.5 cm | Untargeted (AI-ECG alone) | 0.027 | 0.218 [0.166–0.285] | 0.245 [0.183–0.323] | 0.972 [0.967–0.976] | 0.197 [0.148–0.262] | 0.979 [0.975–0.983] | 105 [84–126] | 139 [118–163] |
| | Untargeted (AI-ECG + EHR) | | 0.281 [0.215–0.354] | 0.273 [0.208–0.351] | 0.981 [0.977–0.985] | 0.290 [0.216–0.367] | 0.980 [0.976–0.984] | 101 [81–122] | 93 [74–113] |
| | Targeted (EHR, then AI-ECG) | | 0.722 [0.662–0.788] | 0.590 [0.517–0.677] | 0.999 [0.998–1.000] | 0.932 [0.879–0.978] | 0.989 [0.986–0.992] | 57 [41–71] | 6 [2–11] |
| Left atrial enlargement (moderate or severe) | Untargeted (AI-ECG alone) | 0.104 | 0.370 [0.341–0.398] | 0.561 [0.521–0.599] | 0.831 [0.820–0.841] | 0.277 [0.252–0.302] | 0.942 [0.935–0.949] | 250 [222–281] | 834 [781–887] |
| | Untargeted (AI-ECG + EHR) | | 0.356 [0.321–0.390] | 0.383 [0.346–0.421] | 0.911 [0.904–0.919] | 0.332 [0.297–0.367] | 0.927 [0.920–0.935] | 351 [315–387] | 438 [398–475] |
| | Targeted (EHR, then AI-ECG) | | 0.535 [0.495–0.573] | 0.441 [0.399–0.480] | 0.976 [0.971–0.980] | 0.678 [0.628–0.725] | 0.938 [0.931–0.944] | 318 [286–355] | 119 [98–142] |
| Left ventricular diastolic dysfunction (moderate or severe) | Untargeted (AI-ECG alone) | 0.059 | 0.338 [0.295–0.378] | 0.512 [0.454–0.572] | 0.904 [0.895–0.913] | 0.252 [0.215–0.289] | 0.967 [0.961–0.972] | 122 [102–144] | 379 [346–415] |
| | Untargeted (AI-ECG + EHR) | | 0.371 [0.319–0.420] | 0.392 [0.333–0.453] | 0.954 [0.947–0.960] | 0.351 [0.301–0.408] | 0.961 [0.955–0.967] | 152 [129–177] | 181 [158–208] |
| | Targeted (EHR, then AI-ECG) | | 0.711 [0.663–0.758] | 0.580 [0.525–0.642] | 0.997 [0.995–0.998] | 0.918 [0.875–0.956] | 0.974 [0.970–0.979] | 105 [85–123] | 13 [6–20] |
| Left ventricular internal diameter at end-diastole ≥ 5.5 cm | Untargeted (AI-ECG alone) | 0.088 | 0.375 [0.336–0.415] | 0.359 [0.318–0.407] | 0.947 [0.941–0.953] | 0.393 [0.348–0.439] | 0.939 [0.932–0.946] | 288 [256–322] | 249 [220–279] |
| | Untargeted (AI-ECG + EHR) | | 0.390 [0.346–0.432] | 0.339 [0.295–0.380] | 0.962 [0.957–0.967] | 0.461 [0.410–0.510] | 0.938 [0.931–0.945] | 297 [264–331] | 178 [154–203] |
| | Targeted (EHR, then AI-ECG) | | 0.600 [0.565–0.634] | 0.579 [0.539–0.621] | 0.966 [0.961–0.971] | 0.622 [0.578–0.665] | 0.960 [0.954–0.966] | 189 [162–216] | 158 [134–182] |
| Left ventricular internal diameter at end-systole ≥ 3.5 cm | Untargeted (AI-ECG alone) | 0.193 | 0.485 [0.456–0.510] | 0.502 [0.472–0.532] | 0.864 [0.853–0.874] | 0.469 [0.438–0.500] | 0.879 [0.869–0.888] | 491 [454–531] | 561 [517–607] |
| | Untargeted (AI-ECG + EHR) | | 0.482 [0.452–0.510] | 0.451 [0.419–0.483] | 0.899 [0.889–0.908] | 0.516 [0.484–0.547] | 0.872 [0.863–0.882] | 541 [500–583] | 417 [380–459] |
| | Targeted (EHR, then AI-ECG) | | 0.579 [0.548–0.607] | 0.517 [0.483–0.551] | 0.935 [0.928–0.942] | 0.656 [0.625–0.689] | 0.890 [0.881–0.899] | 476 [435–515] | 267 [237–297] |
| Left ventricular posterior wall thickness ≥ 1.3 cm | Untargeted (AI-ECG alone) | 0.067 | 0.304 [0.268–0.337] | 0.430 [0.382–0.477] | 0.899 [0.890–0.909] | 0.235 [0.204–0.267] | 0.956 [0.951–0.962] | 196 [173–221] | 483 [436–526] |
| | Untargeted (AI-ECG + EHR) | | 0.315 [0.272–0.356] | 0.352 [0.305–0.402] | 0.937 [0.928–0.943] | 0.285 [0.241–0.328] | 0.953 [0.946–0.958] | 223 [199–253] | 303 [271–344] |
| | Targeted (EHR, then AI-ECG) | | 0.573 [0.522–0.619] | 0.477 [0.417–0.528] | 0.987 [0.984–0.990] | 0.719 [0.658–0.773] | 0.963 [0.958–0.968] | 180 [158–207] | 64 [48–79] |
| Left ventricular posterior wall thickness ≥ 1.5 cm | Untargeted (AI-ECG alone) | 0.013 | 0.221 [0.131–0.306] | 0.265 [0.145–0.376] | 0.985 [0.981–0.988] | 0.189 [0.111–0.271] | 0.990 [0.987–0.992] | 50 [38–65] | 77 [61–96] |
| | Untargeted (AI-ECG + EHR) | | 0.234 [0.135–0.332] | 0.235 [0.130–0.352] | 0.990 [0.987–0.992] | 0.232 [0.133–0.336] | 0.990 [0.987–0.992] | 52 [39–66] | 53 [39–67] |
| | Targeted (EHR, then AI-ECG) | | 0.885 [0.825–0.938] | 0.794 [0.702–0.884] | 1.000 [1.000–1.000] | 1.000 [1.000–1.000] | 0.997 [0.996–0.998] | 14 [8–21] | 0 - |
| Left ventricular systolic dysfunction (moderate or greater) | Untargeted (AI-ECG alone) | 0.058 | 0.547 [0.502–0.597] | 0.587 [0.536–0.646] | 0.966 [0.961–0.971] | 0.513 [0.462–0.571] | 0.974 [0.970–0.979] | 124 [102–145] | 167 [144–193] |
| | Untargeted (AI-ECG + EHR) | | 0.547 [0.502–0.595] | 0.487 [0.440–0.542] | 0.982 [0.978–0.986] | 0.624 [0.567–0.683] | 0.969 [0.964–0.974] | 154 [129–177] | 88 [69–106] |
| | Targeted (EHR, then AI-ECG) | | 0.723 [0.676–0.766] | 0.567 [0.510–0.621] | 1.000 [1.000–1.000] | 1.000 [1.000–1.000] | 0.974 [0.969–0.979] | 130 [107–154] | 0 - |
| Left ventricular hypertrophy (moderate or greater) | Untargeted (AI-ECG alone) | 0.058 | 0.288 [0.241–0.336] | 0.316 [0.263–0.368] | 0.946 [0.940–0.953] | 0.265 [0.220–0.313] | 0.958 [0.952–0.963] | 197 [171–223] | 252 [220–281] |
| | Untargeted (AI-ECG + EHR) | | 0.263 [0.216–0.313] | 0.260 [0.208–0.316] | 0.956 [0.950–0.961] | 0.266 [0.214–0.324] | 0.955 [0.949–0.961] | 213 [184–239] | 207 [181–235] |
| | Targeted (EHR, then AI-ECG) | | 0.581 [0.527–0.627] | 0.510 [0.455–0.567] | 0.985 [0.981–0.988] | 0.674 [0.611–0.734] | 0.970 [0.966–0.975] | 141 [120–163] | 71 [56–88] |
| Mitral regurgitation (moderate or greater) | Untargeted (AI-ECG alone) | 0.076 | 0.357 [0.320–0.395] | 0.463 [0.417–0.513] | 0.907 [0.898–0.915] | 0.290 [0.257–0.327] | 0.953 [0.947–0.960] | 223 [193–252] | 470 [428–517] |
| | Untargeted (AI-ECG + EHR) | | 0.356 [0.313–0.400] | 0.376 [0.326–0.424] | 0.939 [0.932–0.947] | 0.338 [0.295–0.389] | 0.948 [0.942–0.954] | 259 [231–291] | 306 [267–341] |
| | Targeted (EHR, then AI-ECG) | | 0.569 [0.526–0.612] | 0.472 [0.423–0.520] | 0.985 [0.981–0.988] | 0.715 [0.663–0.770] | 0.958 [0.952–0.963] | 219 [191–248] | 78 [61–96] |
| Mitral stenosis (any)* | Untargeted (AI-ECG alone) | 0.011 | 0.115 [0.049–0.185] | 0.148 [0.062–0.240] | 0.984 [0.980–0.987] | 0.094 [0.039–0.154] | 0.990 [0.988–0.993] | 52 [39–67] | 87 [69–105] |
| | Untargeted (AI-ECG + EHR) | | 0.117 [0.049–0.204] | 0.115 [0.048–0.208] | 0.990 [0.988–0.993] | 0.119 [0.048–0.212] | 0.990 [0.987–0.993] | 54 [40–70] | 52 [39–67] |
| | Targeted (EHR, then AI-ECG) | | 0.827 [0.742–0.895] | 0.705 [0.590–0.809] | 1.000 [1.000–1.000] | 1.000 [1.000–1.000] | 0.997 [0.995–0.998] | 18 [10–27] | 0 - |
| Right ventricular enlargement (moderate or greater) | Untargeted (AI-ECG alone) | 0.042 | 0.320 [0.263–0.374] | 0.360 [0.295–0.423] | 0.961 [0.956–0.967] | 0.289 [0.235–0.345] | 0.972 [0.967–0.976] | 130 [109–150] | 180 [153–207] |
| | Untargeted (AI-ECG + EHR) | | 0.326 [0.260–0.390] | 0.276 [0.211–0.335] | 0.982 [0.978–0.986] | 0.397 [0.316–0.483] | 0.969 [0.964–0.974] | 147 [124–170] | 85 [66–104] |
| | Targeted (EHR, then AI-ECG) | | 0.723 [0.669–0.774] | 0.586 [0.520–0.652] | 0.998 [0.997–1.000] | 0.944 [0.907–0.983] | 0.982 [0.979–0.986] | 84 [68–101] | 7 [2–12] |
| | Untargeted (AI-ECG alone) | 0.147 | 0.489 [0.457–0.521] | 0.676 [0.630–0.714] | 0.812 [0.798–0.827] | 0.383 [0.351–0.415] | 0.936 [0.926–0.944] | 159 [136–183] | 534 [492–575] |

| | | | | | | | | | |
|---|---|---|---|---|---|---|---|---|---|
| Right ventricular systolic pressure ≥ 40 mm Hg | Untargeted (AI-ECG + EHR) | | 0.526 [0.488–0.564] | 0.553 [0.508–0.599] | 0.905 [0.895–0.915] | 0.501 [0.460–0.547] | 0.922 [0.912–0.932] | 219 [188–246] | 270 [240–298] |
| | Targeted (EHR, then AI-ECG) | | 0.699 [0.661–0.731] | 0.578 [0.530–0.620] | 0.987 [0.983–0.991] | 0.884 [0.846–0.918] | 0.931 [0.923–0.940] | 207 [180–234] | 37 [25–49] |
| Right ventricular systolic pressure ≥ 50 mm Hg | Untargeted (AI-ECG alone) | 0.062 | 0.351 [0.301–0.398] | 0.539 [0.474–0.607] | 0.899 [0.888–0.909] | 0.261 [0.217–0.302] | 0.967 [0.961–0.974] | 95 [76–114] | 315 [282–349] |
| | Untargeted (AI-ECG + EHR) | | 0.396 [0.339–0.447] | 0.432 [0.364–0.500] | 0.950 [0.943–0.958] | 0.365 [0.310–0.421] | 0.962 [0.956–0.969] | 117 [96–136] | 155 [132–177] |
| | Targeted (EHR, then AI-ECG) | | 0.757 [0.703–0.807] | 0.612 [0.544–0.678] | 1.000 [0.999–1.000] | 0.992 [0.969–1.000] | 0.975 [0.970–0.980] | 80 [64–97] | 1 [0–4] |
| Peak systolic velocity of the lateral tricuspid annulus (RV S') <10 cm/sec | Untargeted (AI-ECG alone) | 0.111 | 0.378 [0.341–0.411] | 0.411 [0.368–0.452] | 0.904 [0.896–0.913] | 0.349 [0.314–0.388] | 0.925 [0.916–0.933] | 281 [251–312] | 365 [329–397] |
| | Untargeted (AI-ECG + EHR) | | 0.375 [0.336–0.412] | 0.331 [0.293–0.371] | 0.946 [0.938–0.952] | 0.433 [0.384–0.477] | 0.919 [0.911–0.927] | 319 [288–352] | 207 [183–236] |
| | Targeted (EHR, then AI-ECG) | | 0.557 [0.516–0.594] | 0.430 [0.389–0.469] | 0.986 [0.982–0.989] | 0.792 [0.747–0.837] | 0.933 [0.925–0.940] | 272 [243–302] | 54 [41–68] |
| Right ventricular systolic dysfunction (moderate or severe) | Untargeted (AI-ECG alone) | 0.077 | 0.411 [0.366–0.454] | 0.438 [0.391–0.489] | 0.942 [0.935–0.949] | 0.387 [0.337–0.439] | 0.953 [0.946–0.959] | 209 [182–237] | 258 [228–288] |
| | Untargeted (AI-ECG + EHR) | | 0.413 [0.362–0.457] | 0.358 [0.305–0.403] | 0.969 [0.964–0.974] | 0.489 [0.428–0.547] | 0.948 [0.941–0.954] | 239 [209–270] | 139 [117–162] |
| | Targeted (EHR, then AI-ECG) | | 0.652 [0.606–0.697] | 0.511 [0.460–0.565] | 0.995 [0.993–0.997] | 0.900 [0.857–0.941] | 0.961 [0.955–0.966] | 182 [156–208] | 21 [12–30] |
| Tricuspid annular plane systolic excursion (TAPSE) <1.7 cm | Untargeted (AI-ECG alone) | 0.130 | 0.388 [0.351–0.420] | 0.428 [0.386–0.467] | 0.883 [0.873–0.893] | 0.354 [0.319–0.389] | 0.912 [0.902–0.921] | 331 [299–368] | 452 [416–491] |
| | Untargeted (AI-ECG + EHR) | | 0.352 [0.310–0.391] | 0.294 [0.255–0.332] | 0.944 [0.937–0.951] | 0.438 [0.389–0.492] | 0.899 [0.889–0.909] | 409 [369–449] | 218 [189–245] |
| | Targeted (EHR, then AI-ECG) | | 0.542 [0.500–0.580] | 0.409 [0.368–0.445] | 0.985 [0.981–0.989] | 0.803 [0.755–0.852] | 0.918 [0.909–0.926] | 342 [309–378] | 58 [42–73] |
| Tricuspid regurgitation (moderate or severe) | Untargeted (AI-ECG alone) | 0.085 | 0.403 [0.367–0.441] | 0.486 [0.439–0.532] | 0.914 [0.905–0.922] | 0.345 [0.307–0.382] | 0.950 [0.944–0.956] | 239 [210–267] | 430 [391–473] |
| | Untargeted (AI-ECG + EHR) | | 0.413 [0.370–0.452] | 0.430 [0.382–0.477] | 0.939 [0.932–0.946] | 0.398 [0.352–0.442] | 0.946 [0.940–0.953] | 265 [234–296] | 303 [266–340] |
| | Targeted (EHR, then AI-ECG) | | 0.600 [0.559–0.646] | 0.497 [0.453–0.548] | 0.985 [0.982–0.989] | 0.757 [0.710–0.807] | 0.955 [0.949–0.960] | 234 [204–262] | 74 [57–92] |

**Table S5 | Discrimination performance with targeted versus untargeted AI-ECG screening in the UK Biobank.**

| Label | Strategy | Prevalence | F1 | PPV | NPV | Sensitivity | Specificity | FN | FP |
|---|---|---|---|---|---|---|---|---|---|
| Aortic Root ≥4 cm | Untargeted (AI-ECG alone) | 0.005 | 0.018 [0.014–0.022] | 0.009 [0.007–0.011] | 0.996 [0.995–0.997] | 0.441 [0.366–0.514] | 0.754 [0.749–0.759] | 90 [71–111] | 7682 [7532–7826] |
| | Untargeted (AI-ECG + EHR) | | 0.022 [0.015–0.030] | 0.012 [0.008–0.016] | 0.996 [0.995–0.996] | 0.205 [0.146–0.272] | 0.910 [0.907–0.914] | 128 [106–152] | 2803 [2699–2902] |
| | Targeted (EHR, then AI-ECG) | | 0.121 [0.085–0.163] | 0.091 [0.064–0.125] | 0.996 [0.995–0.996] | 0.180 [0.126–0.244] | 0.991 [0.990–0.992] | 132 [110–158] | 290 [255–326] |
| Aortic Stenosis (any) | Untargeted (AI-ECG alone) | 0.004 | 0.033 [0.021–0.049] | 0.019 [0.012–0.028] | 0.997 [0.996–0.997] | 0.141 [0.092–0.199] | 0.972 [0.971–0.974] | 134 [111–158] | 1141 [1083–1209] |
| | Untargeted (AI-ECG + EHR) | | 0.065 [0.038–0.091] | 0.043 [0.026–0.061] | 0.997 [0.996–0.997] | 0.128 [0.078–0.182] | 0.989 [0.988–0.990] | 136 [115–159] | 441 [403–481] |
| | Targeted (EHR, then AI-ECG) | | 0.229 [0.157–0.301] | 0.403 [0.274–0.525] | 0.997 [0.996–0.997] | 0.160 [0.104–0.223] | 0.999 [0.999–0.999] | 131 [108–154] | 37 [25–49] |
| LVEF ≤40% | Untargeted (AI-ECG alone) | 0.007 | 0.190 [0.144–0.246] | 0.287 [0.217–0.365] | 0.994 [0.993–0.995] | 0.142 [0.104–0.185] | 0.998 [0.997–0.998] | 200 [174–228] | 82 [64–100] |
| | Untargeted (AI-ECG + EHR) | | 0.072 [0.030–0.121] | 0.500 [0.250–0.714] | 0.993 [0.992–0.994] | 0.039 [0.016–0.067] | 1.000 [1.000–1.000] | 224 [198–254] | 9 [4–16] |
| | Targeted (EHR, then AI-ECG) | | 0.241 [0.181–0.308] | 0.614 [0.492–0.745] | 0.994 [0.993–0.995] | 0.150 [0.108–0.197] | 0.999 [0.999–1.000] | 198 [173–227] | 22 [13–31] |
| LA dilation | Untargeted (AI-ECG alone) | 0.005 | 0.024 [0.020–0.029] | 0.012 [0.010–0.015] | 0.999 [0.999–1.000] | 0.879 [0.827–0.928] | 0.690 [0.685–0.695] | 17 [10–25] | 10007 [9841–10177] |
| | Untargeted (AI-ECG + EHR) | | 0.048 [0.038–0.057] | 0.025 [0.020–0.030] | 0.998 [0.998–0.999] | 0.589 [0.509–0.671] | 0.899 [0.896–0.902] | 58 [43–72] | 3268 [3156–3375] |
| | Targeted (EHR, then AI-ECG) | | 0.347 [0.250–0.438] | 0.673 [0.527–0.800] | 0.997 [0.996–0.997] | 0.234 [0.159–0.309] | 1.000 [0.999–1.000] | 108 [89–132] | 16 [8–24] |
| LV hypertrophy (wall thickness) | Untargeted (AI-ECG alone) | 0.005 | 0.039 [0.032–0.047] | 0.020 [0.016–0.024] | 0.998 [0.998–0.999] | 0.745 [0.665–0.810] | 0.817 [0.813–0.821] | 42 [29–57] | 5983 [5859–6121] |
| | Untargeted (AI-ECG + EHR) | | 0.080 [0.064–0.094] | 0.043 [0.034–0.051] | 0.998 [0.997–0.998] | 0.552 [0.481–0.629] | 0.938 [0.935–0.941] | 74 [57–92] | 2027 [1947–2119] |
| | Targeted (EHR, then AI-ECG) | | 0.367 [0.287–0.439] | 0.516 [0.415–0.620] | 0.996 [0.996–0.997] | 0.285 [0.214–0.351] | 0.999 [0.998–0.999] | 118 [96–140] | 44 [32–56] |
| RV dilation (size) | Untargeted (AI-ECG alone) | 0.005 | 0.024 [0.008–0.039] | 0.015 [0.005–0.025] | 0.995 [0.994–0.996] | 0.055 [0.020–0.093] | 0.982 [0.980–0.983] | 154 [131–180] | 593 [547–640] |
| | Untargeted (AI-ECG + EHR) | | 0.034 [0.016–0.054] | 0.022 [0.011–0.036] | 0.995 [0.994–0.996] | 0.067 [0.032–0.108] | 0.985 [0.984–0.986] | 152 [128–179] | 481 [443–522] |
| | Targeted (EHR, then AI-ECG) | | 0.230 [0.169–0.298] | 0.326 [0.241–0.429] | 0.996 [0.995–0.997] | 0.178 [0.126–0.238] | 0.998 [0.998–0.999] | 134 [113–157] | 60 [43–74] |
| RVEF ≤35% | Untargeted (AI-ECG alone) | 0.003 | 0.067 [0.036–0.097] | 0.042 [0.023–0.062] | 0.997 [0.997–0.998] | 0.163 [0.092–0.230] | 0.988 [0.987–0.990] | 87 [71–107] | 387 [347–424] |
| | Untargeted (AI-ECG + EHR) | | 0.065 [0.013–0.120] | 0.102 [0.021–0.191] | 0.997 [0.996–0.998] | 0.048 [0.010–0.094] | 0.999 [0.998–0.999] | 99 [81–121] | 44 [32–58] |
| | Targeted (EHR, then AI-ECG) | | 0.174 [0.092–0.273] | 0.909 [0.714–1.000] | 0.997 [0.997–0.998] | 0.096 [0.049–0.159] | 1.000 [1.000–1.000] | 94 [75–115] | 1 [0–3] |
